# Trust boosts recovery of countries from COVID-19

**DOI:** 10.1101/2021.04.01.21254783

**Authors:** Timothy M. Lenton, Chris A. Boulton, Marten Scheffer

**Affiliations:** Global Systems Institute, University of Exeter, Exeter, UK; Wageningen University, Wageningen, The Netherlands; Santa Fe Institute, Santa Fe, NM, USA

## Abstract

Why have some countries suppressed waves of the COVID-19 pandemic much more effectively than others? We find that the decay rate of daily cases or deaths from peak levels varies by a factor of ∼40 between countries. This measure of country-level resilience to COVID-19 is positively correlated with trust within society, and with the adaptive increase in stringency of government interventions when epidemic waves occur. All countries where >40% agree “most people can be trusted” achieve a near complete reduction of new cases and deaths. In contrast, countries where governments maintain greater background stringency tend to be less trusting and less resilient. Building trust is therefore critical to resilience, both to epidemics and other unexpected disruptions, of which COVID-19 is unlikely to be the last.

## Main Text

One of the big questions in the wake of the COVID-19 pandemic is why some countries seem to have been so much more successful than others in suppressing the waves of infection and deaths. Here we explore this issue using well-established ecological measures for the response of complex systems to perturbation (*1*). ‘Resilience’ describes the rate of recovery of a system from perturbation back towards a presumed, pre-existing stable state – here zero infection and associated deaths – where rapid recovery equals high resilience. The dynamics of infectious disease are such that if the basic reproduction number (*2*), *R*_0_ (the number of secondary infections produced by a single infected individual) exceeds 1, then exponential growth of new cases will result – although the lag between primary and secondary infection events complicates the relationship between *R*_0_ and growth rate of cases (*3*). Estimates that COVID-19 has *R*_0_∼4.5 (*4*) imply the potential for rapid exponential spread. In natural populations, such explosive spread typically results in an infection spreading throughout a population, until acquired immunity and/or mortality stabilizes and ultimately reduces case numbers. In contemporary human societies confronted by COVID-19, a host of responses have been used to try and limit the effective reproduction number (*2*), *R*_e_ (the number of people that can be infected by an individual at any specific time) and hence the spread of infection. Government interventions to limit social contact have been shown to reduce *R*_e_ with a ∼1-3 week lag, although few have a statistically significant effect (*5*). When *R*_e_=1 is achieved, new cases peak. When *R*_e_<1 is achieved, case numbers will decay, and if *R*_e_ is also approximately constant, the decay is exponential (albeit again with a lag (*3*)) and the decay rate quantifies ‘resilience’ (*1*). Deaths should track new cases, with a lag, if mortality rate is constant. Usually, the original stable state of zero infection is not recovered when efforts to limit the spread of infection (*R*_e_) are relaxed and therefore cases may start to rise again. To characterize the social capacity to resist this temptation and maintain measures even if infections go down, we also consider the proportional ‘reduction’ from peak to minimum as a metric.

We take a deliberately coarse-grained approach to measuring resilience as whole-country recovery rates of COVID-19 new cases and deaths, inspired by the observation that these often appear close to exponential decays (Fig. S1). We analyzed openly available COVID-19 data for daily confirmed cases and deaths (*6*), and testing (*7*) (see Materials and Methods). Cases and deaths are normalized by population size, to consider cases/capita and deaths/capita, and the data is smoothed to minimize issues such as weekly cycles in reporting. Given the biases from large known variations in testing intensity (tests/capita) over time and space, we also considered cases/tests, where testing data is available. Up to 1 December 2020, 156 countries had exhibited at least one peak and then decay of cases/capita (of which 36 had experienced a second peak and decay), 151 countries had exhibited at least one peak and then decay of deaths/capita (of which 32 had experienced a second peak and decay), and 93 countries had sufficient testing data to determine at least one peak and then decay of cases/tests (of which 23 had experienced a second peak and decay). Timeseries for all countries and the three metrics are shown in Fig. S1. We filtered cases of reasonably exponential decay for further analysis (r^2^≥0.8) and included multiple instances of well-fitted recovery occurring in one country in the dataset. This gives n=177 decays for cases/capita, n=159 for deaths/capita, n=105 for cases/tests. In a few countries a minimum had not yet been reached by 1 December 2020, so the reduction dataset is smaller (cases/capita n=165, deaths/capita n=150, cases/tests n=101). As some variables are not normally distributed, we calculate Spearman’s rank correlations (*ρ*) throughout unless otherwise stated.

The relative measures of resilience (rate of decay) and (proportional) reduction of cases should be more reliably estimated than absolute case numbers but could be biased by variations in testing intensity across time and space. However, we find across countries and waves, resilience of cases/capita and cases/tests are strongly positively rank correlated (n=100, *ρ* =0.86, p<0.0001) with linear correlation gradient 0.88 (r^2^=0.94) indicating that cases/capita tend to decay slightly faster than cases/tests (Fig. S2A). Reduction of cases/capita and cases/tests are also strongly positively rank correlated (n=94, *ρ* =0.83, p<0.0001) with proportional reductions (linear correlation gradient 1.0, r^2^=0.98; Fig. S2C). As considering cases/tests restricts the sample size and does not qualitatively alter correlations to other factors (Table 1, Table S1), we focus on cases/capita herein.

**Table 1.** Factors correlating with resilience (decay rate) and reduction of COVID-19 cases and deaths across countries. Pairwise Spearman’s rank correlations.

| Explanatory variable | Resilience |  |  |  |  |  | Reduction |  |  |  |  |  |
| --- | --- | --- | --- | --- | --- | --- | --- | --- | --- | --- | --- | --- |
|  | Cases/capita |  |  | Deaths/capita |  |  | Cases/capita |  |  | Deaths/capita |  |  |
| | $\rho$ | p | n | $\rho$ | p | n | $\rho$ | p | n | $\rho$ | p | n |
| Population | -0.23 | <0.01 | 177 | -0.32 | <0.0001 | 159 | -0.22 | <0.01 | 165 | -0.27 | <0.001 | 150 |
| Country size | -0.24 | <0.01 | 175 | -0.23 | <0.01 | 157 | (-0.14) | - | 163 | -0.18 | <0.05 | 148 |
| Population density | (0.08) | - | 175 | (0.02) | - | 157 | (-0.05) | - | 163 | (0.04) | - | 148 |
| GDP/capita | 0.21 | <0.01 | 170 | (0.03) | - | 153 | (0.08) | - | 158 | (0.0) | - | 144 |
| Median age | 0.26 | <0.001 | 172 | (0.09) | - | 154 | (0.08) | - | 160 | (-0.05) | - | 145 |
| Life expectancy | 0.26 | <0.001 | 175 | (0.15) | - | 157 | (0.05) | - | 163 | (0.0) | - | 148 |
| Human Dev. Index | 0.24 | <0.01 | 172 | (0.09) | - | 155 | (0.08) | - | 160 | (0.01) | - | 146 |
| Hospital beds | 0.29 | <0.001 | 156 | 0.19 | <0.05 | 142 | (0.14) | - | 145 | (0.06) | - | 134 |
| Mean stringency | -0.24 | <0.01 | 167 | -0.42 | <0.0001 | 155 | -0.41 | <0.0001 | 156 | -0.48 | <0.0001 | 146 |
| Decay stringency | 0.15 | <0.05 | 167 | (0.01) | - | 153 | -0.17 | <0.05 | 156 | -0.22 | <0.01 | 144 |
| Background stringency | -0.30 | <0.0001 | 167 | -0.51 | <0.0001 | 155 | -0.47 | <0.0001 | 156 | -0.56 | <0.0001 | 146 |
| Adaptive stringency | 0.47 | <0.0001 | 167 | 0.39 | <0.0001 | 153 | 0.29 | <0.001 | 156 | 0.21 | <0.05 | 144 |
| Trust | 0.43 | <0.0001 | 77 | 0.40 | <0.001 | 75 | 0.51 | <0.0001 | 72 | 0.48 | <0.0001 | 72 |
| Power distance | -0.34 | <0.001 | 109 | -0.28 | <0.01 | 102 | -0.22 | <0.05 | 105 | -0.27 | <0.01 | 97 |
| Individualism | 0.21 | <0.05 | 109 | 0.27 | <0.01 | 102 | (0.10) | - | 105 | 0.23 | <0.05 | 97 |
| Masculinity | (0.05) | - | 109 | (0.0) | - | 102 | (-0.10) | - | 105 | (-0.02) | - | 97 |
| Uncertainty avoidance | (-0.05) | - | 109 | -0.23 | <0.05 | 102 | -0.23 | <0.05 | 105 | -0.31 | <0.01 | 97 |
| Long-term orientation | 0.25 | <0.01 | 129 | (0.11) | - | 116 | (0.06) | - | 122 | (-0.01) | - | 110 |
| Indulgence | (0.01) | - | 129 | (-0.07) | - | 117 | (0.15) | - | 122 | 0.28 | <0.01 | 111 |

Resilience of cases/capita, measured as magnitude of decay rate, ranges by a factor of ∼40, from 0.16 d^-1^ (Mauritius; most resilient) to 0.0041 d^-1^ (Costa Rica; least resilient), corresponding to a half-life of ∼4 to ∼170 days (Fig. 1A). Resilience of deaths/capita, ranges by a factor of ∼25 from 0.10 d^-1^ (Slovakia; most resilient) to 0.0042 d^-1^ (Indonesia, Mexico, Romania; least resilient) (half-life ∼7 to 165 days) (Fig. 1B). Resilience of cases/capita and deaths/capita are positively rank correlated (n=150, *ρ* =0.61, p<0.0001) with linear correlation (gradient 0.95, r^2^=0.75) indicating cases tend to decay slightly faster than deaths (Fig. S2B).

**Fig. 1.**
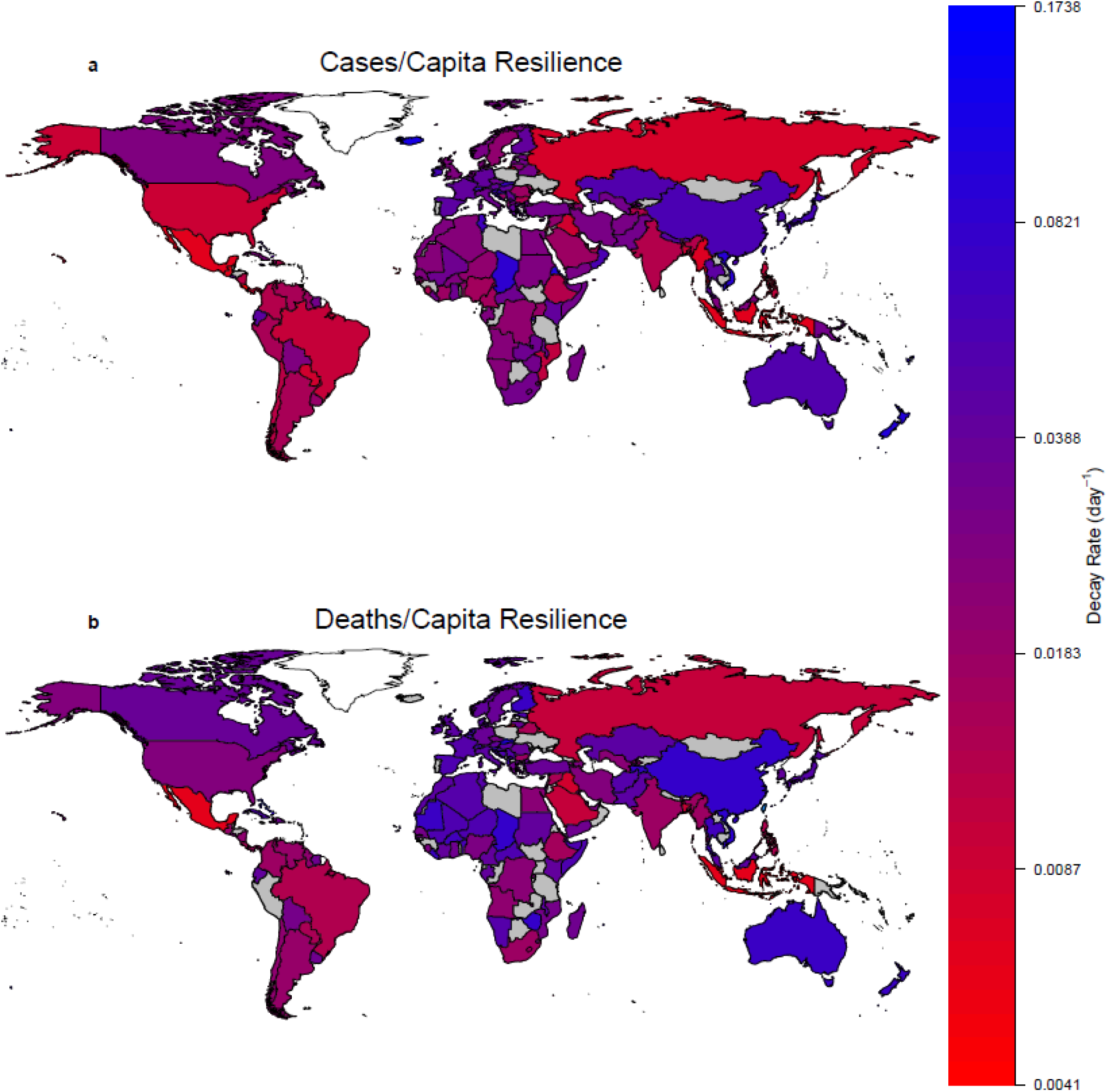
World maps of country-level resilience to COVID-19. Decay rate (d^-1^) from the first peak of: (**A**) cases/capita. (**B**) deaths/capita. In both cases countries are colored where the fit of an exponential decay has r^2^≥0.8. Countries in grey either have insufficient data or a poorer fit.

Reduction (from peak to next minimum) is strongly positively correlated with resilience, as expected mathematically, following a non-linear relationship (Fig. S3). Really high resilience >∼0.1 d^-1^ (half-life <∼1 week) tends to end in near complete reduction, but few countries have achieved this level of resilience. Instead several countries still achieve a near complete reduction of cases or deaths if they have a resilience of >∼0.02 d^-1^ (half-life <∼1 month). Below that threshold level of resilience, reduction inevitably drops. Thus, poor resilience leads to failure to eliminate cases and deaths. Reduction of cases/capita and deaths/capita are positively rank correlated (n=136, *ρ* =0.76, p<0.0001) with linear correlation (gradient 1.06, r^2^=0.96) indicating deaths tend to be reduced slightly more effectively than cases (Fig. S2D).

To interpret the resilience results, we focused on the potential role of social and cultural factors.

### Wealth

Demographic and public-health-related factors may be expected to influence country-level resilience, given that some significantly influence spread of the infection in within-country analysis (*8*). However, no particularly strong controls emerge (Table 1). There are weak negative correlations between population and resilience of cases/capita or deaths/capita, and between country area and resilience of cases/capita or deaths/capita, but these counteract, leaving no significant effect of population density (Table 1). Richer populations (GDP/capita) tend to have greater median age (n=175, *ρ* =0.84, p<0.0001), life expectancy (n=179, *ρ* =0.85, p<0.0001), human development index (HDI) (n=178, *ρ* =0.96, p<0.0001), and hospital beds (per 1000) (n=161, *ρ* =0.61, p<0.0001). This leads to shared significant weak positive correlations of GDP/capita, median age, life expectancy, HDI, and hospital beds (per 1000) with resilience of cases/capita (Table 1). However, only hospital beds (per 1000) show a significant weak positive correlation with resilience of deaths/capita, and none of these factors significantly correlate with reduction of cases/capita or deaths/capita (Table 1).

### Stringency

Deliberate government interventions to limit social contact and thus *R*_e_ are expected (*5*) to result in greater resilience (faster decay of cases and deaths). Hence we looked for relationships with the ‘stringency index’ from the Oxford COVID-19 Government Response Tracker (*9*), which is an additive score of indicators including school and workplace closures and restrictions of movement (see Materials and Methods). We considered ‘mean stringency’ across the whole time series since the start of the pandemic, ‘decay stringency’ averaged over the fitted cases/capita decay intervals, ‘background stringency’ averaged over the intervals when decay is not occurring, and ‘adaptive stringency’ – the change in stringency from before to during a decay interval. We only found a weak positive relationship between decay stringency and resilience (rate of decay) of cases/capita, no relationship for deaths/capita, and a weak negative relationship with reduction of cases/capita and deaths/capita (Table 1). Mean stringency is significantly negatively correlated with resilience of cases/capita and especially deaths/capita (Table 1). Background stringency is significantly and more strongly negatively correlated with resilience of cases/capita and deaths/capita, and especially with reduction of cases/capita and deaths/capita (Table 1). Only adaptive stringency (the change in stringency from before to during decay intervals) has a significant positive correlation with resilience of cases/capita (Fig. 2A) and deaths/capita (Fig. 2B), with significant but weaker positive correlations to reduction of cases/capita and deaths/capita (Fig. 2A,B; Table 1). Thus, deploying stringent measures decisively when an epidemic wave erupts is beneficial, but governments that maintain greater background and overall stringency have slower recovery and are less effective at reducing cases and deaths.

**Fig. 2.**
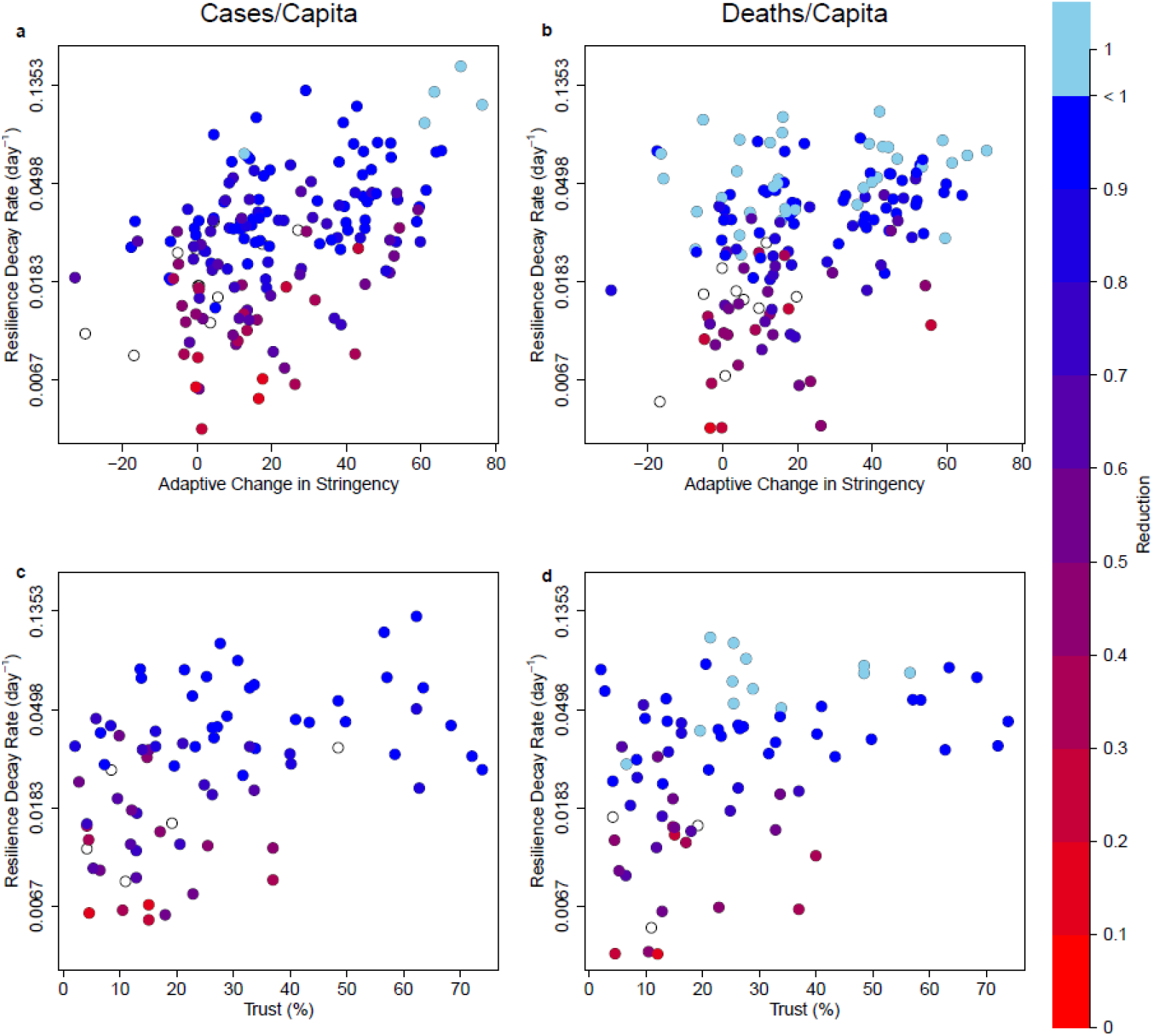
Country-level relationships between adaptive stringency or trust, resilience to COVID-19 and resulting reduction of cases and deaths. (**A**) cases/capita relationships between adaptive stringency and resilience (n=167, ⍰ =0.47, p<0.0001) and between adaptive stringency and reduction (n=156, ⍰ =0.29, p<0.001). (**B**) deaths/capita relationships between adaptive stringency and resilience (n=153, ⍰ =0.39, p<0.0001) and between adaptive stringency and reduction (n=144, ⍰ =0.21, p<0.05). (**C**) cases/capita relationships between trust and resilience (n=77, ⍰ =0.43, p<0.0001) and between trust and reduction (n=72, ⍰ =0.51, p<0.0001). (**D**) deaths/capita relationships between trust and resilience (n=75, ⍰ =0.40, p<0.001) and between trust and reduction (n=72, ⍰ =0.48, p<0.0001). Note the threshold effect whereby trust >40% (of population agreeing with the statement “most people can be trusted”) ensures resilience of cases/capita >0.02 d^-1^ and deaths/capita >0.03d^-1^, which in turn support successful reduction of cases and deaths. Cases of complete reduction – i.e. elimination of cases or deaths – are denoted with pale blue. Cases where reduction is incomplete at the end of the timeseries are denoted with open circles. The trust-reduction relationships are further analyzed in Fig. S4.

### Trust

We examined the effect of generalized trust using the World Values Survey (*10*) results (2017-20) for the percentage of correspondents who agree that “most people can be trusted” (available for 79 countries). Trust is significantly positively correlated (Table 1) with resilience of cases/capita (Fig. 2C) and deaths/capita (Fig. 2D) and especially with reduction of cases/capita and deaths/capita (Fig. 2C,D; Fig. S4). There is a clear threshold effect whereby trust >40% ensures sufficient resilience to end in a large or complete reduction of cases and deaths (Fig. 2C,D; Fig. S4A,B). Reduction distributions for trust ≤40% and trust >40% are significantly different (cases/capita Mann-Whitney p<0.001; deaths/capita Mann-Whitney p<0.0001; Fig. S4C, D). Trust and adaptive stringency are not significantly correlated, and in each case when controlling for one of the variables the resilience residuals remain strongly positively correlated with the other variable (for cases/capita resilience, the adaptive stringency correlation *ρ* goes from 0.428 to 0.449 and the trust correlation *ρ* goes from 0.432 to 0.472, for deaths/capita resilience, the adaptive stringency correlation *ρ* goes from 0.519 to 0.549 and the trust correlation *ρ* goes from 0.398 to 0.388). Trust is negatively correlated with mean stringency (n=71, *ρ* =-0.44, p<0.001) and background stringency (n=67, *ρ* =-0.47, p<0.0001), which may help explain why governments with greater background stringency are less effective at reducing COVID-19 cases and deaths – because they reflect less trusting societies. Trust supports economic growth (*11*) and hence has a well-known (*12*) positive correlation with GDP/capita (n=72, *ρ* =0.70, p<0.0001), which leads to positive correlations with median age (n=73, *ρ* =0.55, p<0.0001), life expectancy (n=74, *ρ* =0.57, p<0.0001), HDI (n=73, *ρ* =0.69, p<0.0001), and hospital beds (per 1000) (n=71, *ρ* =0.45, p<0.0001). Pairwise correlation results (Table 1) suggest that trust exerts a stronger control than any of these factors on resilience or reduction, but this could be influenced by the smaller sample of countries with trust data.

To examine this further we built various multiple linear regression models for resilience and reduction, with different mixes of the above factors (see Materials and Methods). Trust and adaptive stringency are consistently retained as the most significant beneficial factors. A model for resilience of cases/capita considering trust, adaptive stringency, GDP/capita, population, and hospital beds, retains adaptive stringency and trust as the most significant beneficial factors, followed by hospital beds, and rejects GDP/capita (n=71, r^2^=0.409; Table S2, Fig. S5A). A model for resilience of deaths/capita considering the same factors retains adaptive stringency and trust as the most significant beneficial factors, and GDP/capita as detrimental (n=69, r^2^=0.508; Table S3, Fig. S5B). A model for reduction of cases/capita retains trust and adaptive stringency and trust as the most significant beneficial factors, followed by hospital beds, with GDP/capita as detrimental (n=66, r^2^=0.352; Table S4, Fig. S5C). A model for reduction of deaths/capita retains adaptive stringency and trust as the most significant beneficial factors (n=66, r^2^=0.414; Table S5, Fig. S5d). These results confirm that trust and adaptive stringency are beneficial to resilience and reduction of both cases/capita and deaths/capita, and they suggest that trust gives rise to the significant pairwise positive correlation of GDP/capita and cases/capita resilience (Table 1) rather than vice versa.

We also considered whether resilience correlates with any of Hofstede’s six cultural dimensions (*13*) of power distance, individualism, uncertainty avoidance, masculinity, long-term orientation, and indulgence (see Materials and Methods). Power distance (expectation from the less powerful that power is distributed unequally) is anti-correlated with trust (n=49, *ρ* =-0.70, p<0.0001) and, consistent with that, decreases resilience of cases/capita and deaths/capita (Table 1). Individualism is positively correlated with trust (n=49, *ρ* =0.59, p<0.0001) and less strongly with resilience of cases/capita, and deaths/capita (Table 1). Long term orientation (pragmatism and preparation for the future) is positively correlated with trust (n=63, *ρ* =0.34, p<0.0001) and resilience of cases/capita but not deaths/capita (Table 1). Uncertainty avoidance is negatively correlated with trust (n=49, *ρ* =-0.43, p<0.01) and resilience of deaths/capita but not cases/capita (Table 1). Masculinity and indulgence do not show significant pairwise correlations with trust or resilience. Including Hofstede’s six cultural dimensions in place of trust in multiple linear regression models allows us to analyze a larger set of countries but explains less of their variance (Tables S6-S9). Mixing trust and the Hofstede dimensions in the models always retains trust as more significant than any of the retained Hofstede dimensions (Tables S10-S13).

Our results are robust to analyzing just the first peaks in each country (Table S14) or using a more stringent fitting of exponential decay (Table S15) – both of which reduce the sample size. The results support suggestions (*14, 15*) that variation in resilience to COVID-19 reflects, among other things, variation in the nature and strength of the ‘social contract’ across countries. Different theories of the social contract (*16*) encompass both reciprocal trust among citizens (*17*), and between citizens and their government (political elite) (*18, 19*). In the latter relationship, the individual surrenders some of their freedoms and submits to an authority in return for protection of their remaining rights, usually including the right to protection of life, and of a minimum standard of health (*20*). Resilience to the COVID-19 pandemic depends both on the action of governments (e.g. instigating social distancing measures) and the actions of citizens (e.g. complying or not with those measures). We find that adaptive stringency of government interventions is important to resilience, but resilience is also critically dependent on trust. More trusting societies tend to bring down cases and deaths faster and carry on with containment efforts more effectively until the full benefits are realized. A threshold level of >40% interpersonal trust in society, seems to guarantee effective resilience to COVID-19 and near complete reduction of deaths. There of course remains lots of unexplained variance, recognizing that environmental factors such as temperature (*21*), humidity (*21*), and UV exposure (*22*) may affect the spread of COVID-19, and that different strains of COVID-19 differ in their basic reproduction number (*8*). Despite all this we find a marked effect of trust, which adds to evidence that trust can support increased COVID-19 risk perception (*23*), decreased mortality early in the pandemic (*24*), and an earlier peak of new infections (*25*). Before the pandemic, the rise of neoliberalism since the 1980s and austerity policies since the 2008/9 financial crash had eroded the social contract between citizens and government in many nations (*15*). Most governments have responded to the pandemic with social protection policies to strengthen the social contract (*15*), and vaccination is also part of the social contract (*26*). Trust in institutions has increased in the short-term (*27-29*), but pandemics can erode trust in the long-term (*30*). Our results add to evidence that building trust within society is critical to resilience to epidemics (*31-33*). Trust also makes societies more resilient to other types of unexpected disruption (*34*). COVID-19 will surely not be the last.

## Data Availability

Data sources are listed in the Materials and Methods. R code and resilience results are available on request.

## Acknowledgments

This study would not have been possible without the commitment of all those who provide open data on the pandemic, and ‘Our World in Data’ for pulling relevant data together. We thank Fenna Blomsma for the idea to analyze the Hofstede dimensions.

## Author contributions

Conceptualization: TML. Methodology: TML, CAB, MS. Investigation: CAB, TML, MS. Visualization: CAB, TML. Funding acquisition: TML. Project administration: TML. Supervision: TML. Writing – original draft: TML. Writing – review & editing: TML, CAB, MS.

