## Supplementary Materials for "Trust boosts recovery of countries from COVID-19"

**This PDF file includes:**

Materials and Methods
Supplementary Text
Figs. S1 to S5
Tables S1 to S15

### Materials and Methods

Cases, deaths, tests data

All data were sourced from Our World in Data COVID-19 dataset (*35*). Raw data on daily confirmed cases and deaths for all countries is from the COVID-19 Data Repository by the Center for Systems Science and Engineering (CSSE) at Johns Hopkins University (*6*, *36*). Testing data is from Our World in Data (*7*). Note that the list of 'countries' includes the disputed territory of Kosovo (OWID_KOS). The world aggregate (OWID_WRL) is removed from our comparative analysis of countries.

We use a Kernal smoothing function with a bandwidth of 10 to smooth the daily timeseries of cases/capita, tests/capita, and deaths/capita. To create cases/tests timeseries, we divide the smoothed cases/capita timeseries by the test/capita timeseries. Peaks in timeseries were detected by eye, looking across all timeseries (cases/capita, deaths/capita, cases/tests) available for each country (Fig. S1). For a given country, peaks were manually lined up across timeseries (in some cases this means a peak may be considered a 'second peak' despite being the only peak in that specific time series due to lining up with a second peak in another timeseries from the same country). All instances of peaks and decays are grouped in a combined dataset (so the same country exhibiting multiple peaks and decays will appear more than once). Restricting the dataset to first peaks only does not notably affect the correlation results (compare Table S14 to Table 1).

Resilience and reduction

Resilience is estimated from the interval of data from a maximum to the next minimum in each smoothed timeseries (or the end of the timeseries if it happens first). The data were natural logged and linear regression used to determine the goodness of fit of an exponential decay. Most of the results cluster at $r^2 \geq 0.8$, with a clear drop off and scattering of $r^2$ values below 0.8. Visual inspection confirmed that $r^2 \geq 0.8$ captures cases of reasonably exponential decay for further analysis (red lines in Fig. S1). A more stringent cut-off of $r^2 \geq 0.9$ reduces the sample size but does not notably affect the range of resilience results, or their correlations to explanatory factors (compare Table S15 to Table 1).

Reduction is calculated as minimum divided by preceding maximum in the smoothed timeseries and is not reported if the fit of decay is poor ($r^2 < 0.8$ for Table 1, $r^2 < 0.9$ for Table S15), or if the end of the time series occurs before a minimum. The more stringent cut-off of $r^2 \geq 0.9$ does not notably affect the correlations of reduction results to explanatory factors (compare Table S15 to Table 1).

Government Stringency Index

Data for the Government Stringency Index are from the Oxford COVID-19 Government Response Tracker (*9*, *37*) (OxCGRT) as reported by Our World in Data (*35*). It is a composite measure based on nine response indicators, rescaled to a value from 0 to 100 (100 = strictest). If policies vary at the subnational level, the index is shown as the response level of the strictest sub-region. The response indicators are: school closing (C1), workplace closing (C2), cancel public events (C3), restrictions on gatherings (C4), close public transport (C5), stay at home requirements (C6), restrictions on internal movement (C7), international travel controls (C8), and public info campaigns (H1).

For each country, 'mean stringency' was calculated as the average across the whole time series since the start of the pandemic. 'Background stringency' was calculated as the average over the intervals when fitted decay intervals are not occurring. 'Decay stringency' was calculated as the average over each fitted decay interval. 'Adaptive stringency' was calculated for each decay interval as the difference from a 'pre stringency' to 'decay stringency' – where ‘pre stringency’ was averaged over the preceding interval, starting either at the start of the timeseries or at the end of a previous decay interval. All stringency metrics (except mean stringency which is the same) were calculated separately for cases/capita and deaths/capita (as they have separate decay intervals). However, even when considering resilience of deaths/capita the correlation results are comparable or better using stringency measures calculated for cases/capita – presumably because stringent policy interventions typically respond to cases data and the response of deaths lags weeks behind. Hence, we focus on stringency measures calculated for cases, even when considering resilience of deaths.

Demographic, financial, and public-health factors

Data were sourced from Our World in Data COVID-19 dataset (*35*). Population and life expectancy are from United Nations, Department of Economic and Social Affairs, Population Division, World Population Prospects 2019 Revision (*38*). Population density is from the World Bank – World Development Indicators (*39*). Country area is from dividing population by population density (to maintain internal consistency). GDP/capita is from the Maddison Project Database, version 2018 (*40*). Human Development Index (HDI) is from the United Nations Development Programme (UNDP) (*41*). Hospital beds (per 1000) for the most recent year available since 2010 is compiled by Our World in Data from multiple sources (*42*).

Trust

Trust data is from the World Values Survey (*10, 43*) Wave 7 (2017-20) Q57 and is the percentage of respondents who agree with the statement “most people can be trusted” (for 79 countries). This is often referred to in the literature as generalized trust, and sometimes as unspecified trust.

### Hofstede dimensions

Data for the Hofstede Dimensions is from the dimension data matrix (version 2015 12 08) (*13, 44*). The six dimensions are defined as follows: (i) *Power distance* is the extent to which the less powerful members of organizations and institutions (like the family) accept and expect that power is distributed unequally. (ii) *Individualism* is the inverse of the degree to which people in a society are integrated into groups. (iii) *Uncertainty avoidance* describes a society's tolerance for ambiguity where societies that score highly opt for stiff codes of behavior, guidelines, laws, and generally rely on absolute truth, or the belief that one lone truth dictates everything and people know what it is. (iv) *Masculinity* describes preference in society for achievement, heroism, assertiveness, and material rewards for success, whilst its counterpart represents a preference for cooperation, modesty, caring for the weak and quality of life. (v) *Long-term orientation* describes connection of the past with the current and future actions/challenges – societies with a high degree in this index view adaptation and circumstantial, pragmatic problem-solving as a necessity. (vi) *Indulgence* describes the degree of freedom that societal norms give to citizens in fulfilling their human desires.

### Pairwise regressions

We use Spearman’s rank correlation coefficient because not all variables considered are normally distributed and we wanted to detect any non-linear relationships.

### Multiple regression

Variables that are not normally distributed were first log transformed to achieve a normal distribution. The reduction distribution is strongly skewed and there was no improvement in transforming it. We experimented with different sets of independent variables, informed by the pairwise regression analysis in seeking to limit the number of independent variables. We considered both multiple linear regression and logistic regression models but found the logistic model fits were either slightly worse (resilience) or comparable (reduction). Hence for simplicity we present multiple linear regression models throughout. Once models were fitted, we used the step() function in R to optimize the model by Akaike information criterion (AIC), by adding or removing variables until the optimum fit is found.

### Supplementary Text

Resilience results for cases/tests

Resilience of cases/tests, measured as the exponential decay rate (magnitude), ranges by a factor of ~40, from 0.10 $d^{-1}$ (Fiji, New Zealand; most resilient) to 0.0026 $d^{-1}$ (Indonesia; least resilient). Differences in resilience results between cases/capita and cases/tests were examined on a case-by-case basis using Our World in Data to visualize the data for testing intensity by country over time. New Zealand shows a comparable decay rate for cases/capita, despite testing intensity increasing through the first wave. Iceland, which has a fast decay of cases/capita (0.13 $d^{-1}$) has a slower decay for cases/tests (0.085 $d^{-1}$), because testing intensity tended to track cases albeit with a lag. Indonesia's interval of declining cases in September-October interrupts an overall increase. Cases/tests show an earlier peak and longer, slower decline than cases/capita (0.0063 $d^{-1}$) thanks to fluctuating testing levels. Costa Rica which had the slowest decay rate of cases/capita (0.0041 $d^{-1}$) shows a poor fit of cases/tests not least because testing data ceases whilst cases/capita were declining through September-October. This discussion serves to illustrate why cases/tests does not produce an improved resilience dataset compared to cases/capita.

That said, correlation results for hypothesized factors affecting resilience are generally consistent between cases/capita (Table 1) and cases/tests (Table S1) reinforcing the robustness of the overall messages. To summarize (Table S1): Resilience of cases/tests is not significantly correlated with population size, country size, or population density. Resilience of cases/tests is positively correlated with median age, life expectancy, HDI, and hospital beds (per 1000), and weakly with GDP/capita. There is no significant relationship between decay stringency and resilience of cases/tests. Resilience of cases/tests is significantly negatively correlated with mean stringency and background stringency. Adaptive stringency is strongly positively correlated with resilience of cases/tests. Trust is positively correlated with resilience of cases/tests. Resilience of cases/tests declines with power distance and increases with long term orientation and individualism.

Correlation results for factors affecting reduction generally mirror those for resilience (Table S1). In contrast to cases/capita (Table 1), significant effects of median age, life expectancy, HDI, hospital beds (per 1000), and GDP/capita carry over to reduction of cases/tests.

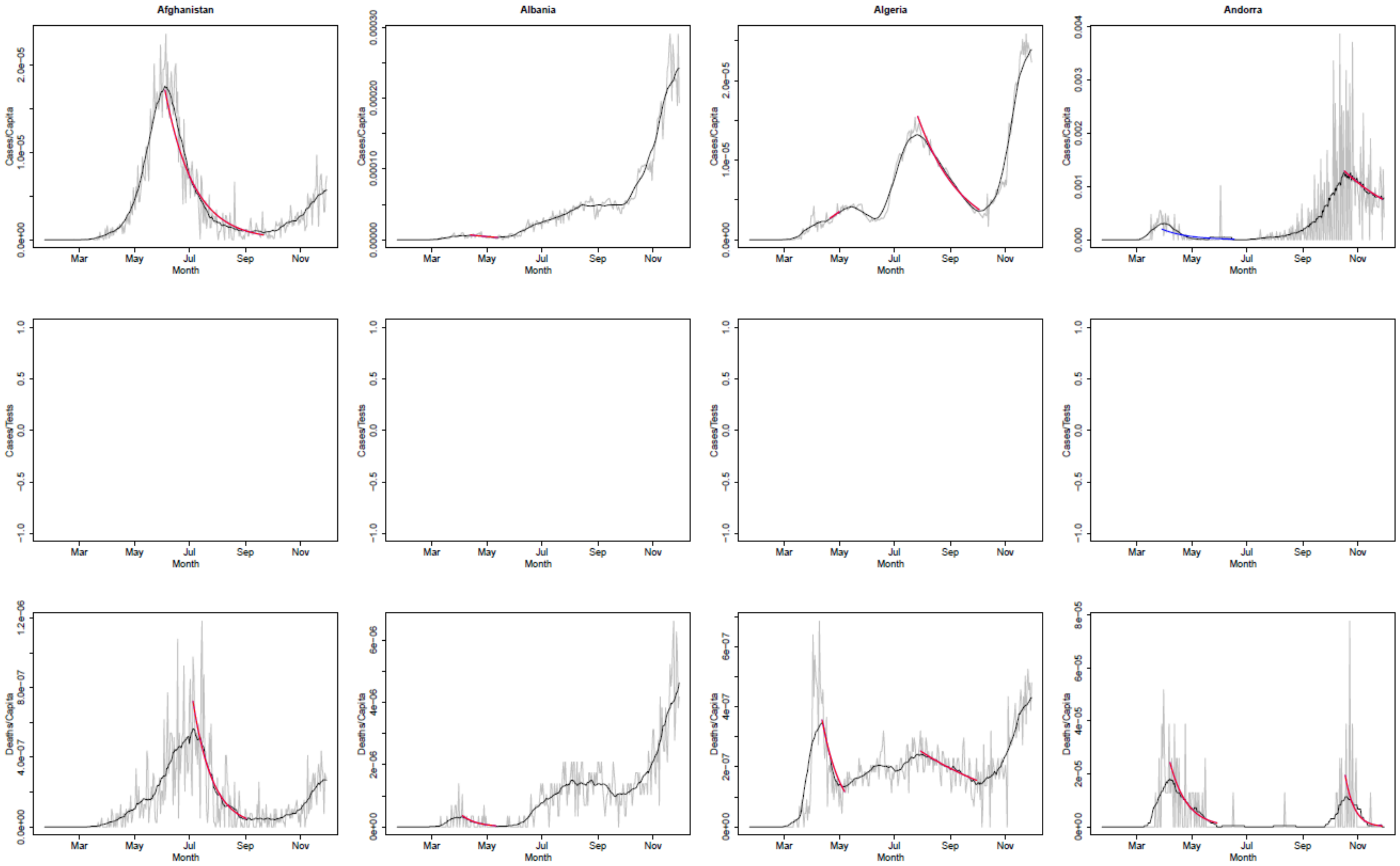

**Fig. S1.**
Timeseries for all countries that have sufficient data for analysis (alphabetical order) showing raw data (grey), smoothed data (black), and decay interval exponential fits (red $r^2 \geq 0.8$, blue $r^2 < 0.8$). Top row cases/capita, middle row cases/tests, bottom row deaths/capita.

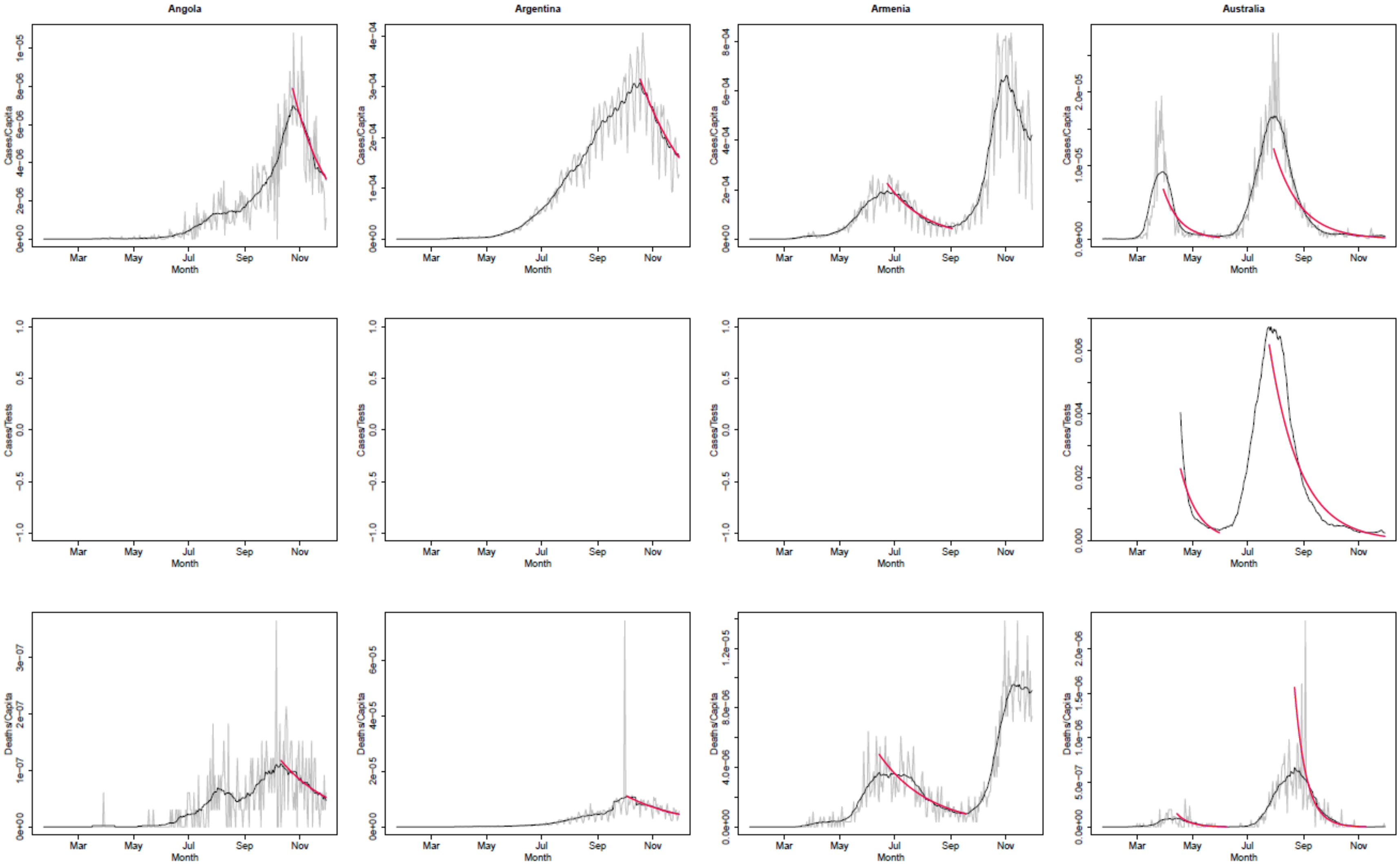
Angola
Argentina
Armenia
Australia
Cases/Capita
Cases/Tests
Deaths/Capita
Month
Mar
May
Jul
Sep
Nov

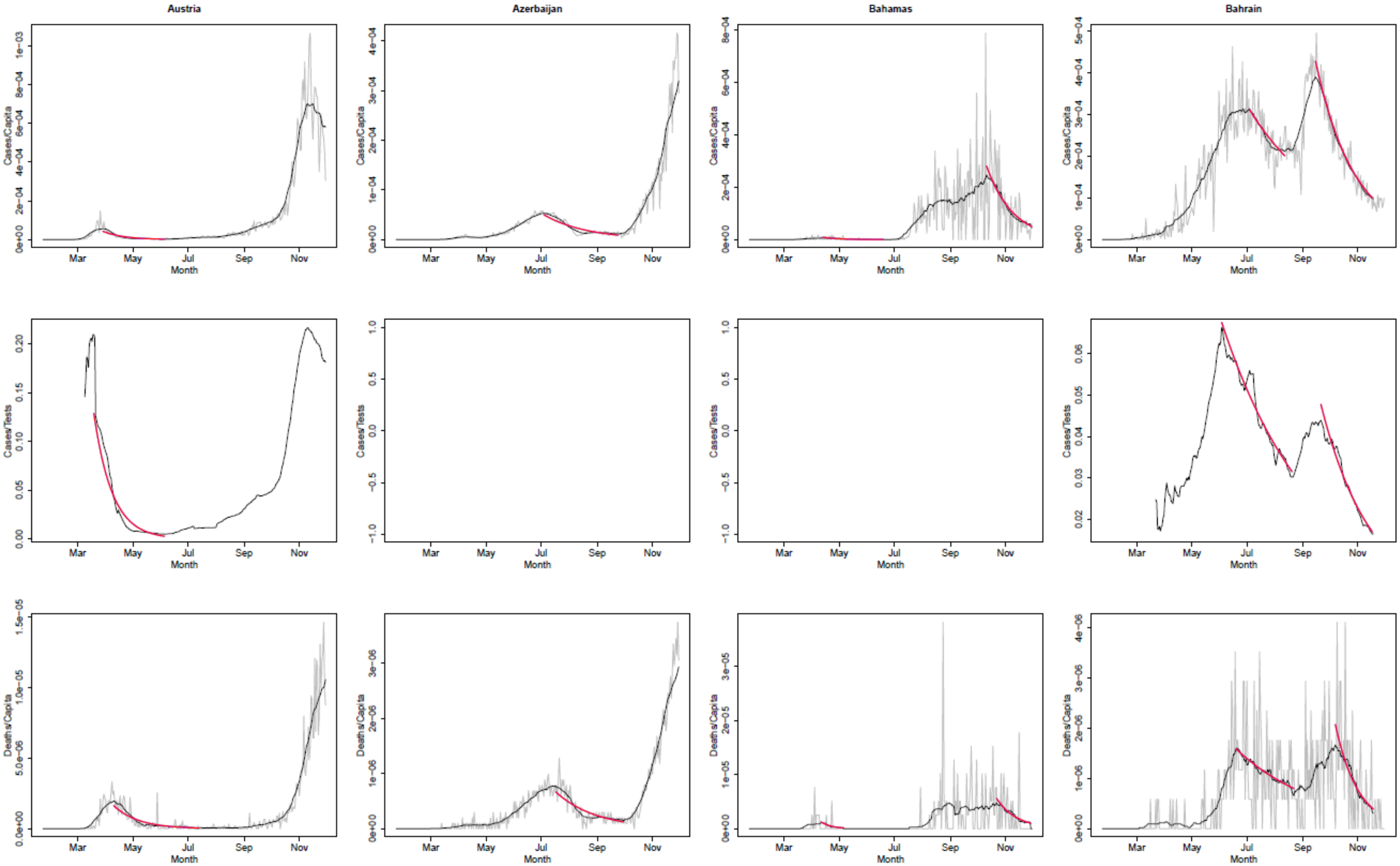
Austria
Azerbaijan
Bahamas
Bahrain
Cases/Capita
Cases/Tests
Deaths/Capita
Month
Mar
May
Jul
Sep
Nov

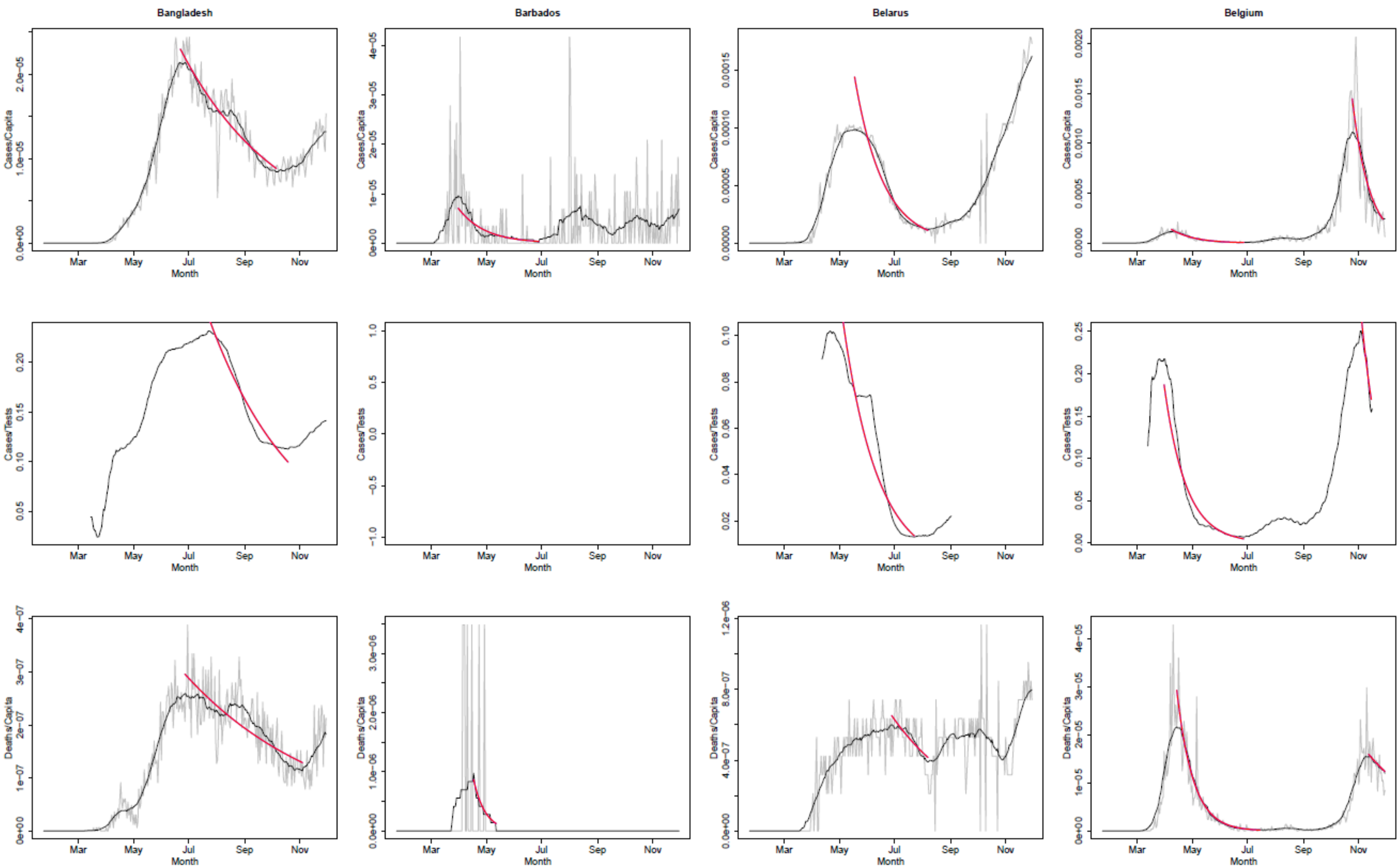
Bangladesh
Barbados
Belarus
Belgium
Cases/Capita
Cases/Tests
Deaths/Capita
Mar
May
Jul
Sep
Nov
Month

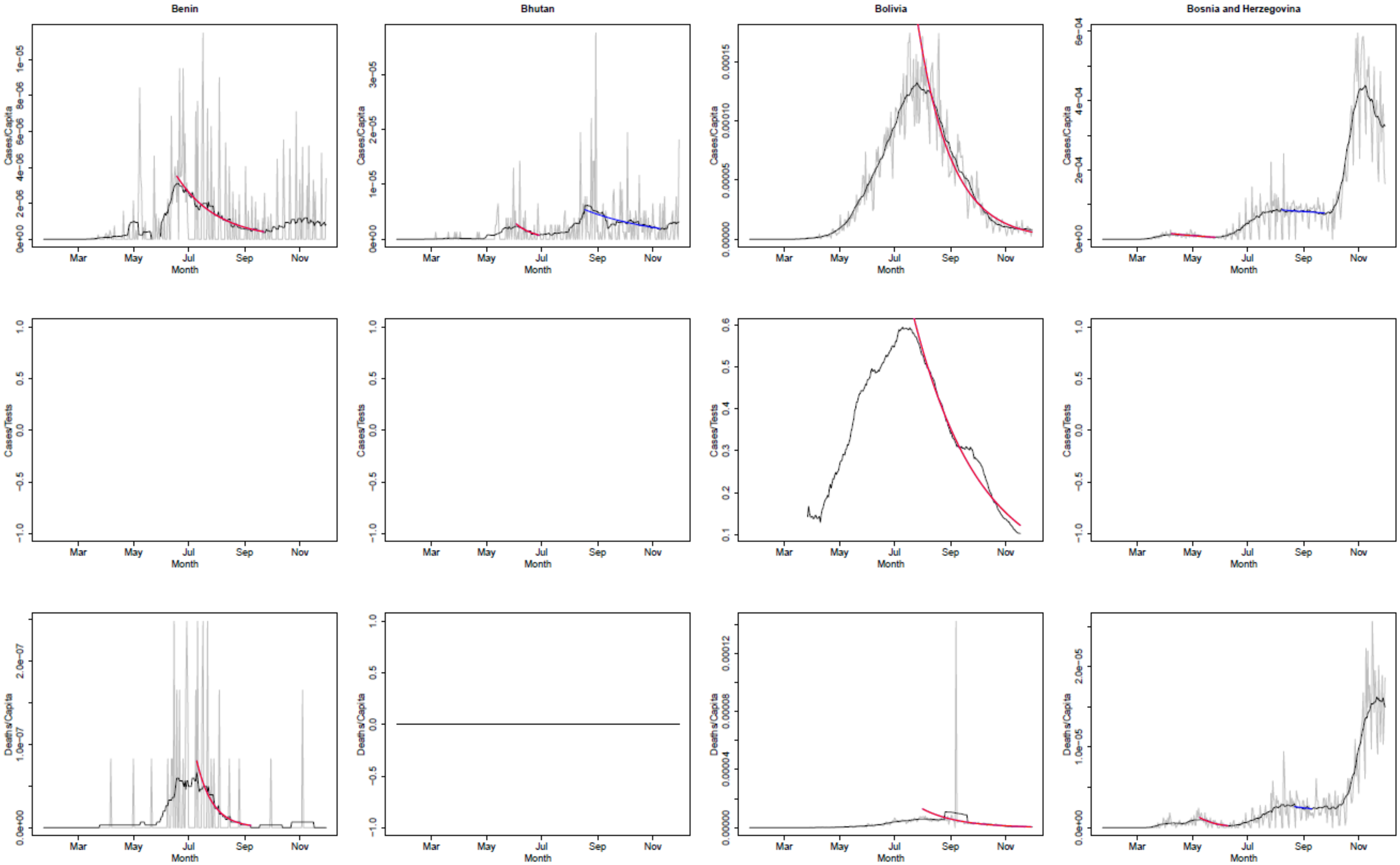

Benin
Bhutan
Bolivia
Bosnia and Herzegovina
Cases/Capita
Cases/Tests
Deaths/Capita
Month
Mar
May
Jul
Sep
Nov

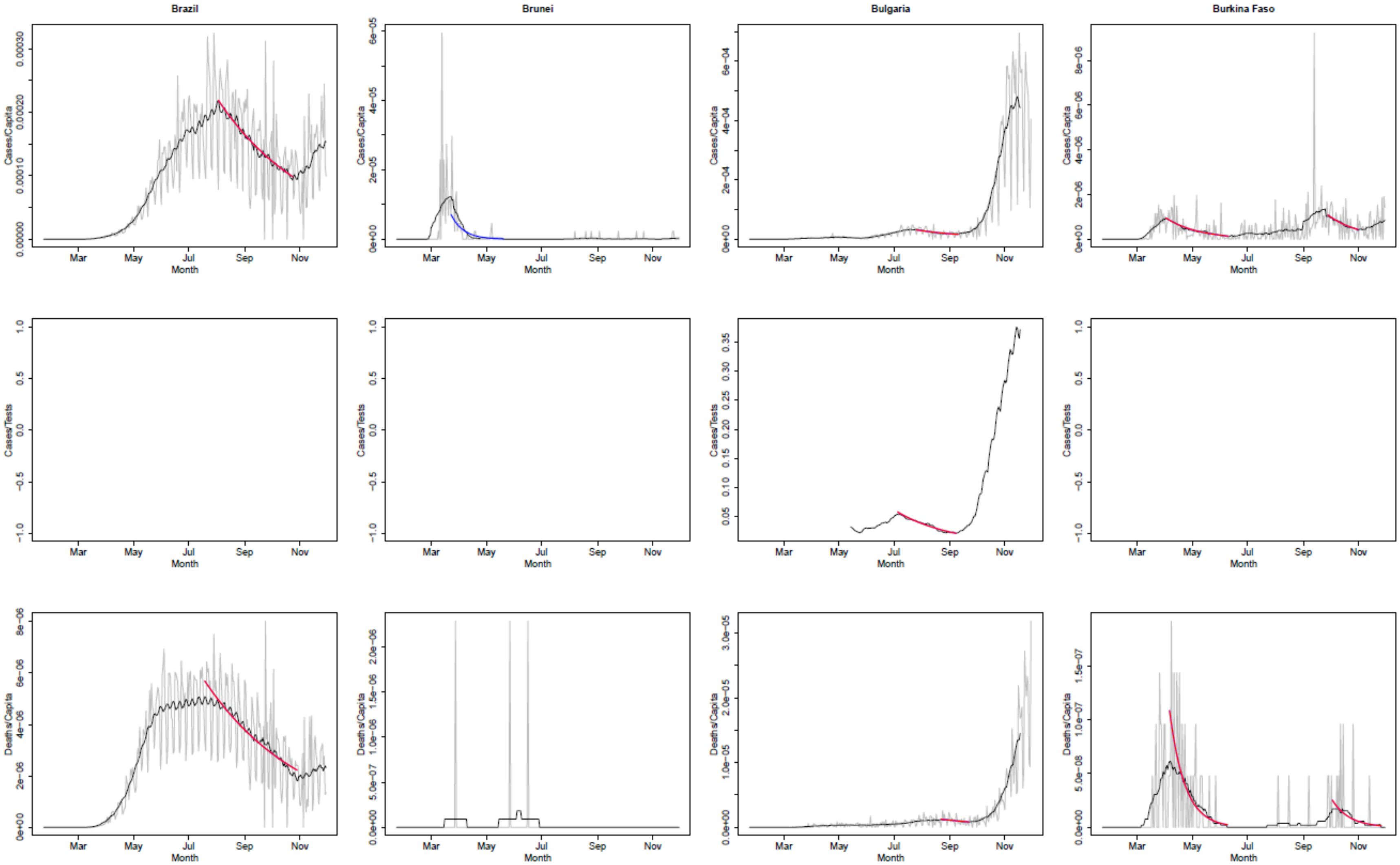
Brazil
Brunei
Bulgaria
Burkina Faso
Cases/Capita
Cases/Tests
Deaths/Capita
Month
Mar
May
Jul
Sep
Nov

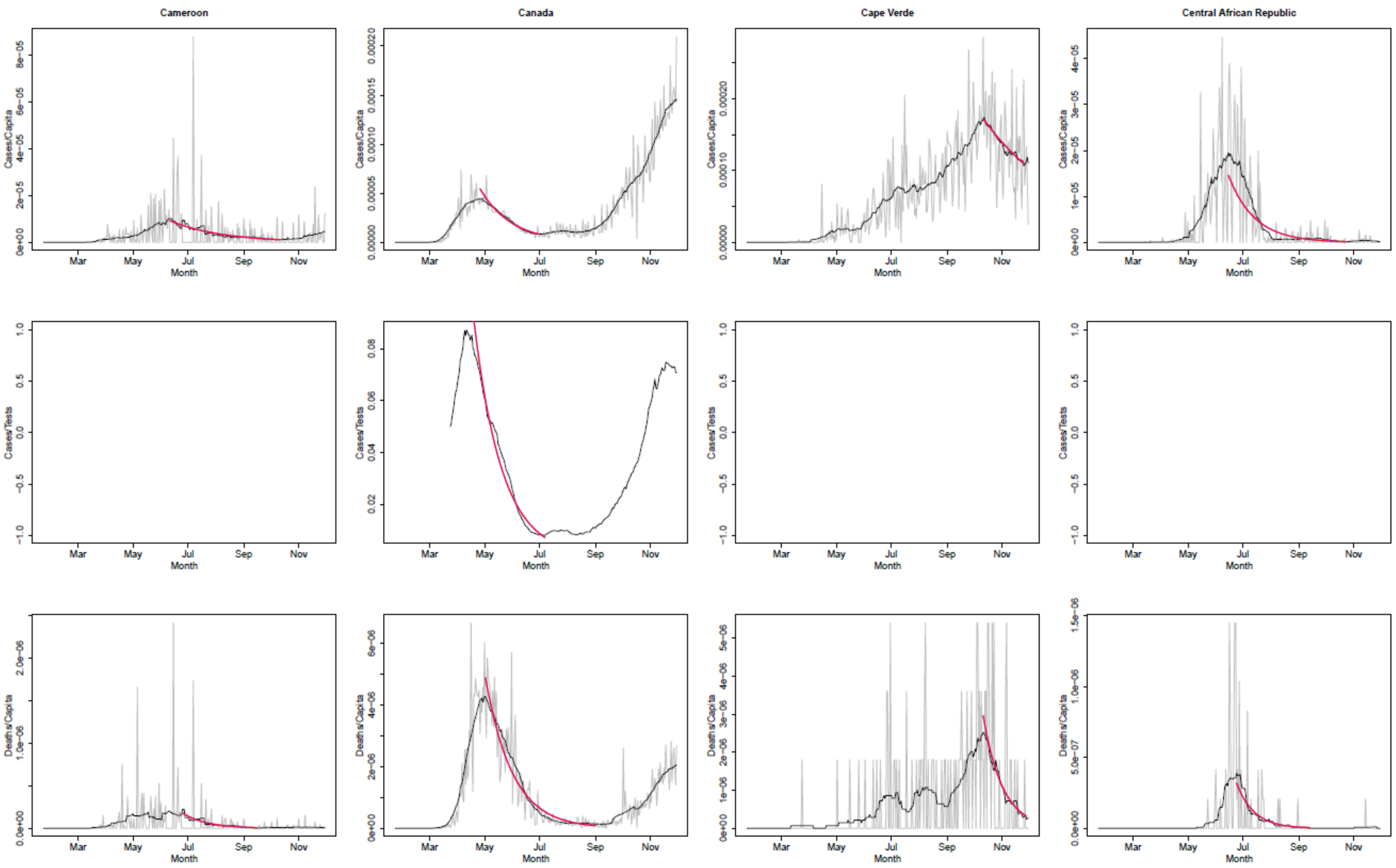
Cameroon
Canada
Cape Verde
Central African Republic
Cases/Capita
Cases/Tests
Deaths/Capita
Month
Mar
May
Jul
Sep
Nov

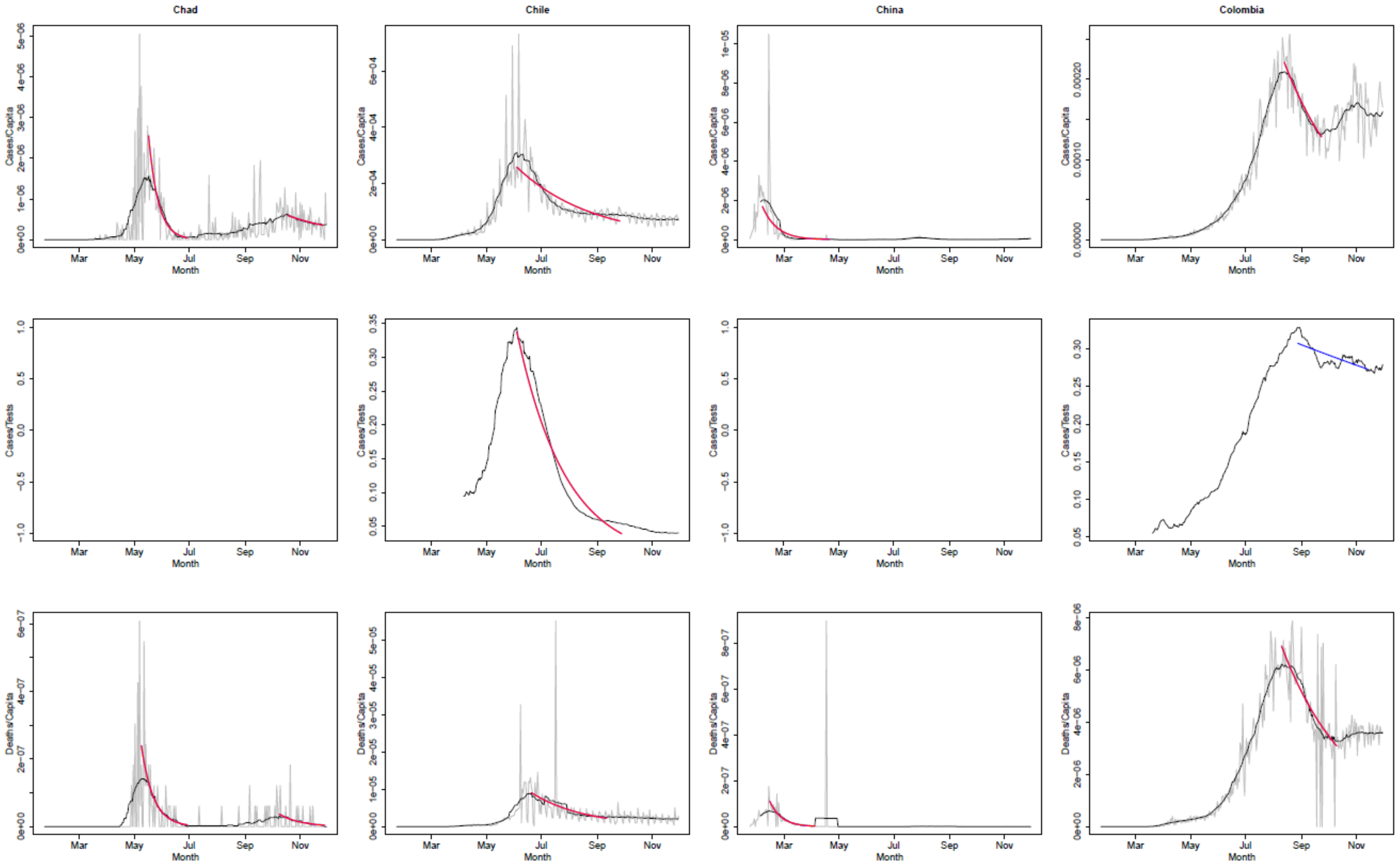
Chad
Chile
China
Colombia
Cases/Capita
Cases/Tests
Deaths/Capita
Month
Mar
May
Jul
Sep
Nov

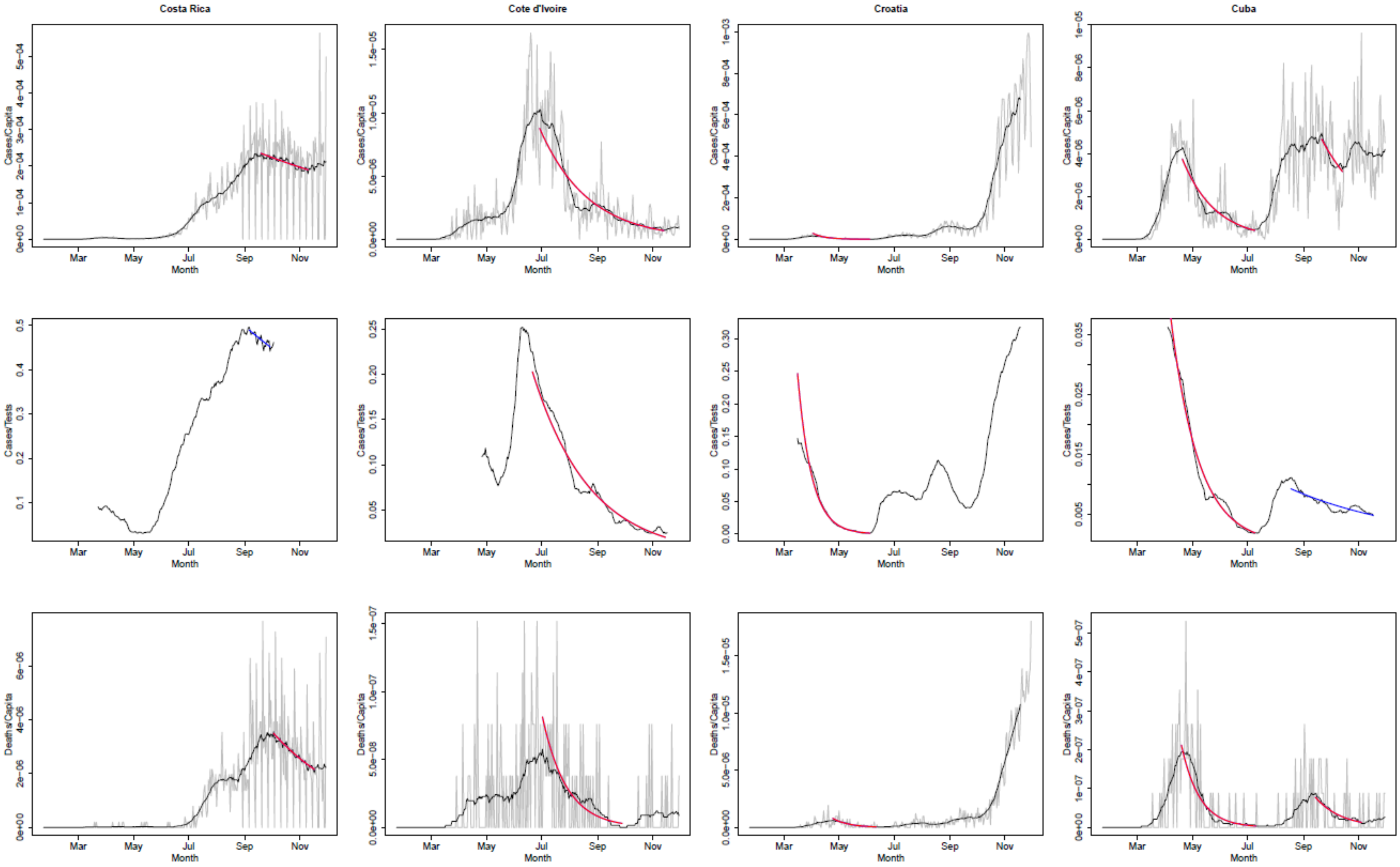
Costa Rica
Cote d'Ivoire
Croatia
Cuba
Cases/Capita
Cases/Tests
Deaths/Capita
Month
Mar
May
Jul
Sep
Nov

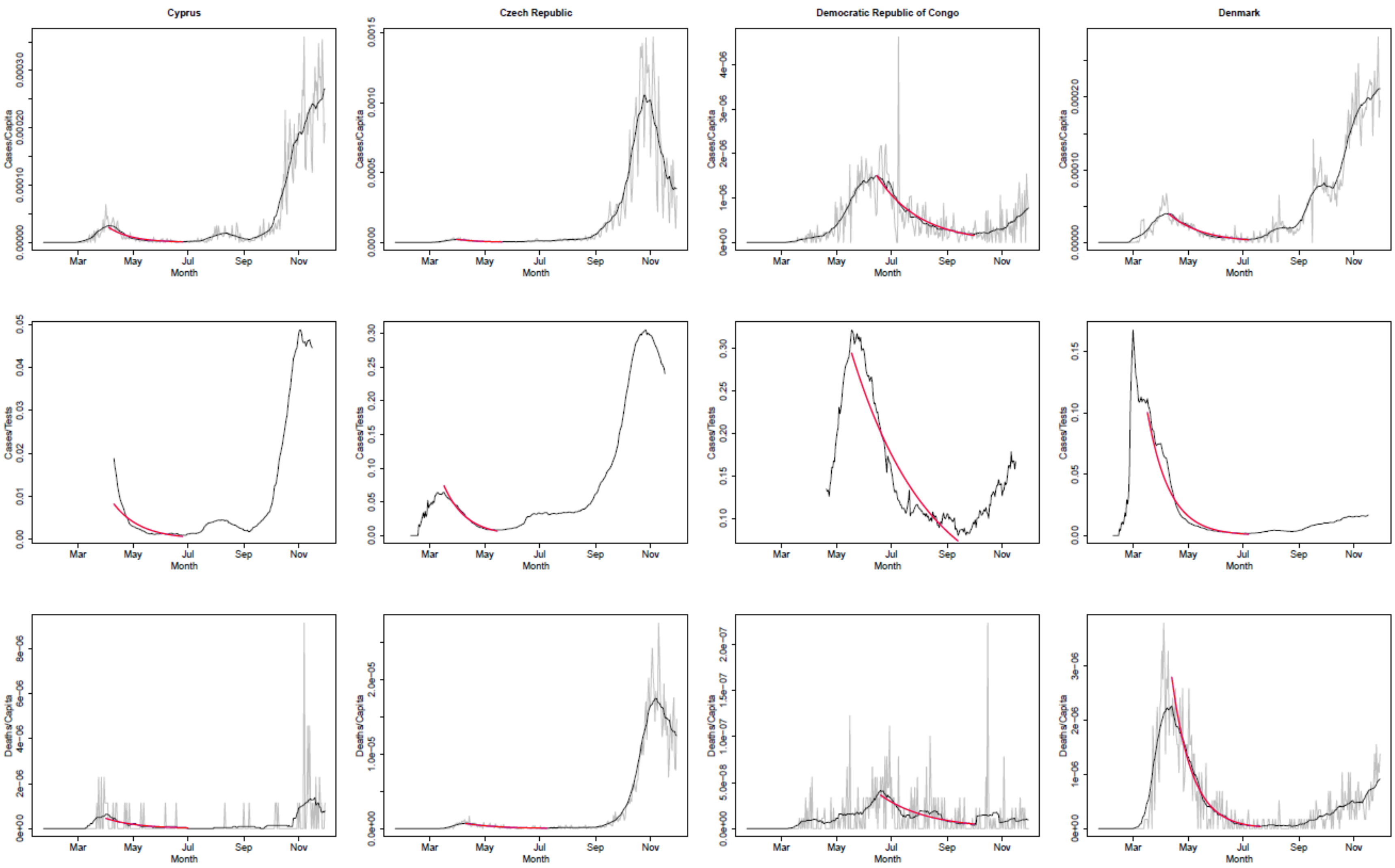
Cyprus
Czech Republic
Democratic Republic of Congo
Denmark
Cases/Capita
Cases/Tests
Deaths/Capita
Month
Mar
May
Jul
Sep
Nov

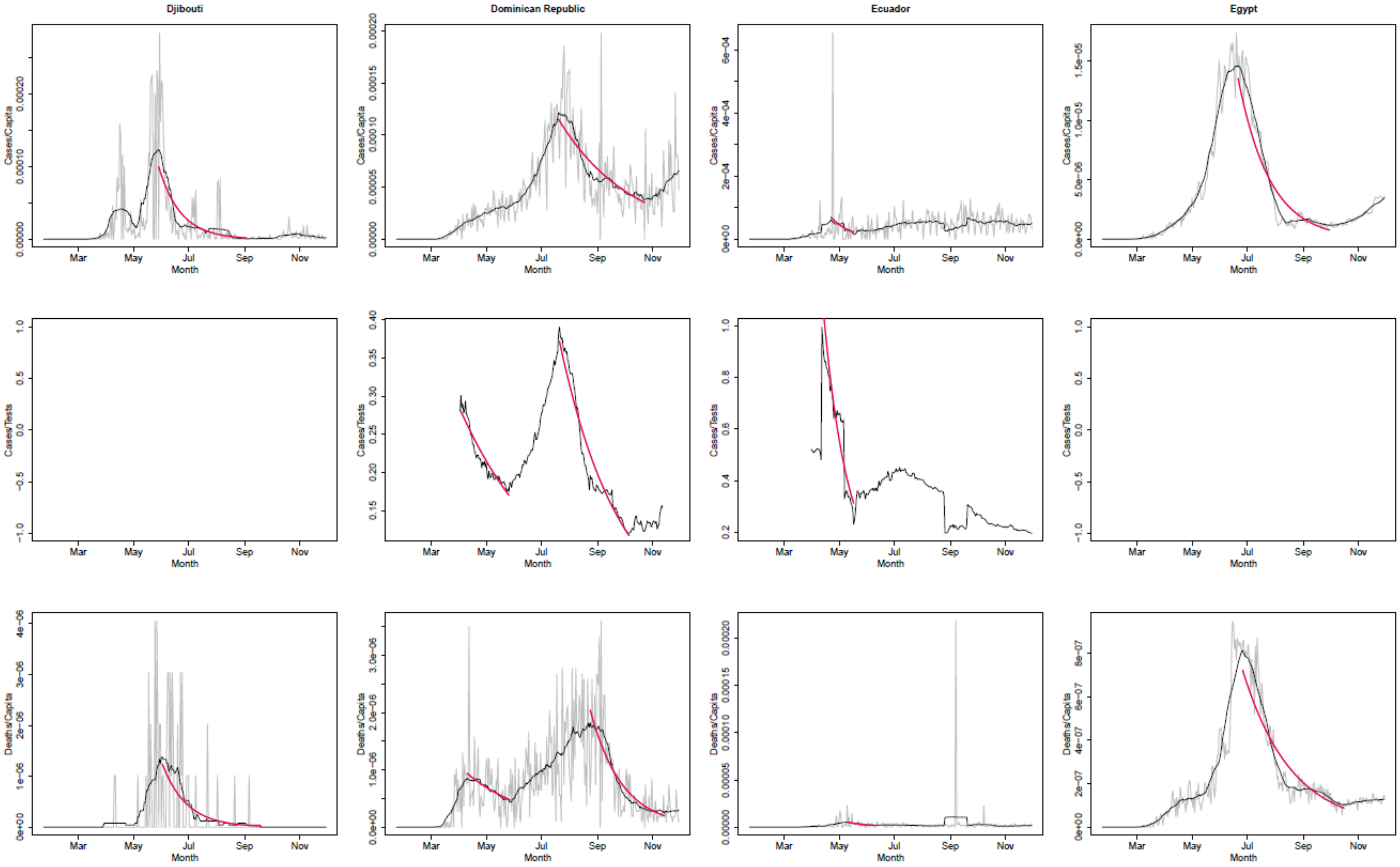

Djibouti
Dominican Republic
Ecuador
Egypt
Cases/Capita
Cases/Tests
Deaths/Capita
Month
Mar
May
Jul
Sep
Nov

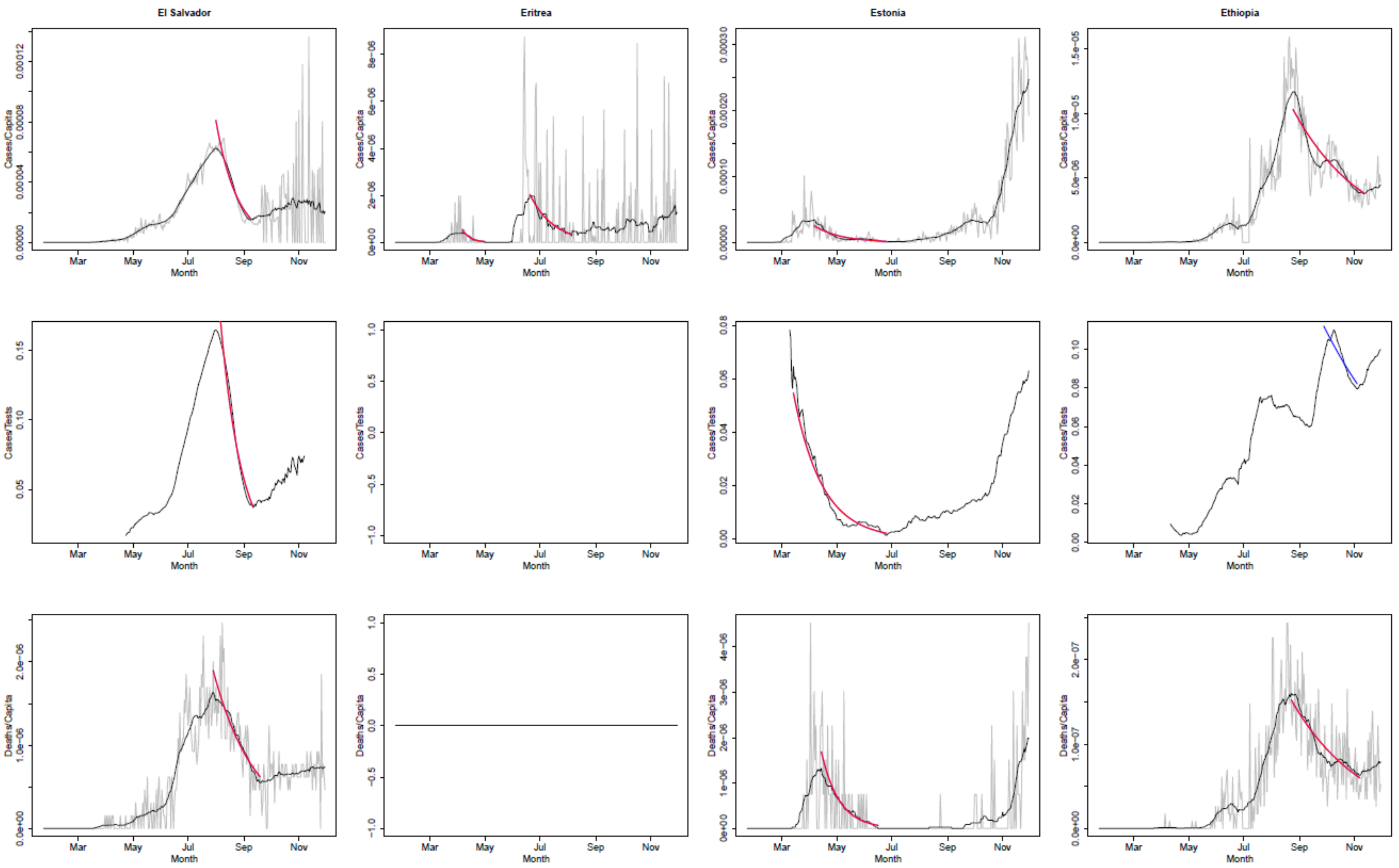
El Salvador
Eritrea
Estonia
Ethiopia
Cases/Capita
Cases/Tests
Deaths/Capita
Month
Mar
May
Jul
Sep
Nov

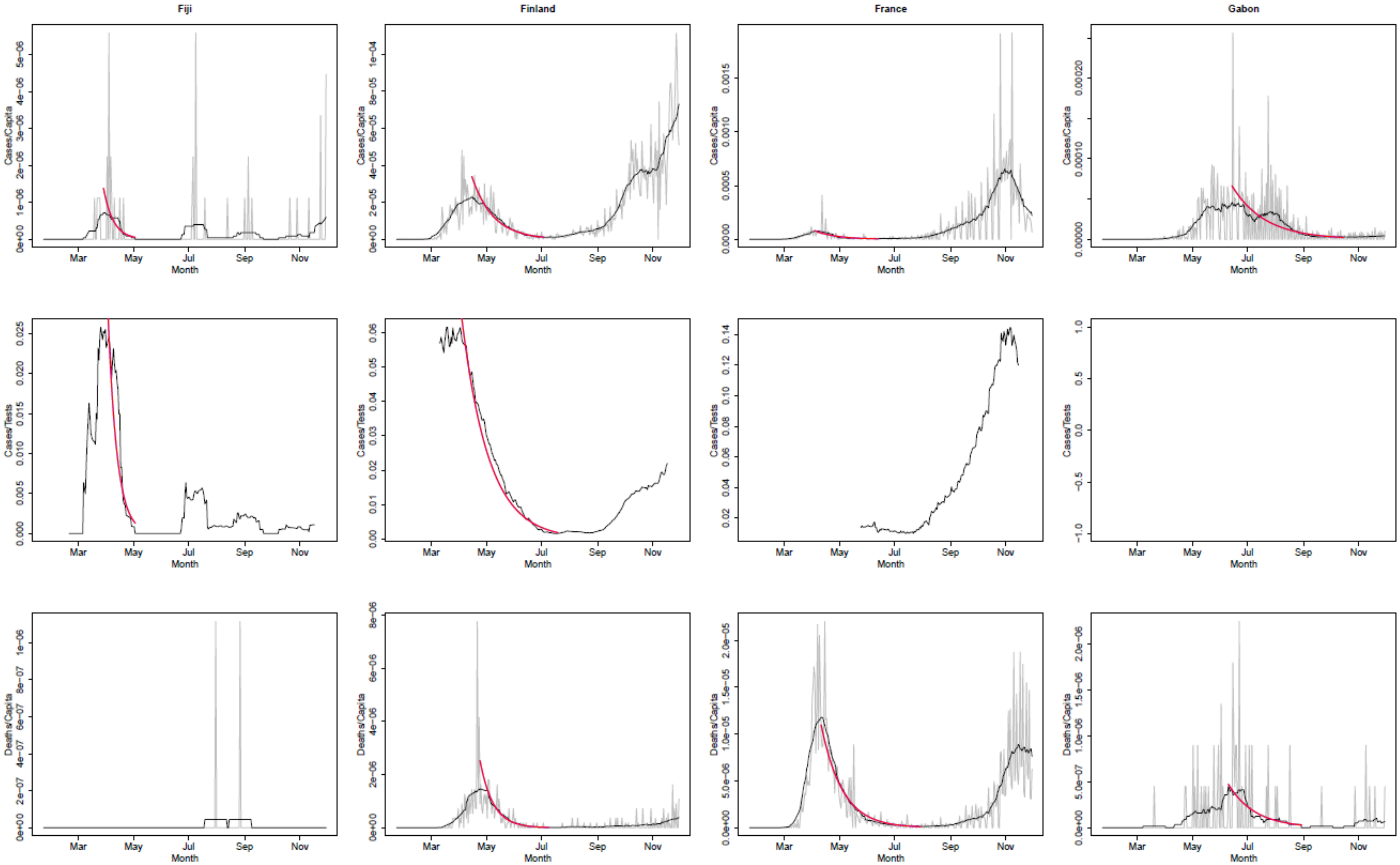
Fiji
Finland
France
Gabon
Cases/Capita
Cases/Tests
Deaths/Capita
Month
Mar
May
Jul
Sep
Nov

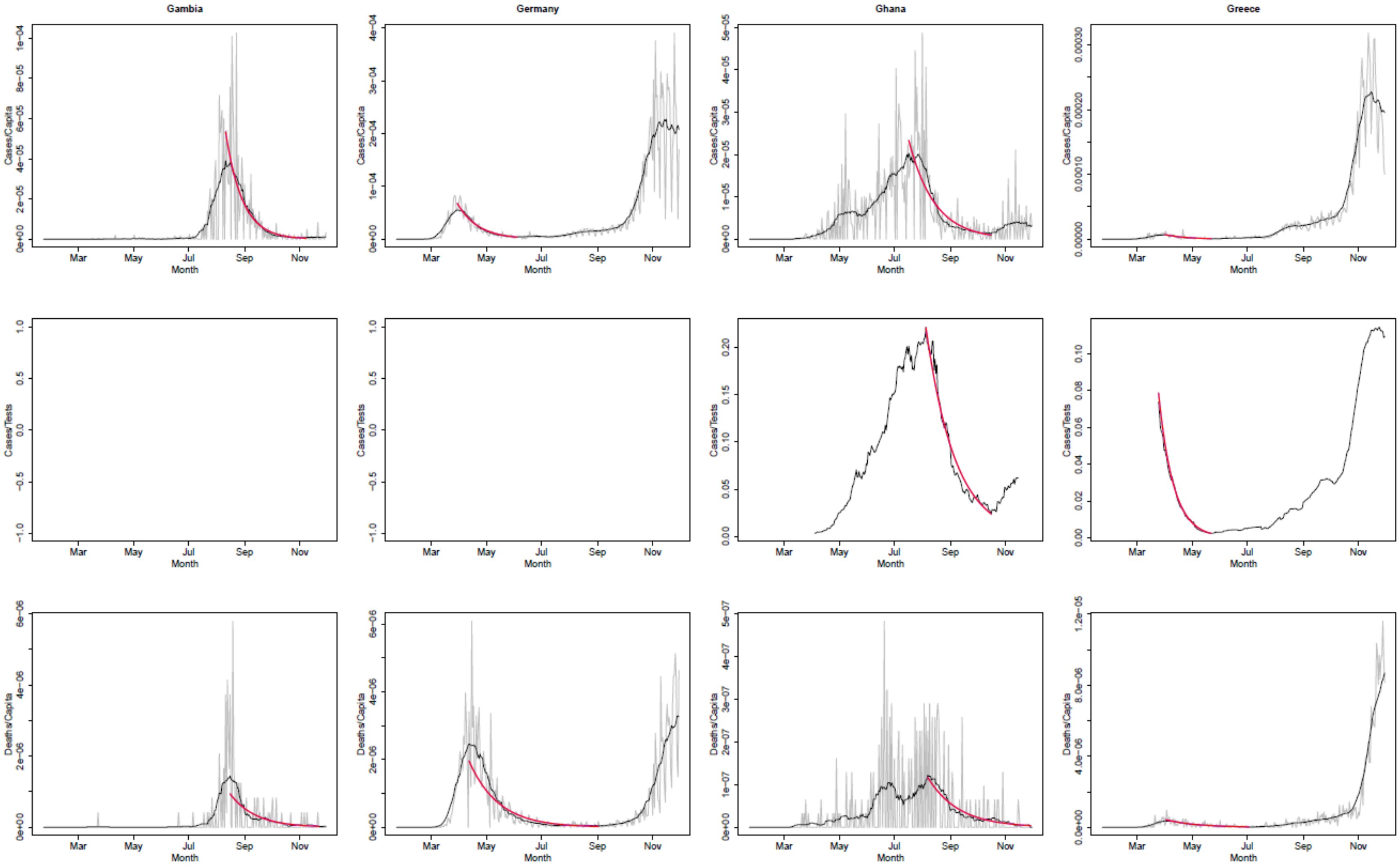
Gambia
Germany
Ghana
Greece
Cases/Capita
Cases/Tests
Deaths/Capita
Month
Mar
May
Jul
Sep
Nov

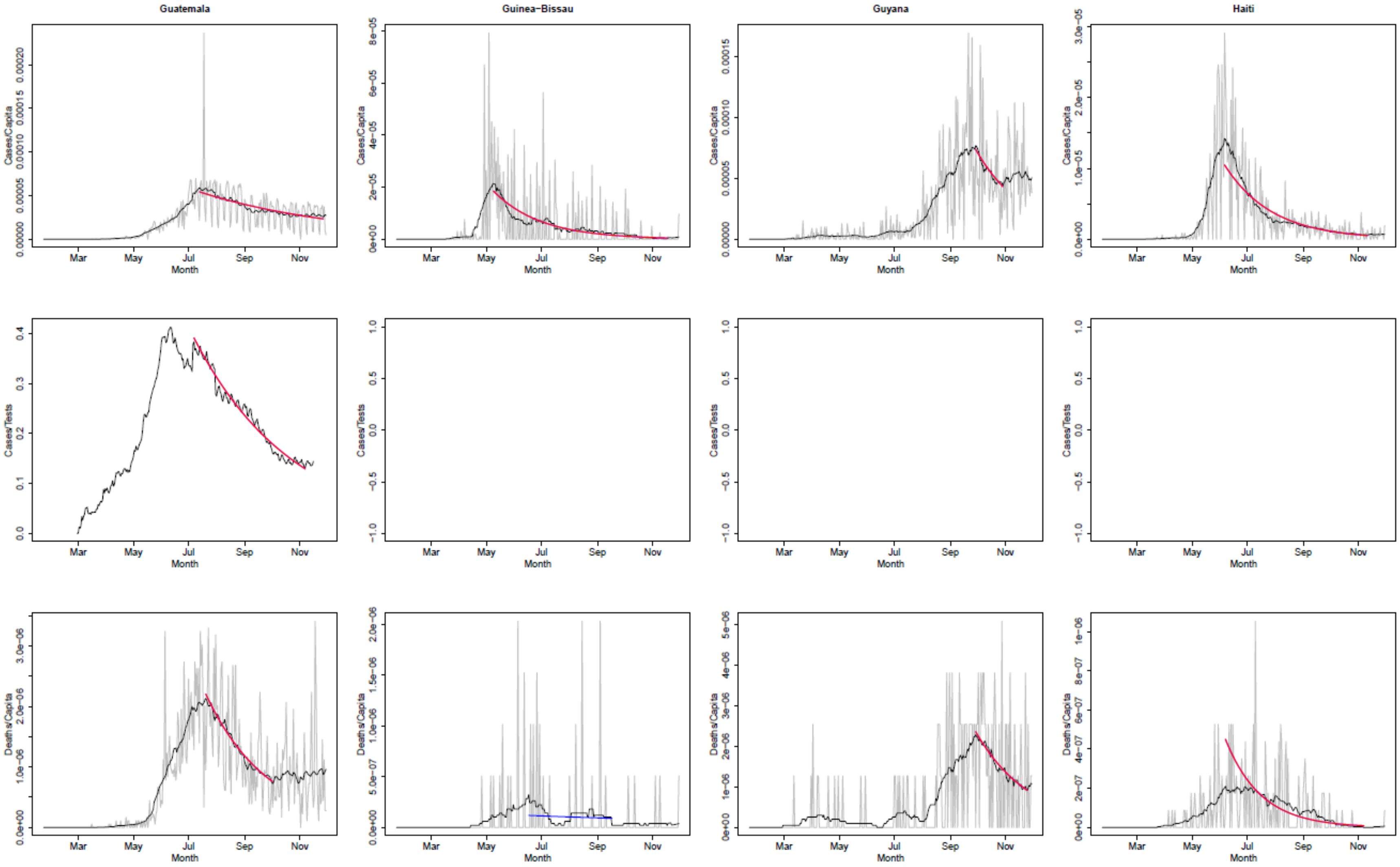
Guatemala
Guinea−Bissau
Guyana
Haiti
Cases/Capita
Cases/Tests
Deaths/Capita
Month
Mar
May
Jul
Sep
Nov

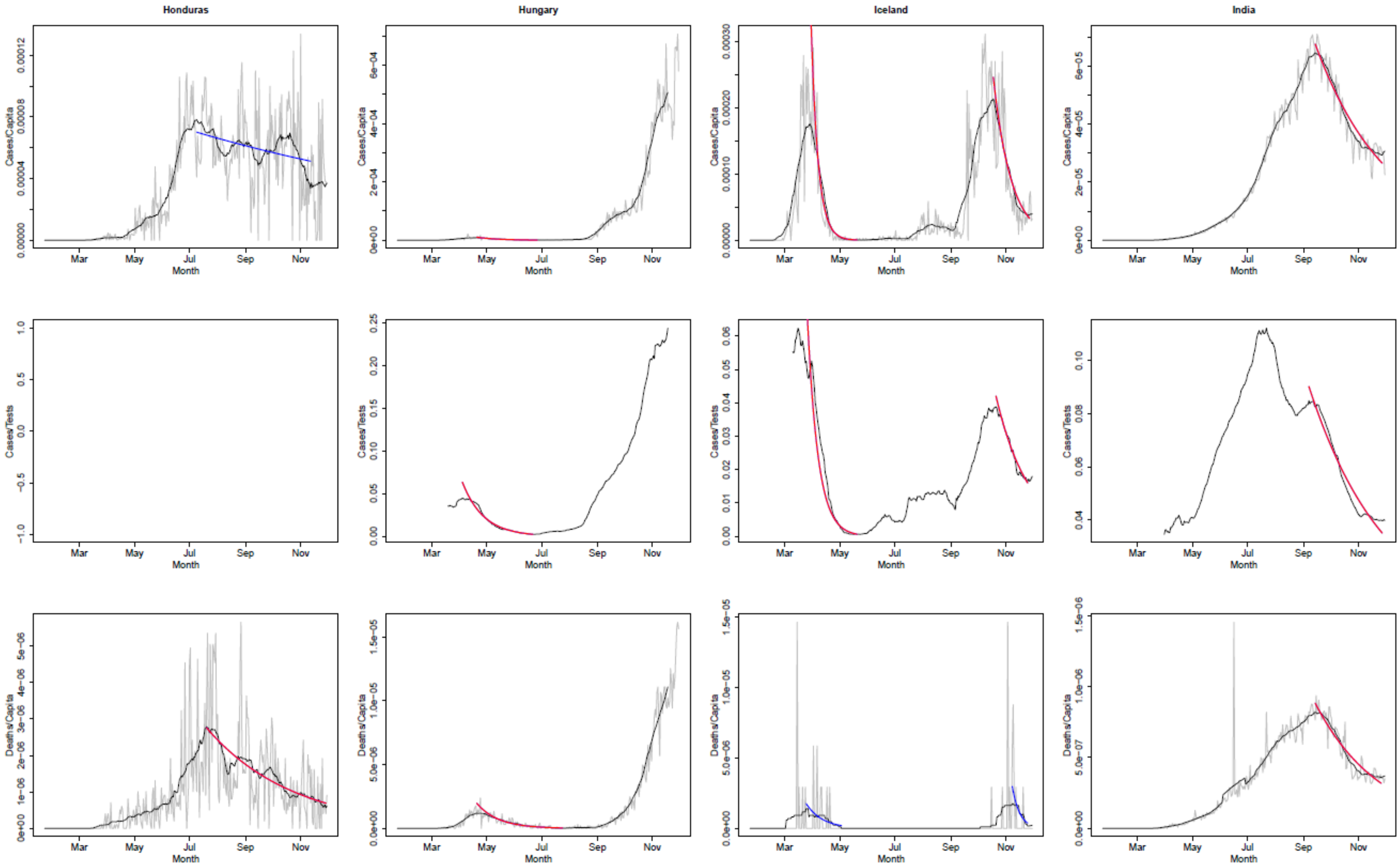

Honduras
Hungary
Iceland
India
Cases/Capita
Cases/Tests
Deaths/Capita
Mar
May
Jul
Sep
Nov
Month

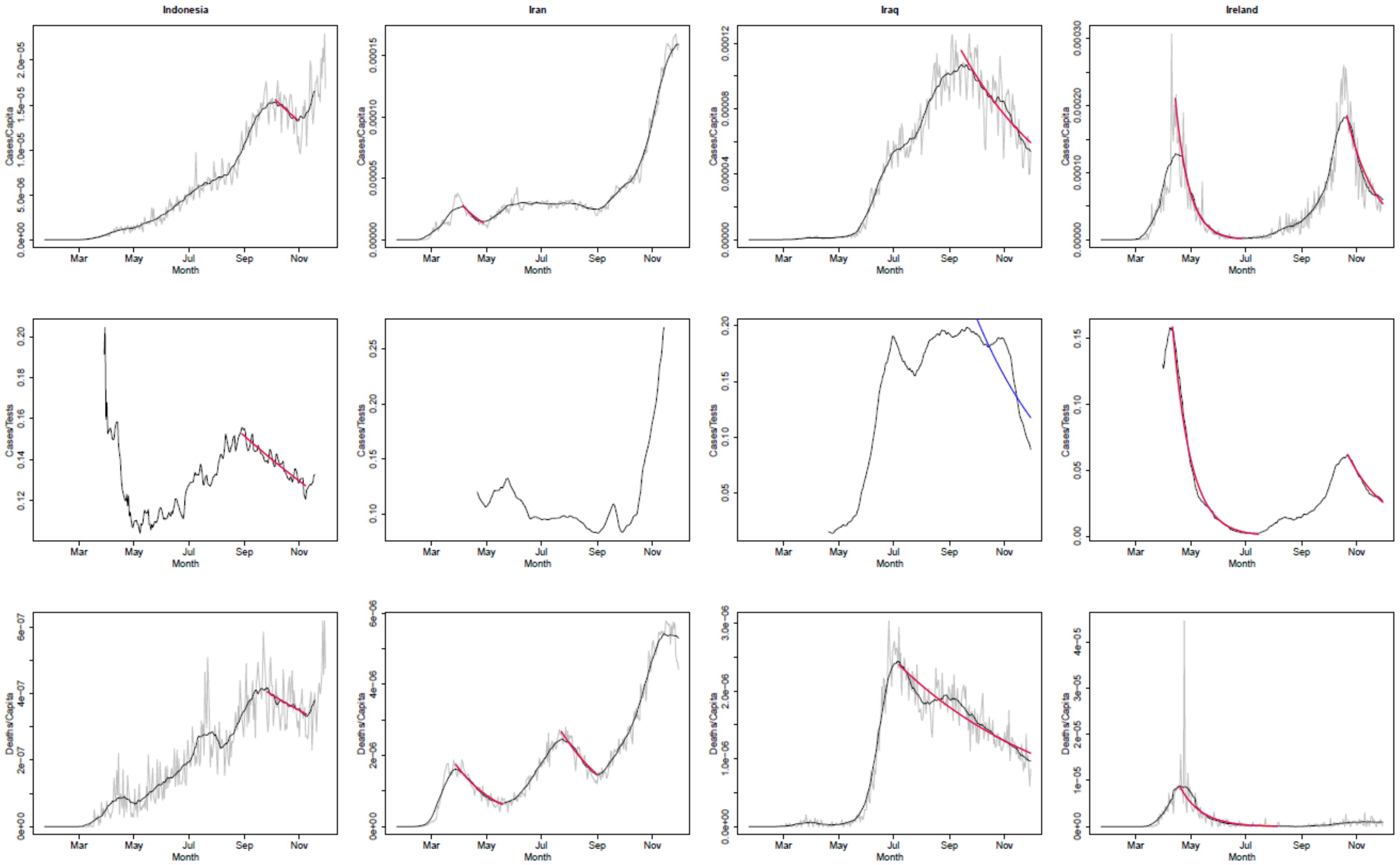
Indonesia
Iran
Iraq
Ireland
Cases/Capita
Cases/Tests
Deaths/Capita
Month
Mar
May
Jul
Sep
Nov

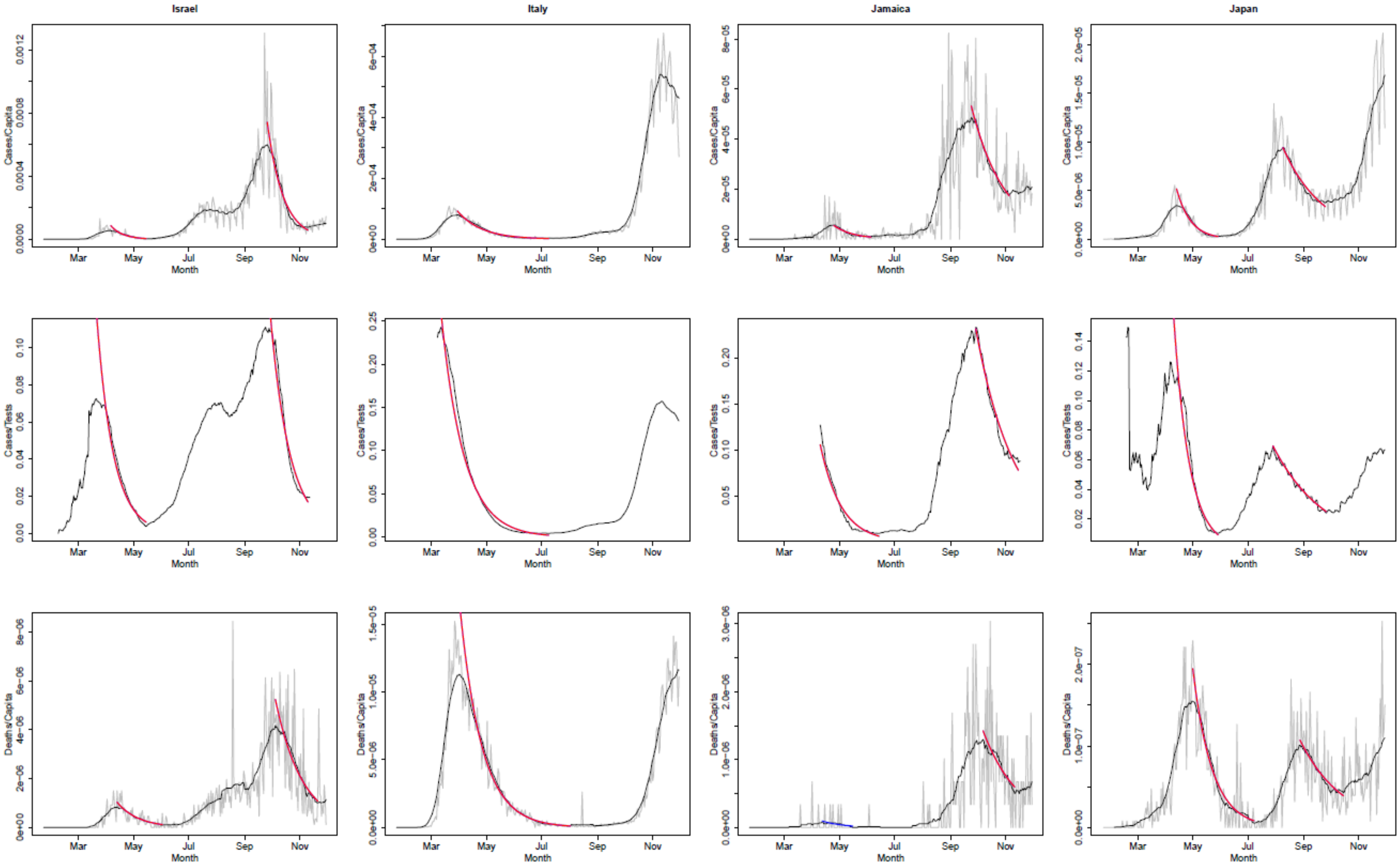

Israel
Italy
Jamaica
Japan
Cases/Capita
Cases/Tests
Deaths/Capita
Month
Mar
May
Jul
Sep
Nov

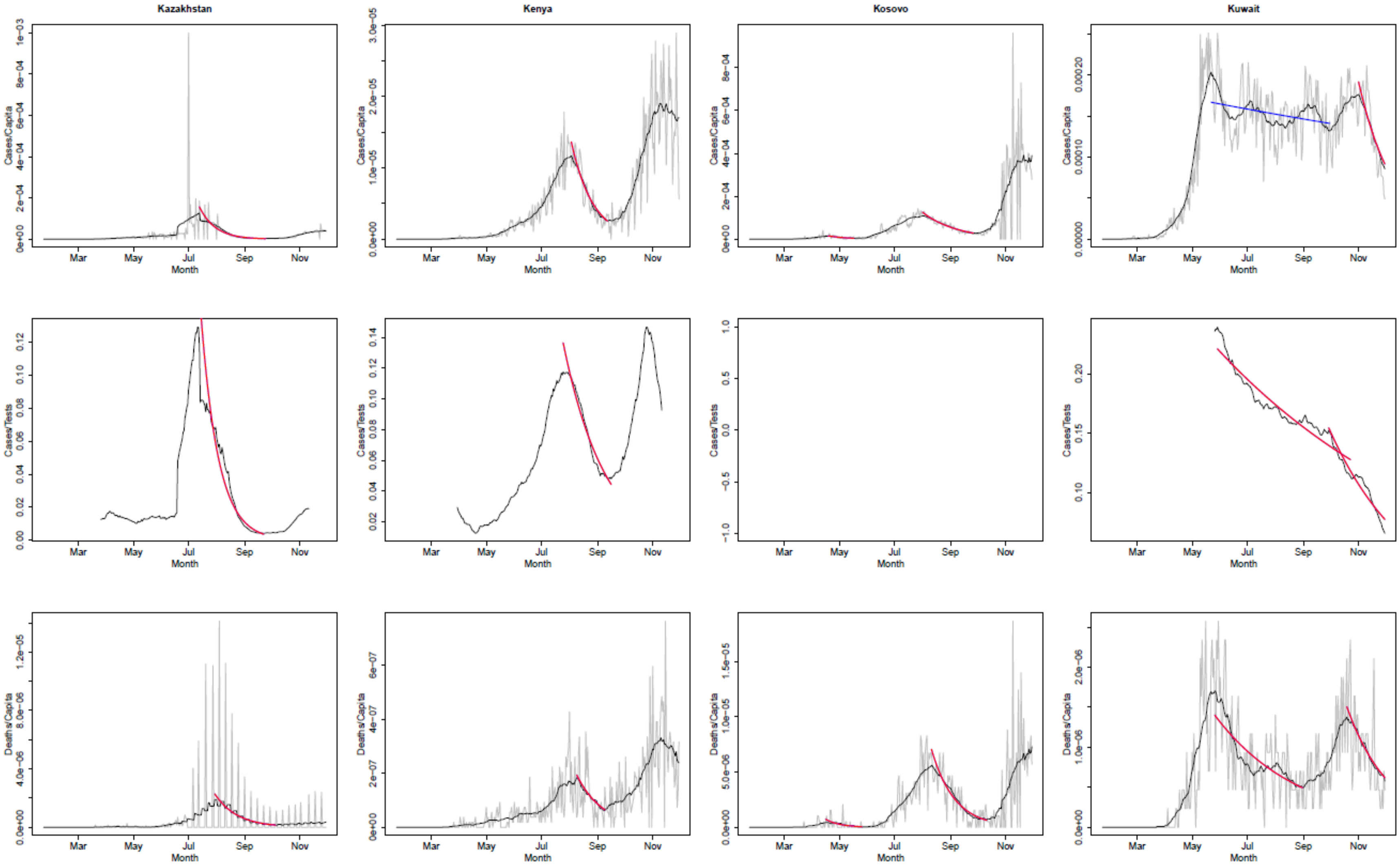

Kazakhstan
Kenya
Kosovo
Kuwait
Cases/Capita
Cases/Tests
Deaths/Capita
Month
Mar
May
Jul
Sep
Nov

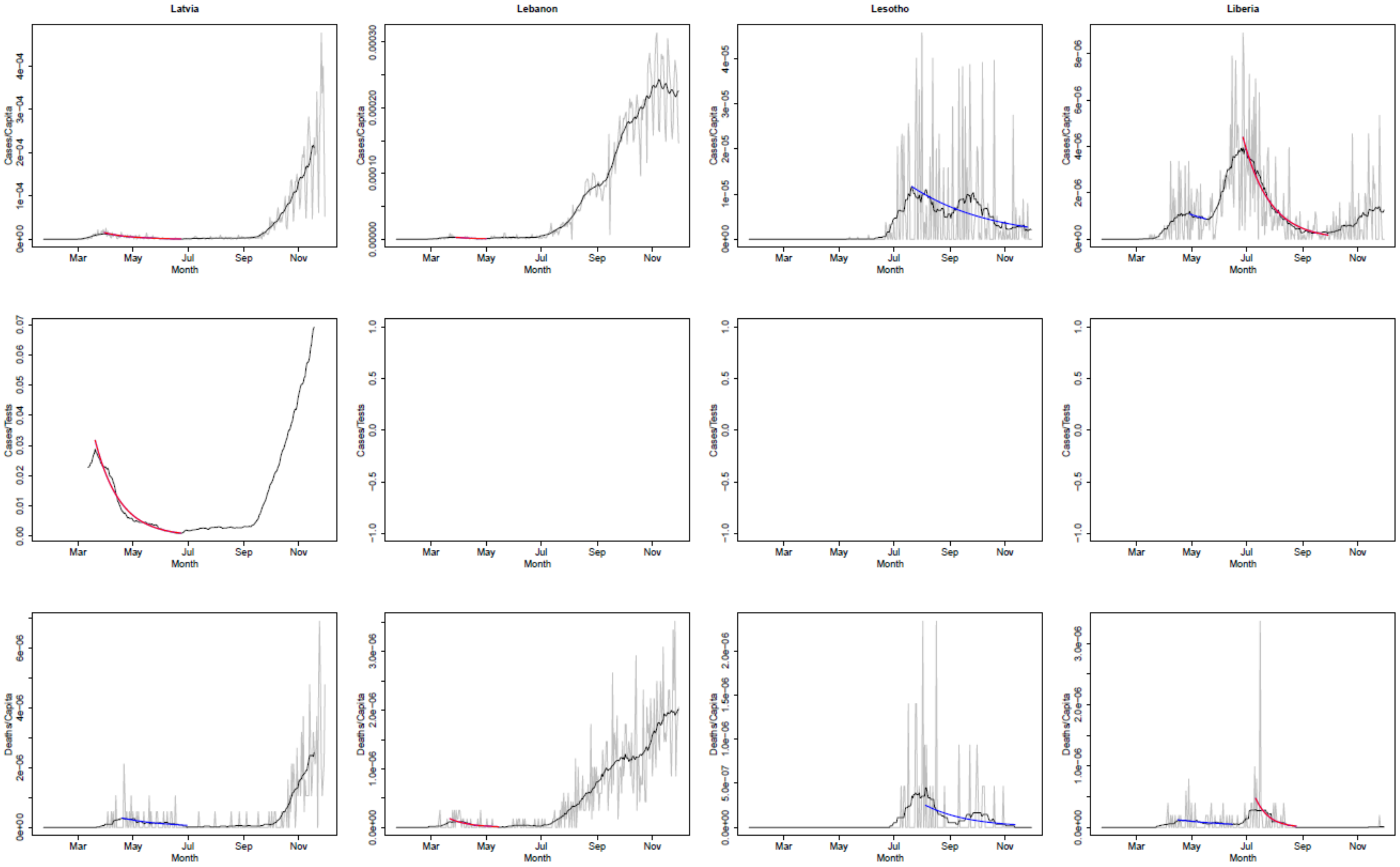
Latvia
Lebanon
Lesotho
Liberia
Cases/Capita
Cases/Tests
Deaths/Capita
Month
Mar
May
Jul
Sep
Nov

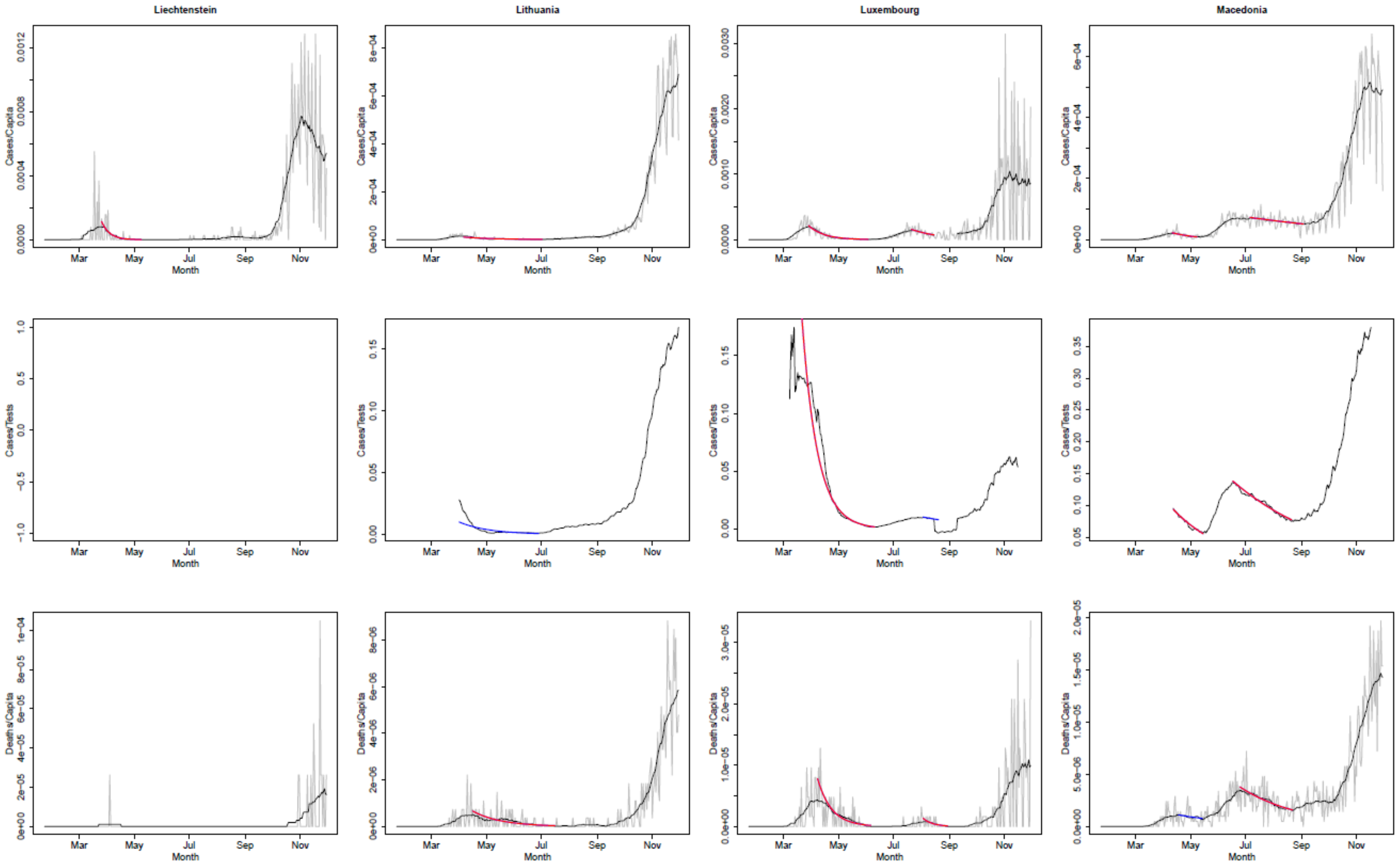
Liechtenstein
Lithuania
Luxembourg
Macedonia
Cases/Capita
Cases/Tests
Deaths/Capita
Mar
May
Jul
Sep
Nov
Month

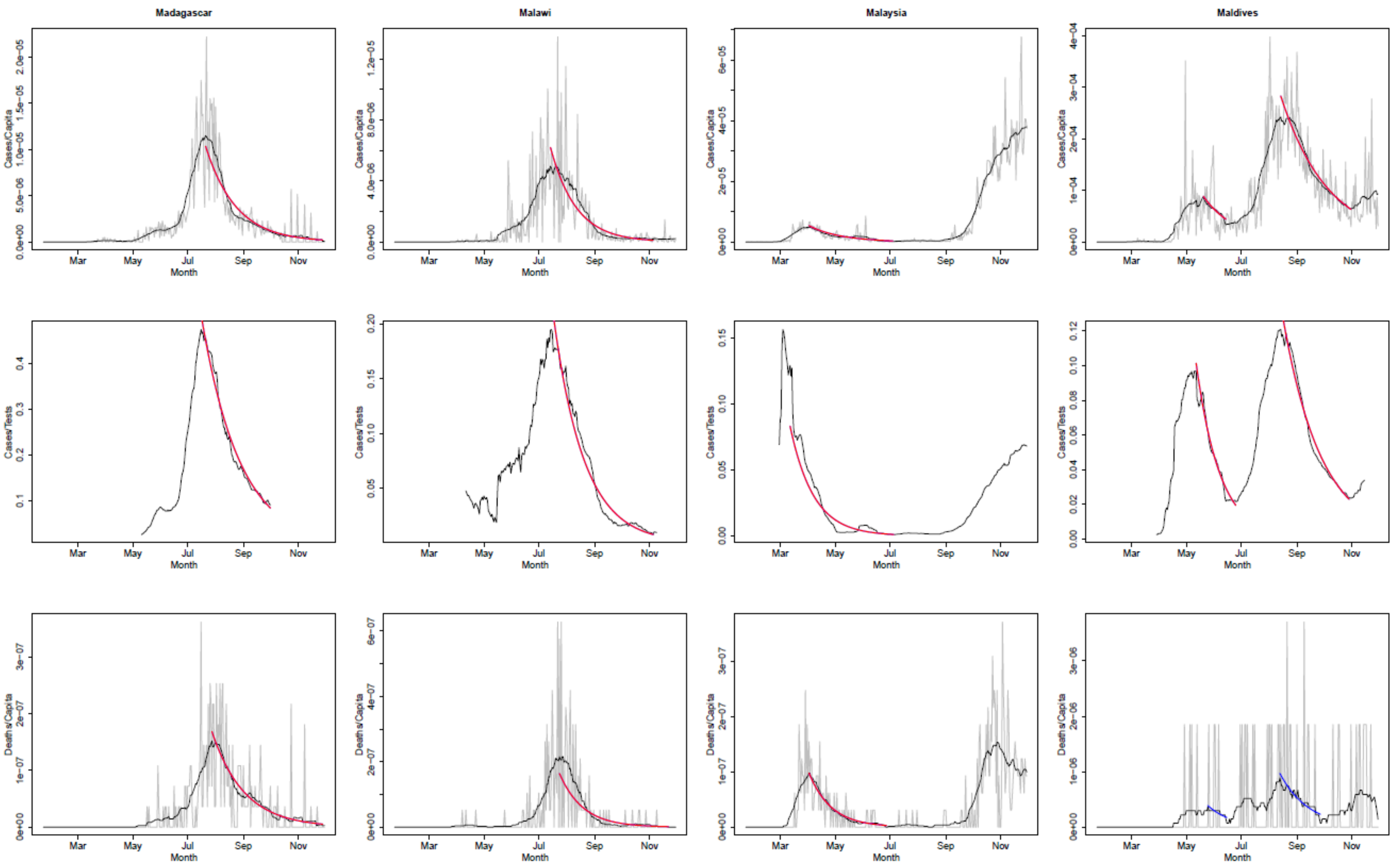
Madagascar
Malawi
Malaysia
Maldives
Cases/Capita
Cases/Tests
Deaths/Capita
Month
Mar
May
Jul
Sep
Nov

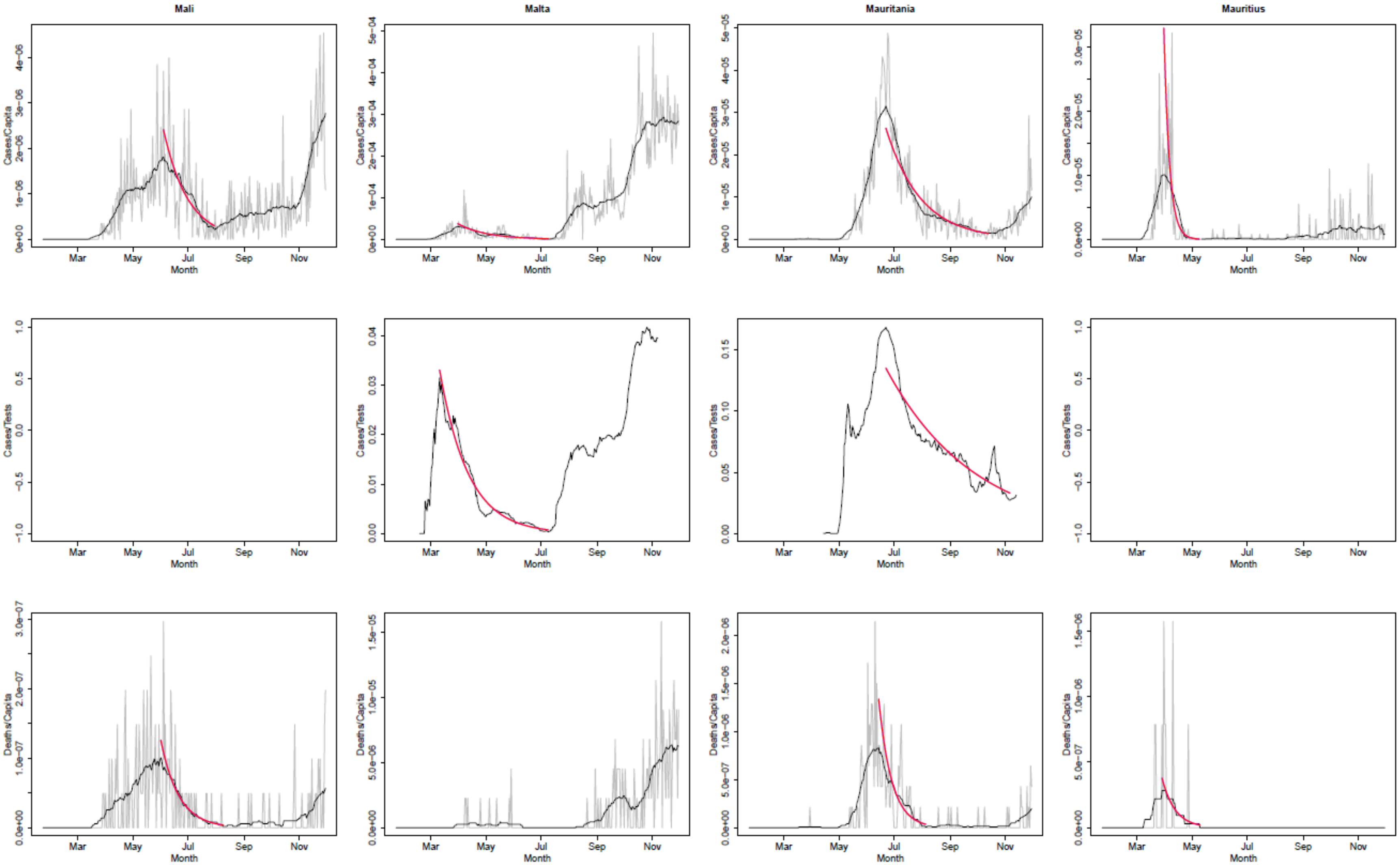

Mali
Malta
Mauritania
Mauritius
Cases/Capita
Cases/Tests
Deaths/Capita
Mar
May
Jul
Sep
Nov
Month

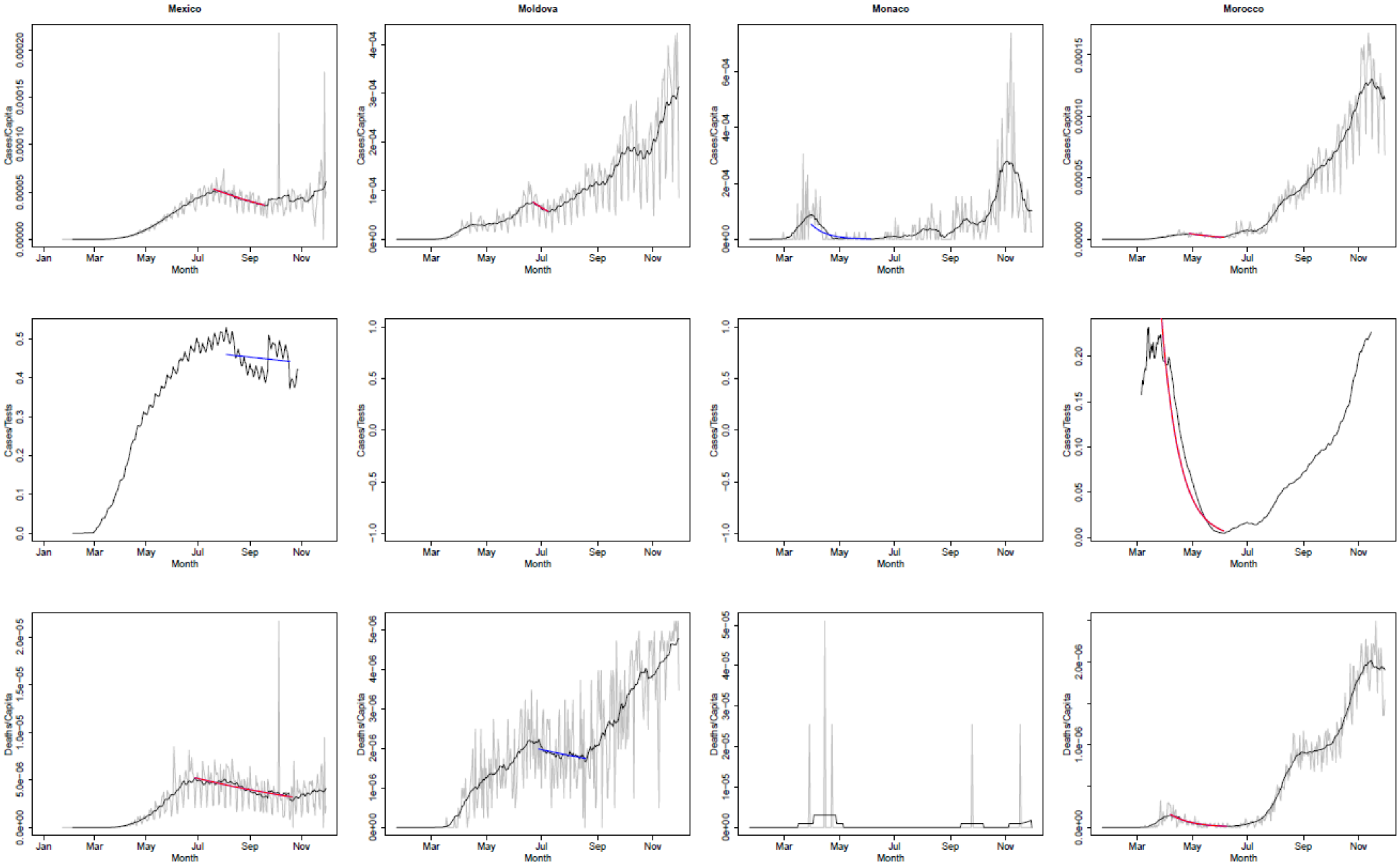
Mexico
Moldova
Monaco
Morocco
Cases/Capita
Cases/Tests
Deaths/Capita
Month
Jan
Mar
May
Jul
Sep
Nov

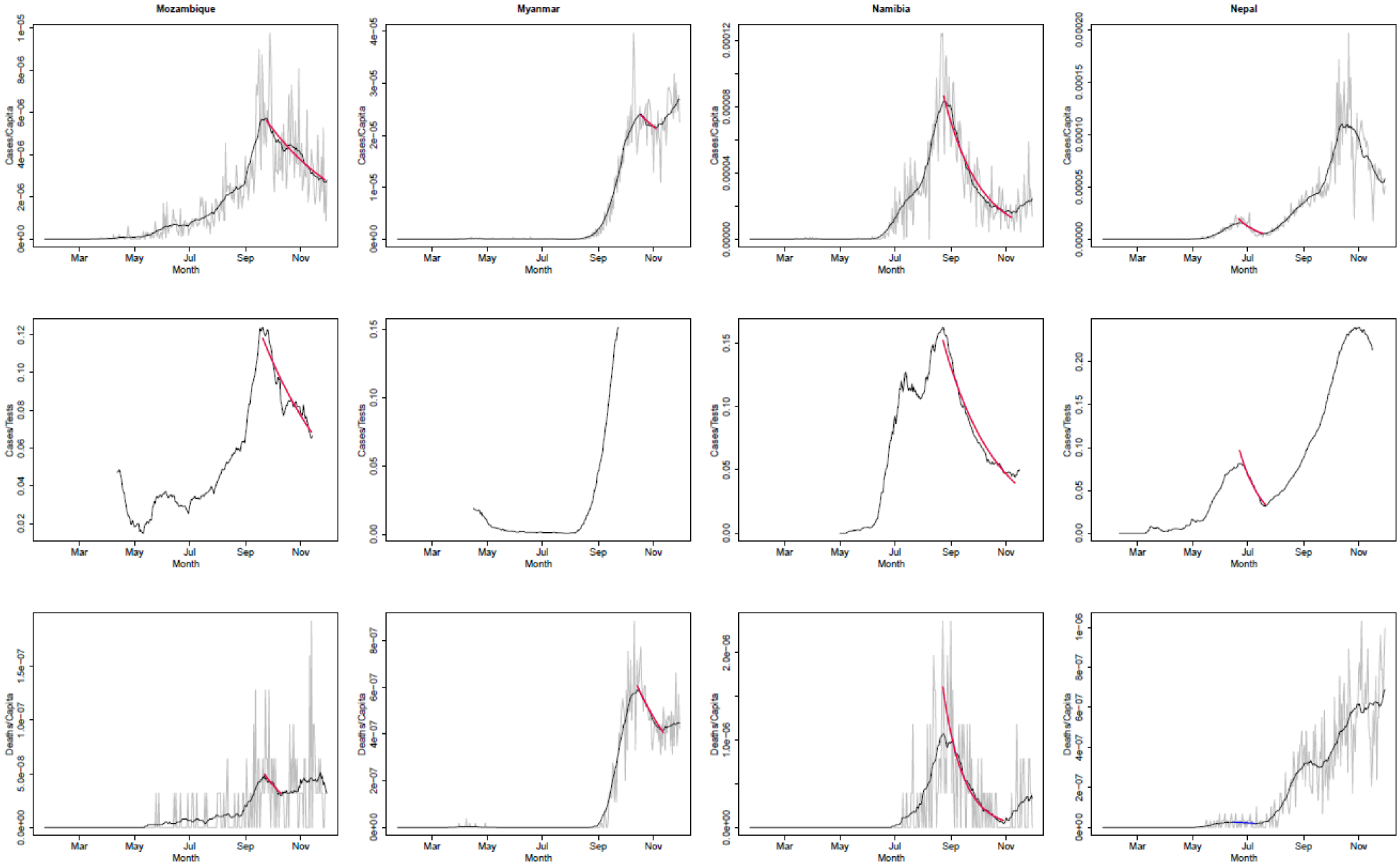
Mozambique
Myanmar
Namibia
Nepal
Cases/Capita
Cases/Tests
Deaths/Capita
Month
Mar
May
Jul
Sep
Nov

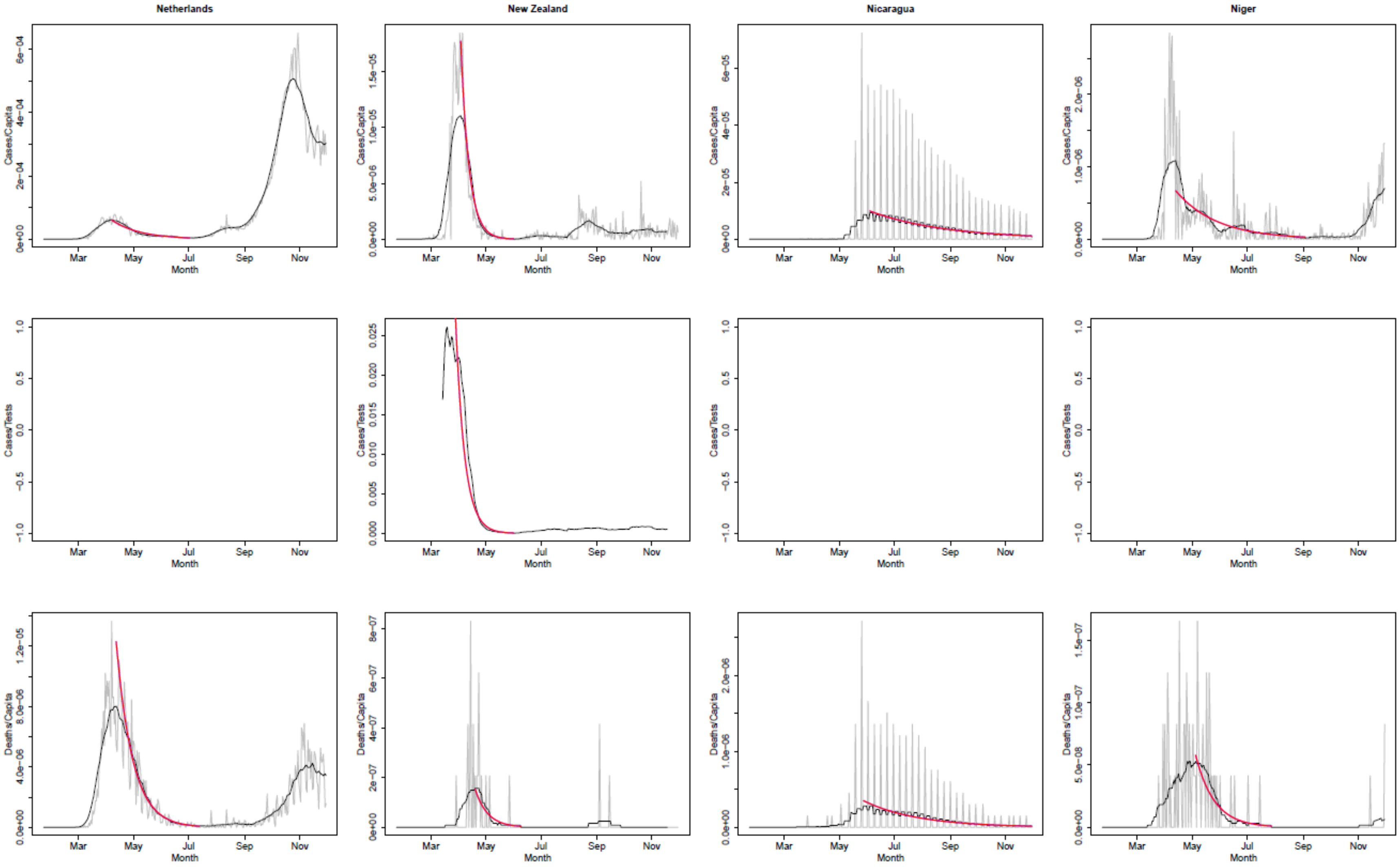
Netherlands
New Zealand
Nicaragua
Niger
Cases/Capita
Cases/Tests
Deaths/Capita
Month
Mar
May
Jul
Sep
Nov

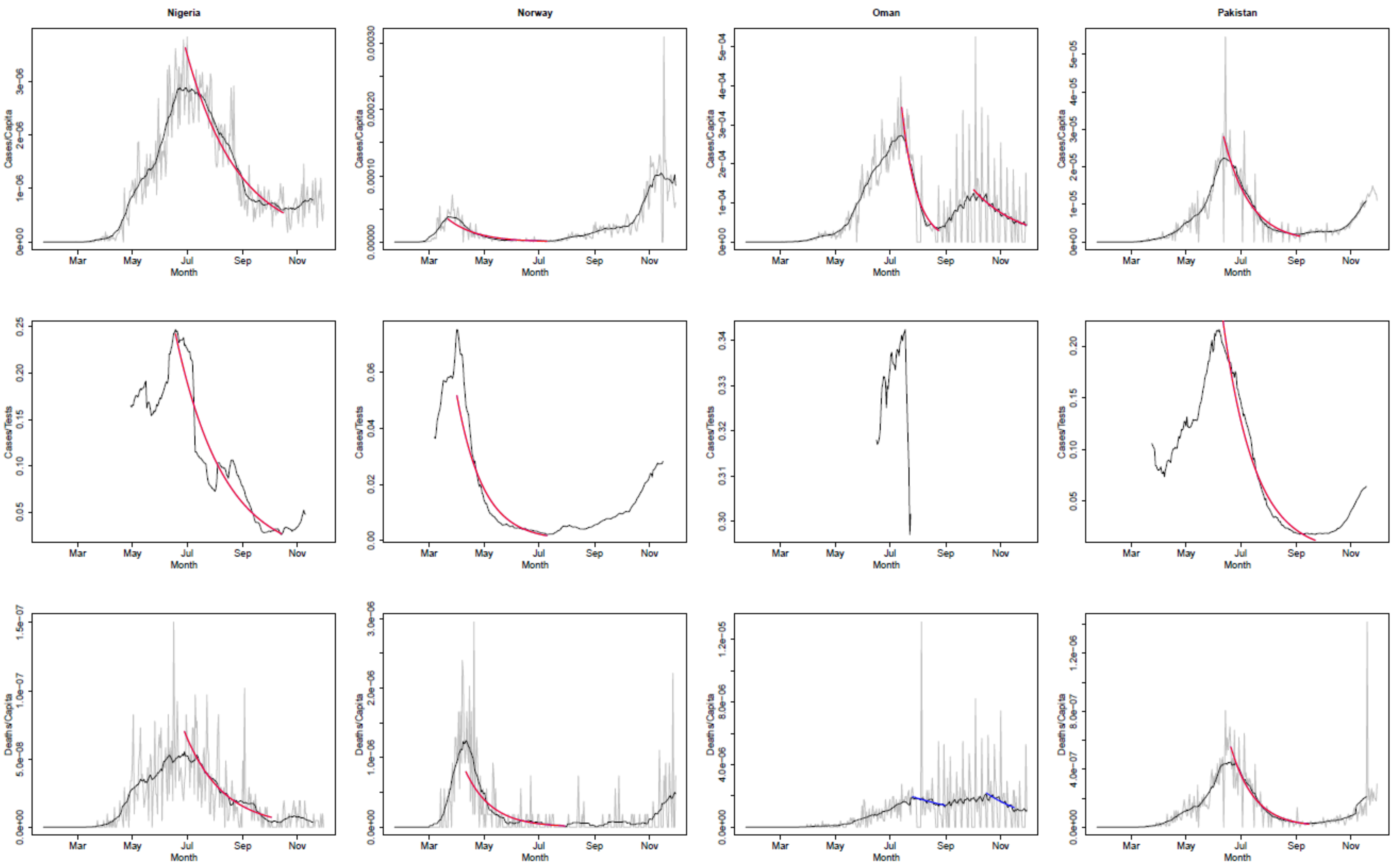
Nigeria
Norway
Oman
Pakistan
Cases/Capita
Cases/Tests
Deaths/Capita
Month
Mar
May
Jul
Sep
Nov

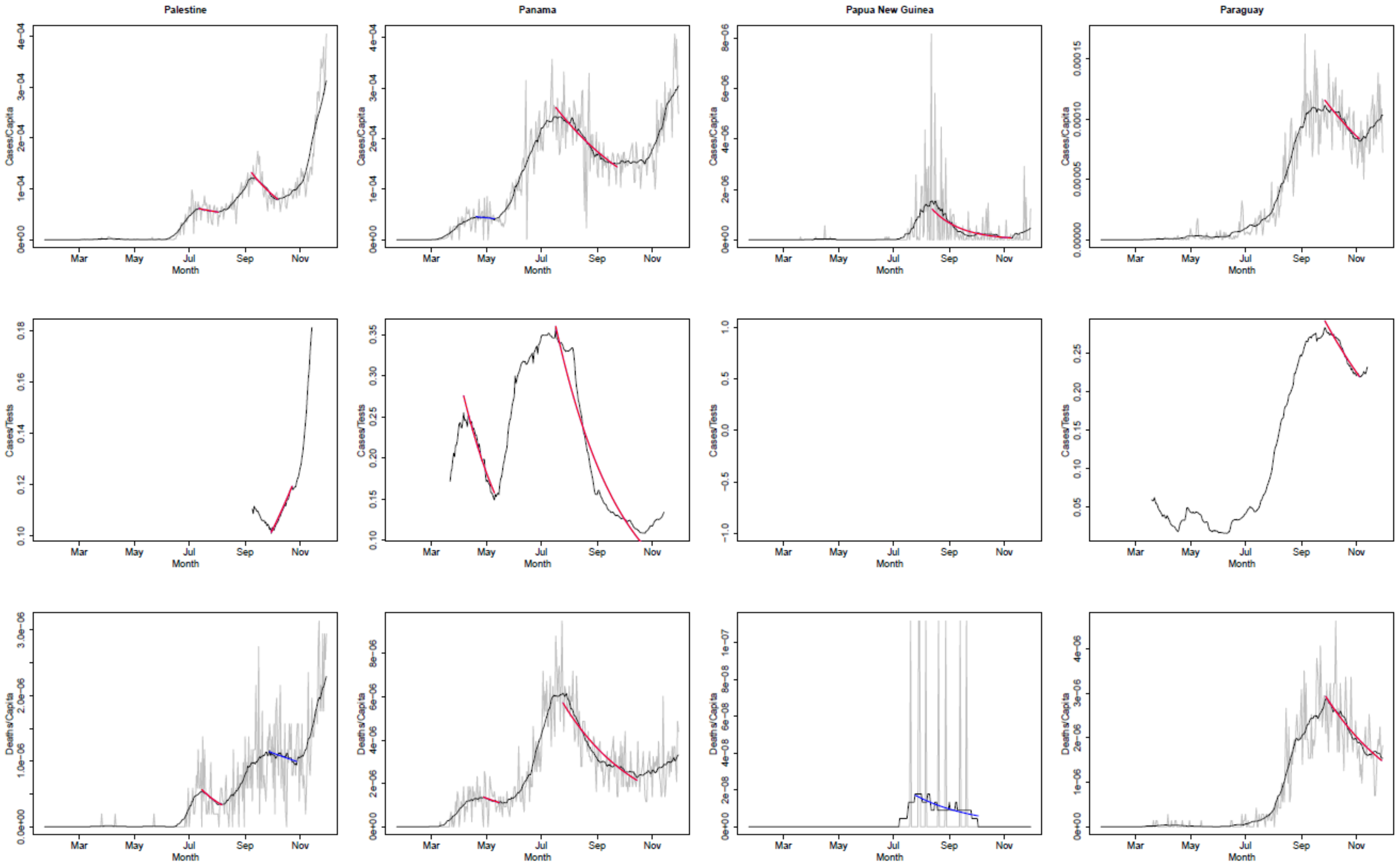
Palestine
Panama
Papua New Guinea
Paraguay
Cases/Capita
Cases/Tests
Deaths/Capita
Month
Mar
May
Jul
Sep
Nov

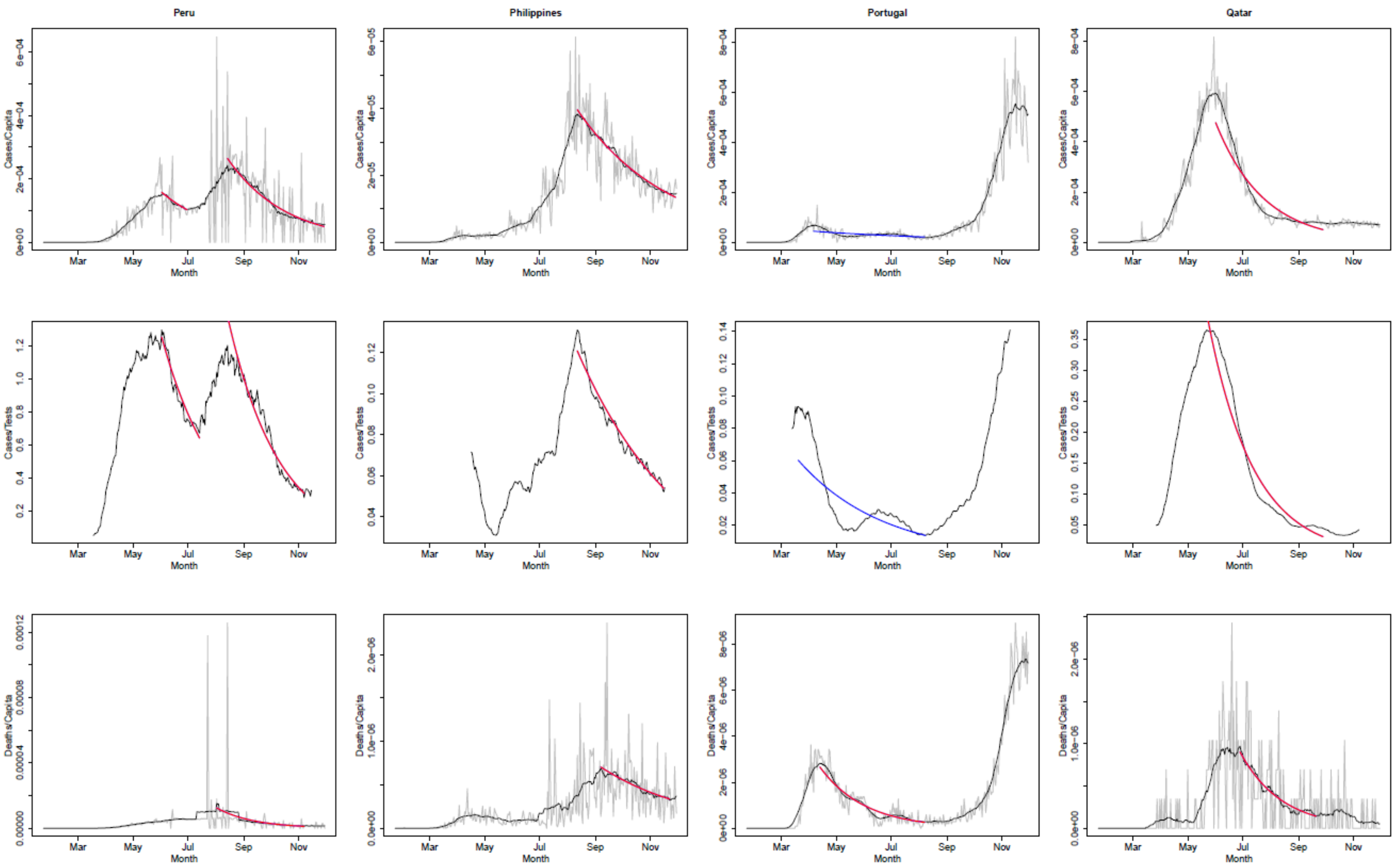
Peru
Philippines
Portugal
Qatar
Cases/Capita
Cases/Tests
Deaths/Capita
Month
Mar
May
Jul
Sep
Nov

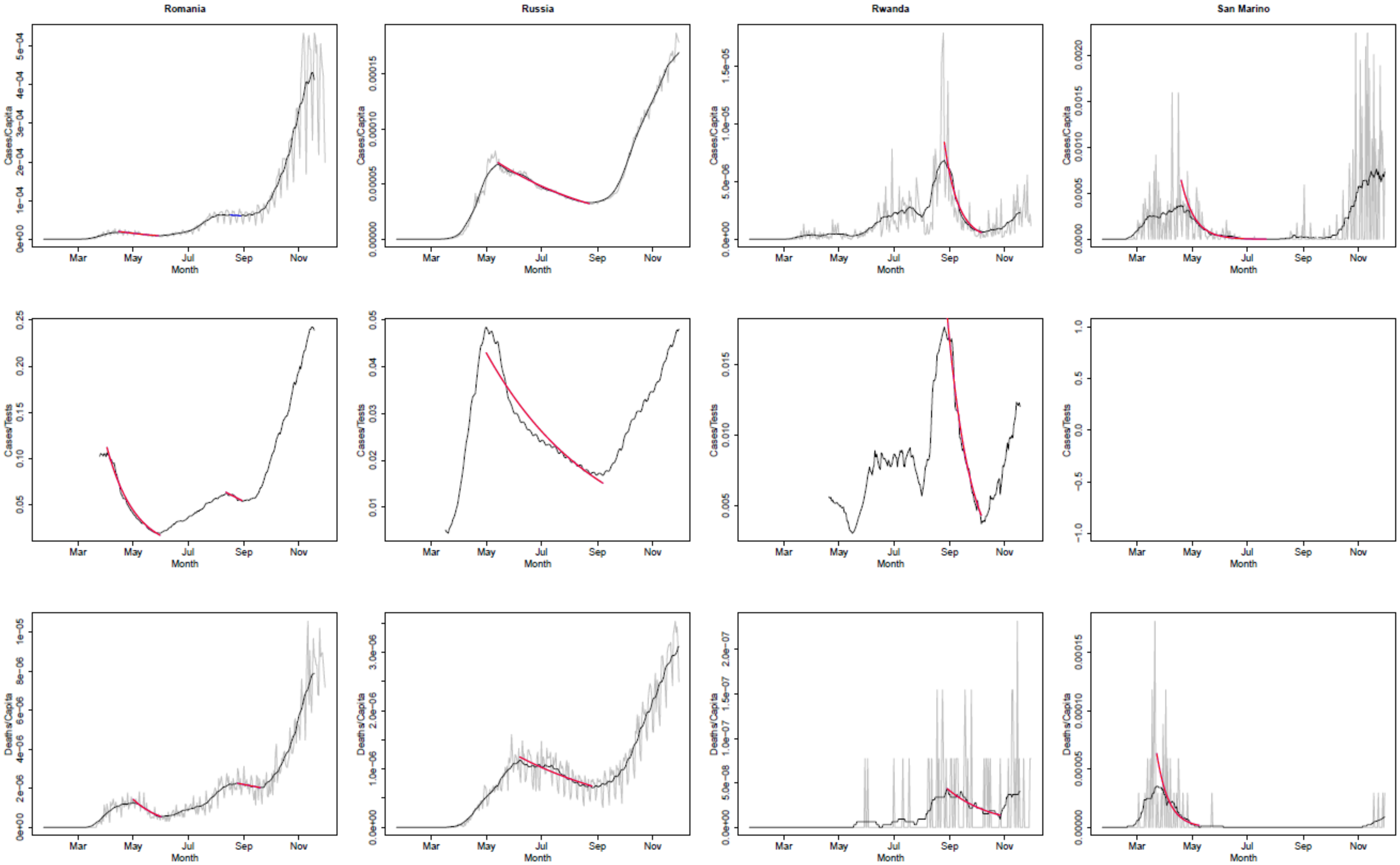
Romania
Russia
Rwanda
San Marino
Cases/Capita
Cases/Tests
Deaths/Capita
Month
Mar
May
Jul
Sep
Nov

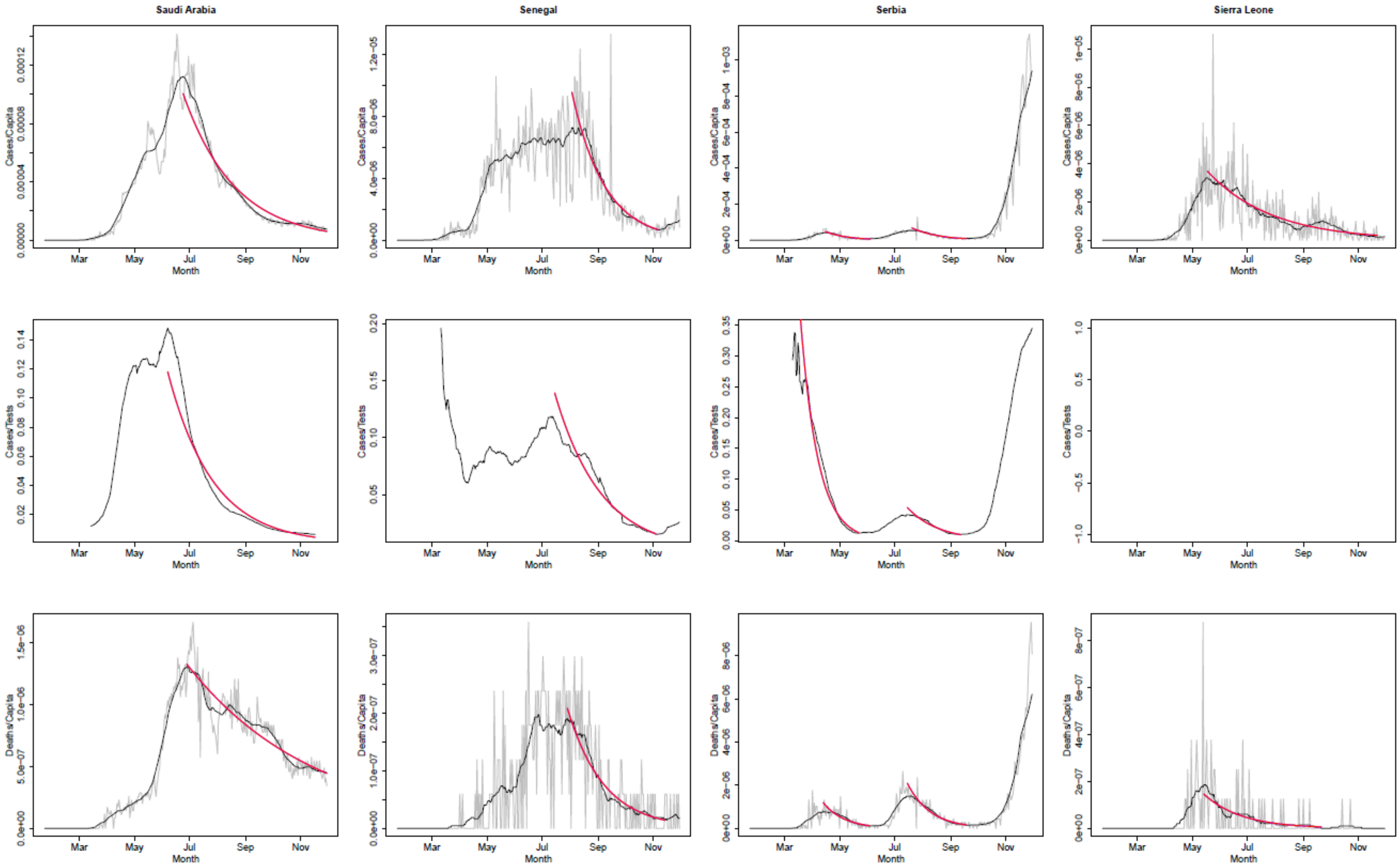

Saudi Arabia
Senegal
Serbia
Sierra Leone
Cases/Capita
Cases/Tests
Deaths/Capita
Month
Mar
May
Jul
Sep
Nov

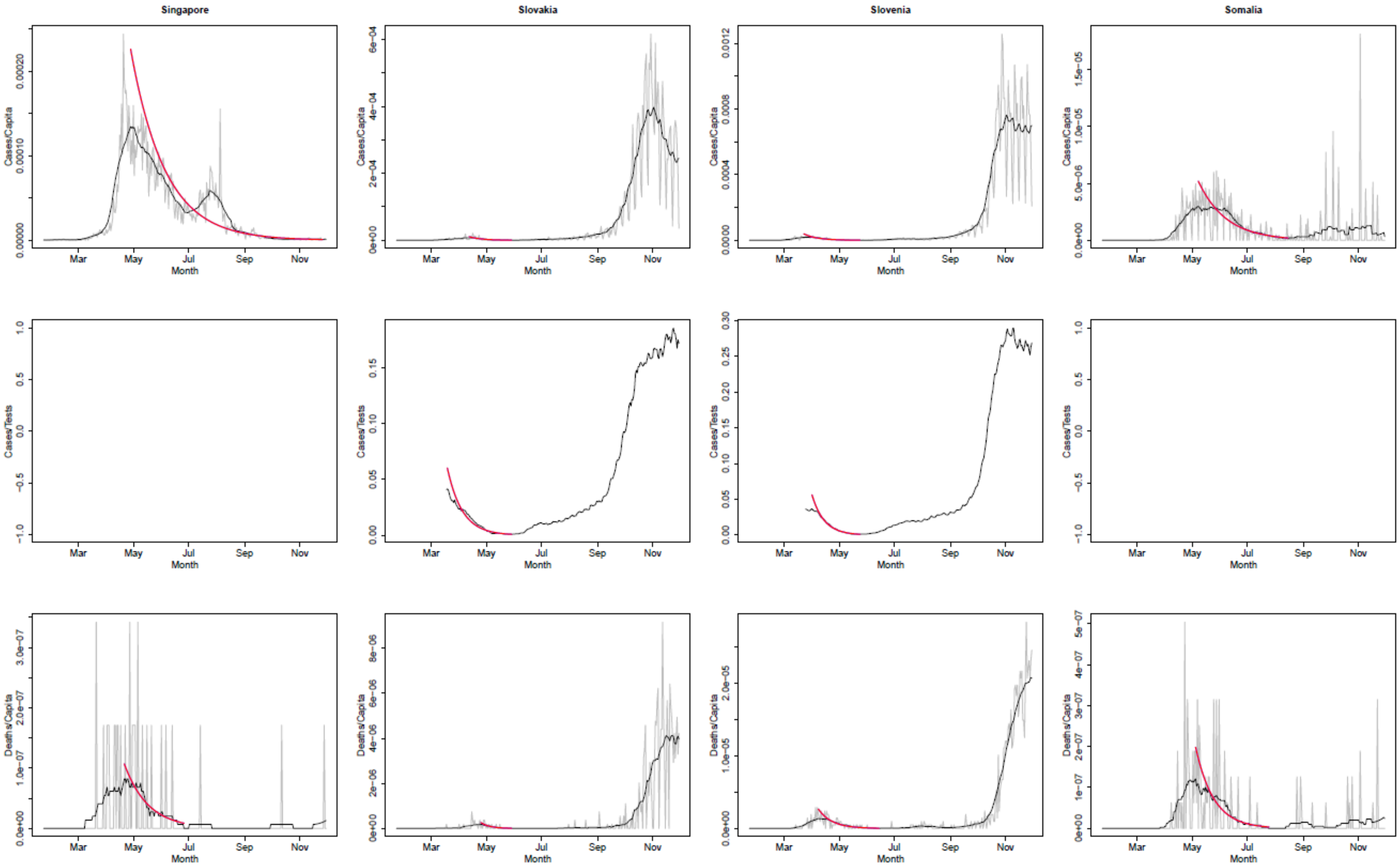
Singapore
Slovakia
Slovenia
Somalia
Cases/Capita
Cases/Tests
Deaths/Capita
Mar
May
Jul
Sep
Nov
Month

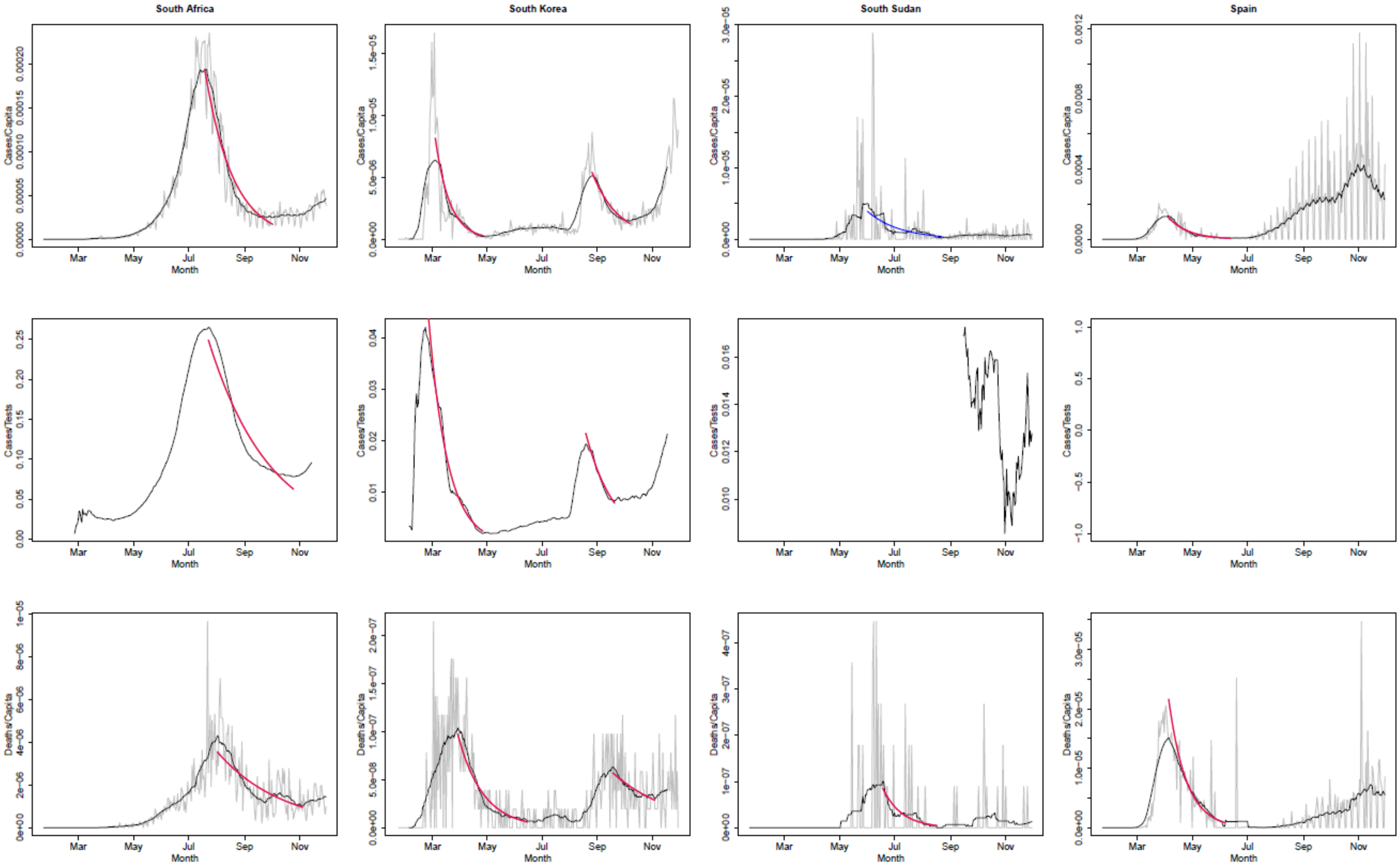
South Africa
South Korea
South Sudan
Spain
Cases/Capita
Cases/Tests
Deaths/Capita
Mar
May
Jul
Sep
Nov
Month

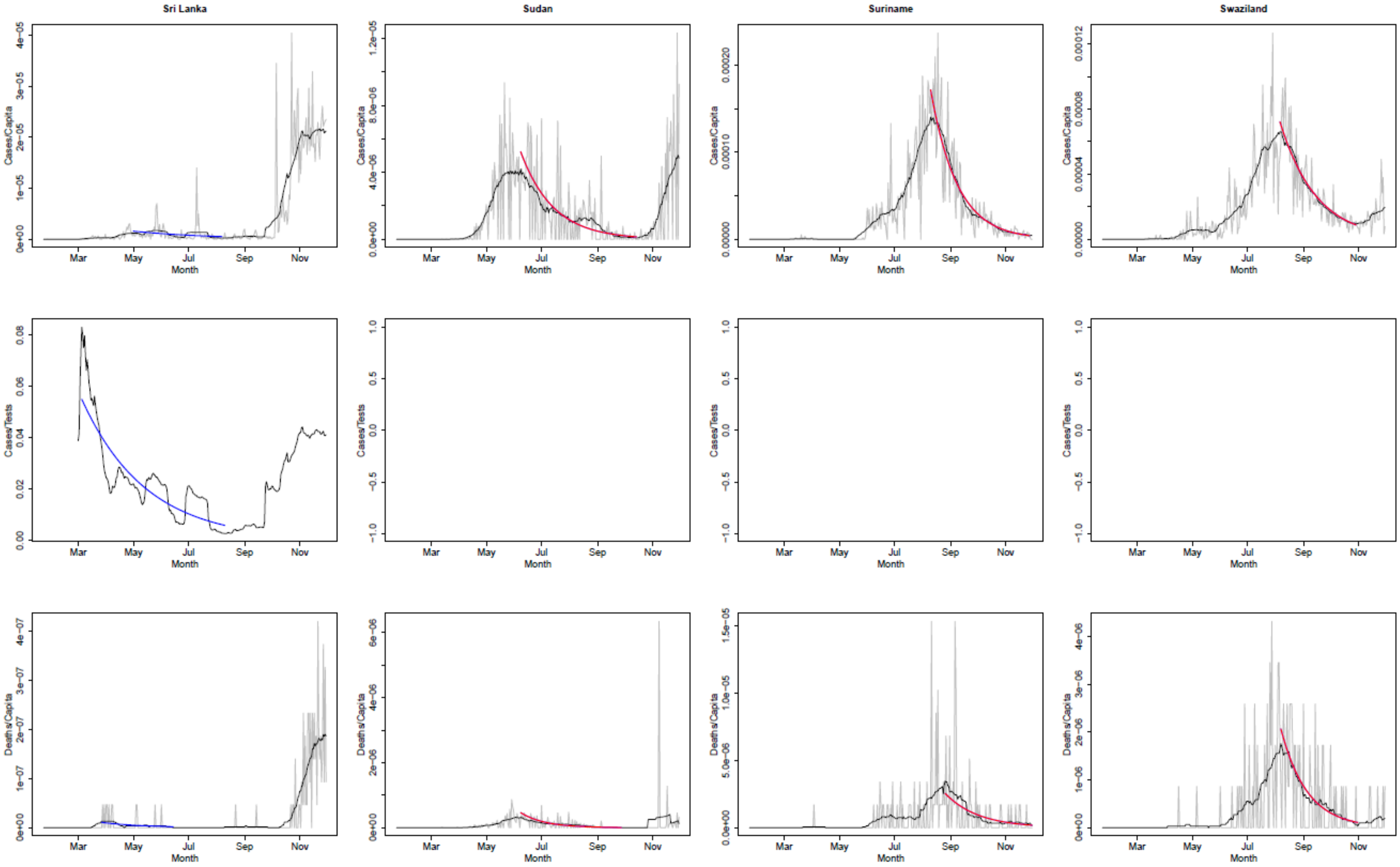

Sri Lanka
Sudan
Suriname
Swaziland
Cases/Capita
Cases/Tests
Deaths/Capita
Month
Mar
May
Jul
Sep
Nov

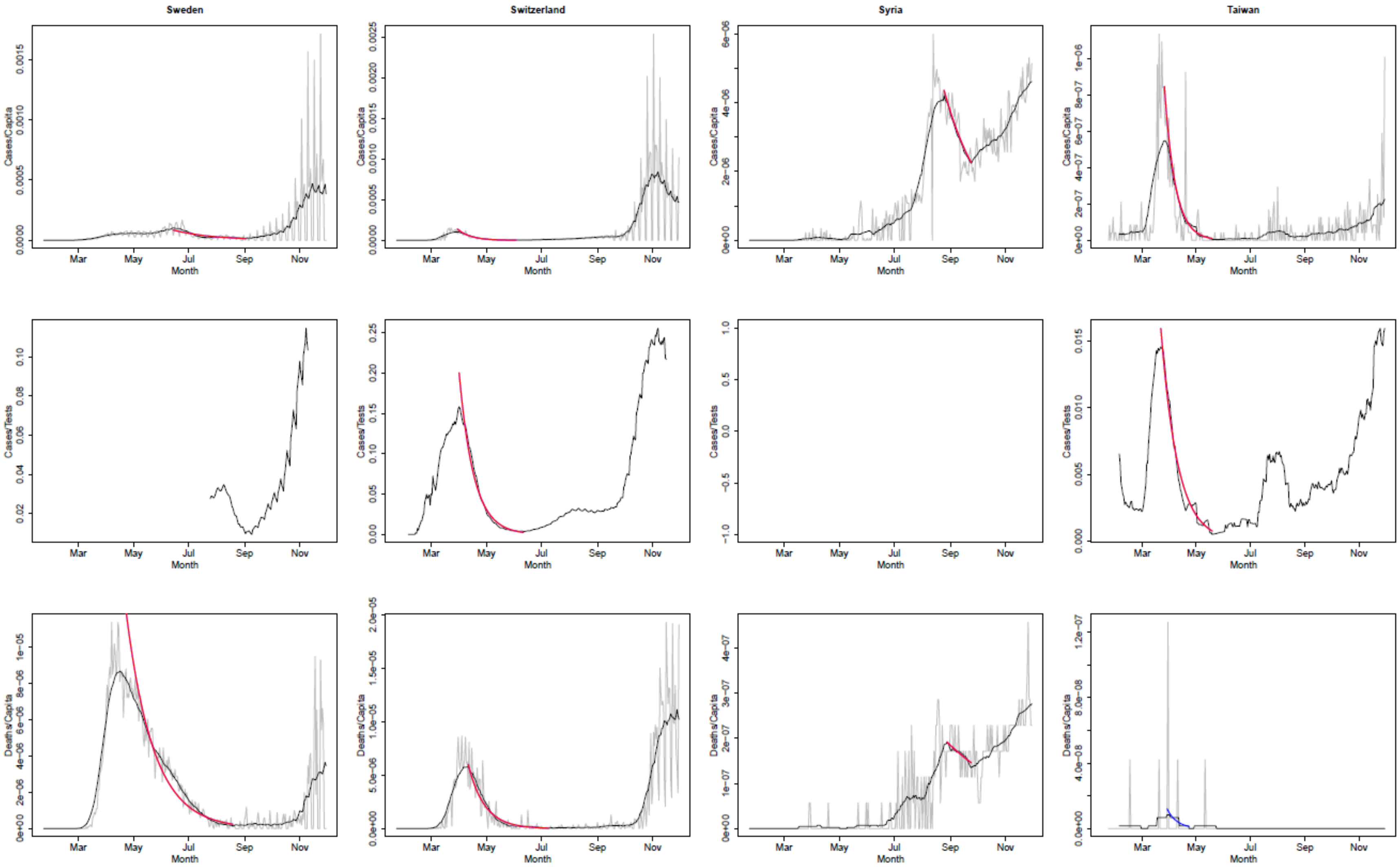
Sweden
Switzerland
Syria
Taiwan
Cases/Capita
Cases/Tests
Deaths/Capita
Month
Mar
May
Jul
Sep
Nov

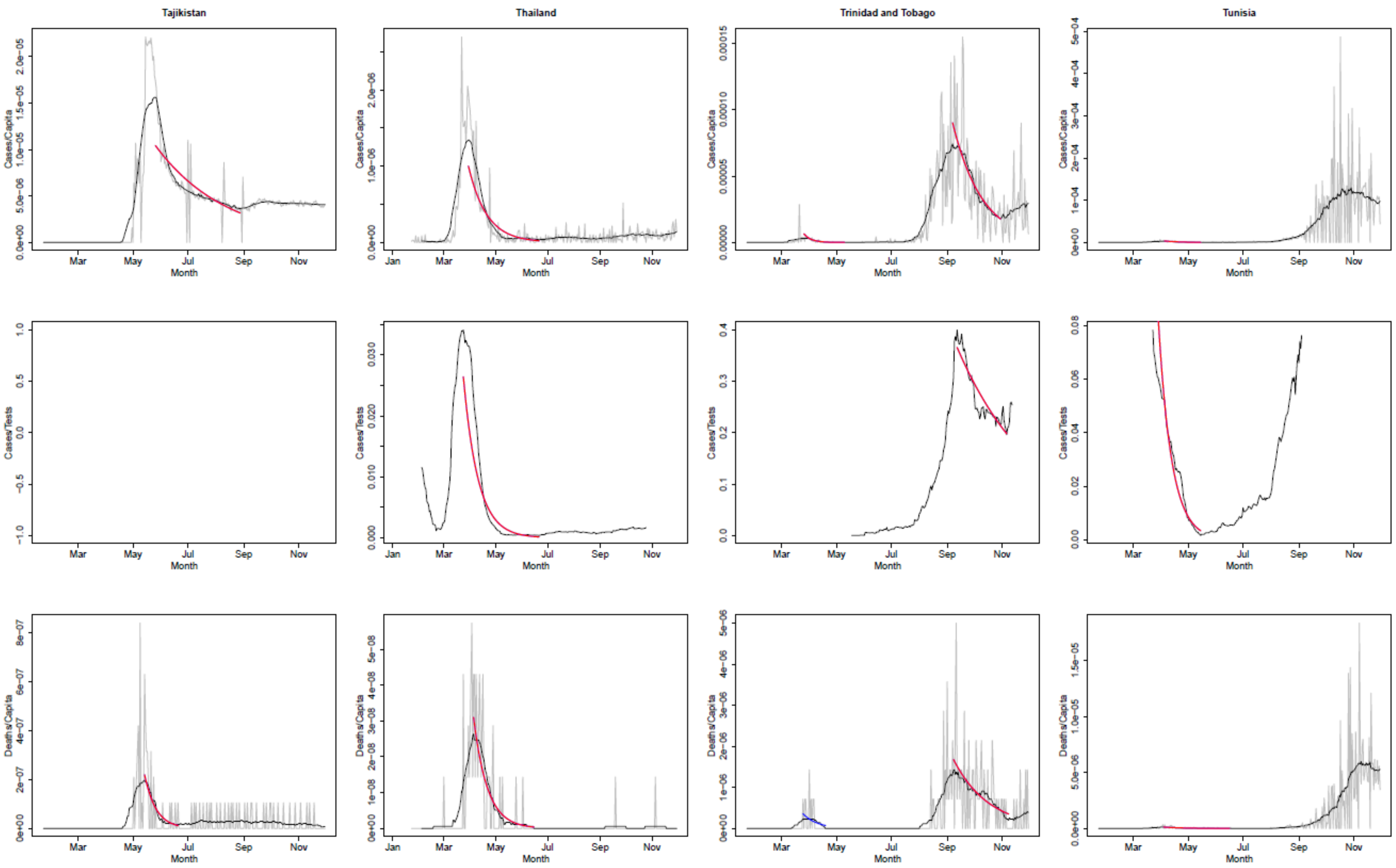
Tajikistan
Thailand
Trinidad and Tobago
Tunisia
Cases/Capita
Cases/Tests
Deaths/Capita
Month
Jan
Mar
May
Jul
Sep
Nov

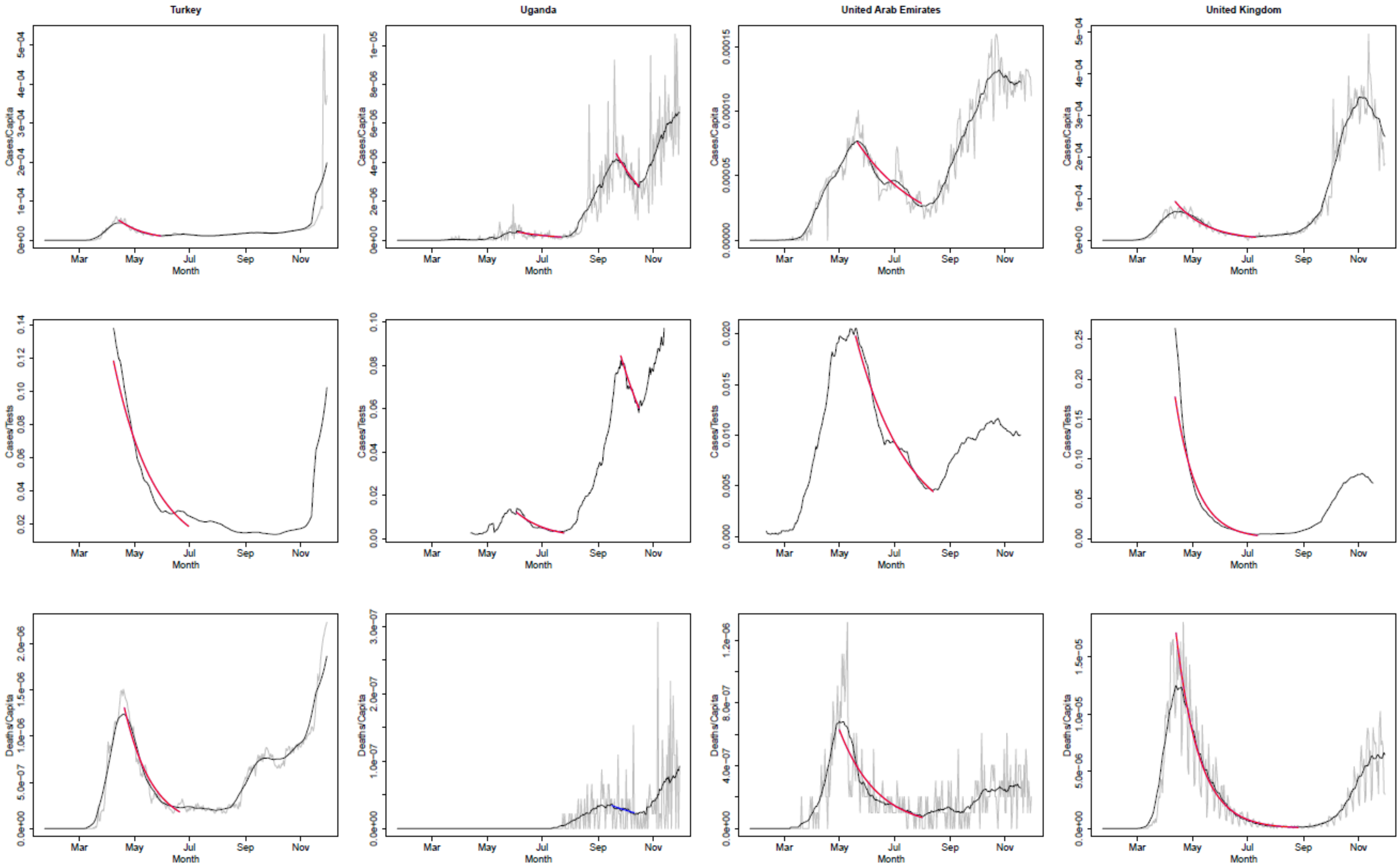
Turkey
Uganda
United Arab Emirates
United Kingdom
Cases/Capita
Cases/Tests
Deaths/Capita
Month
Mar
May
Jul
Sep
Nov

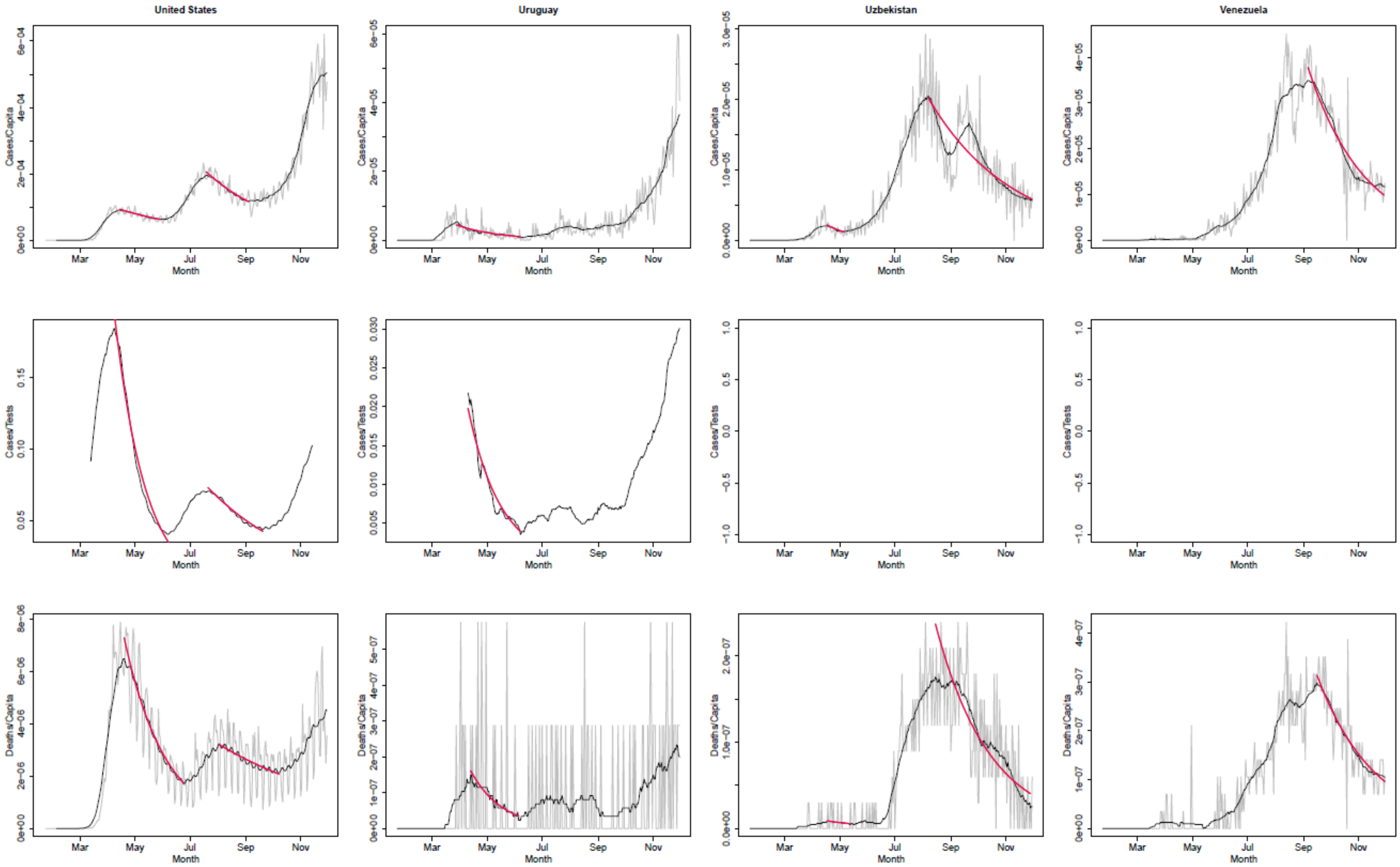
United States
Uruguay
Uzbekistan
Venezuela
Cases/Capita
Cases/Tests
Deaths/Capita
Mar
May
Jul
Sep
Nov
Month

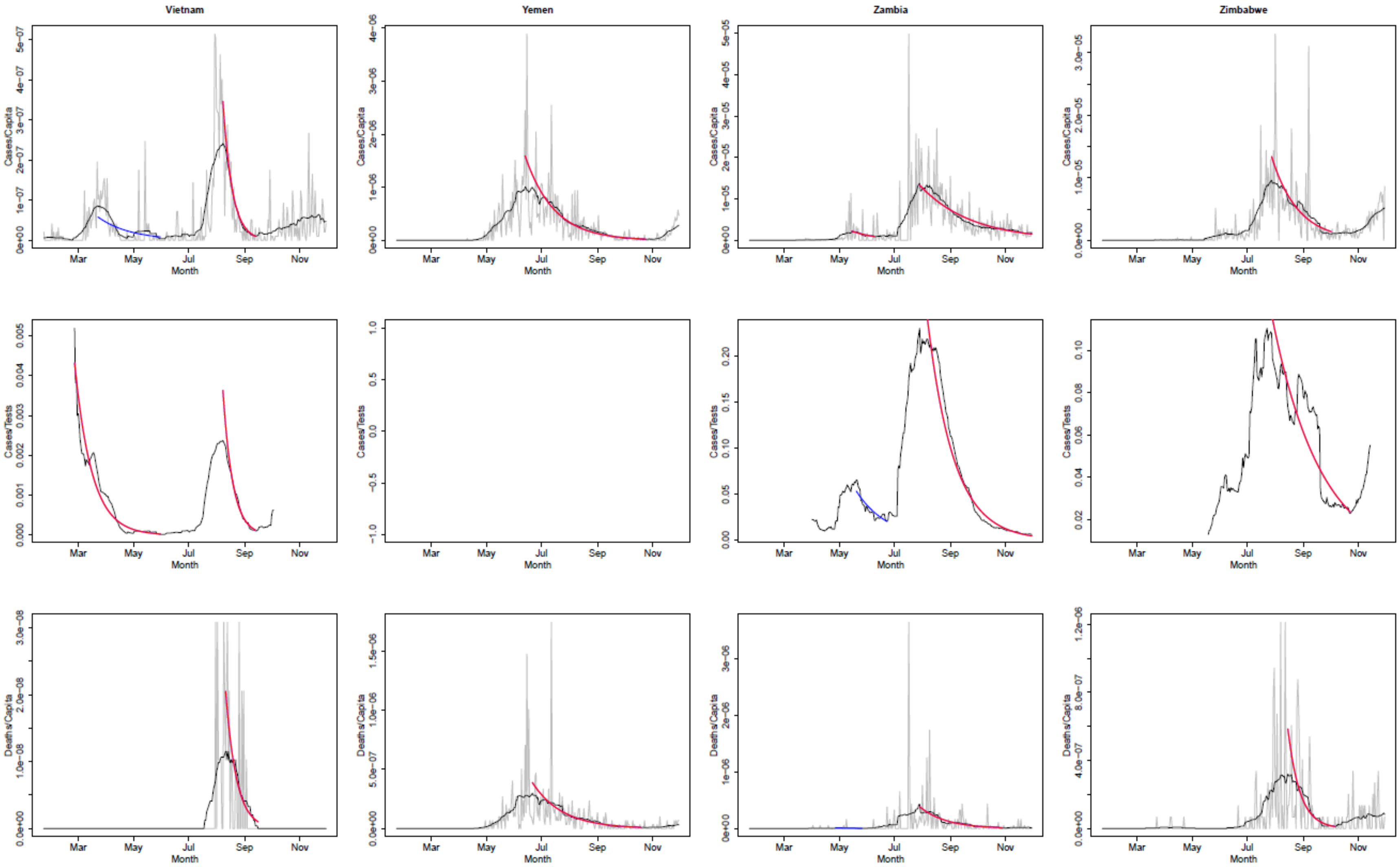

Vietnam
Yemen
Zambia
Zimbabwe
Cases/Capita
Cases/Tests
Deaths/Capita
Month
Mar
May
Jul
Sep
Nov

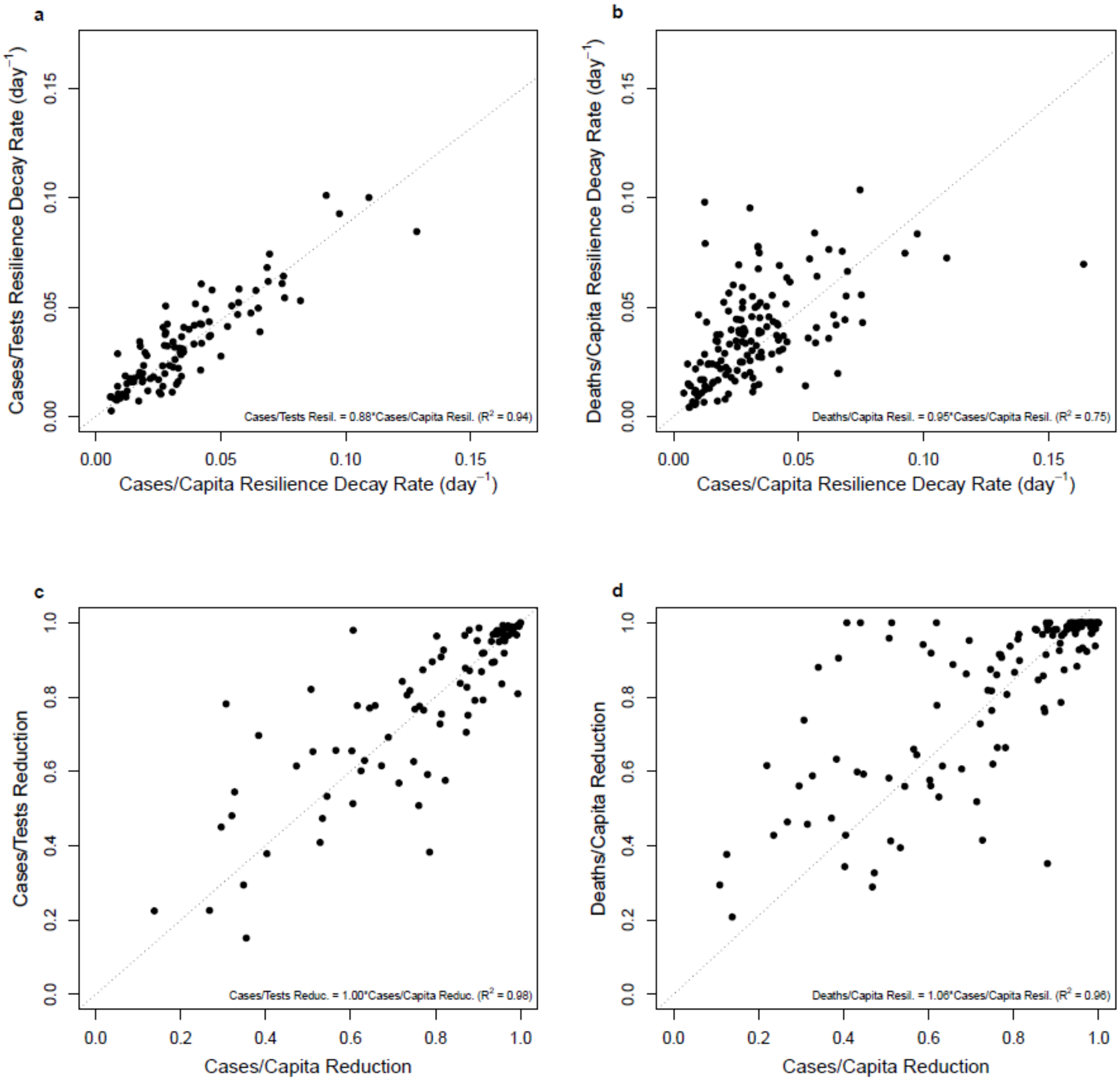

**Fig. S2.**

Comparing different metrics: **a.** resilience of cases/capita vs cases/tests (n=100, $\rho$=0.86, $p<0.0001$). **b.** resilience of cases/capita vs deaths/capita (n=150, $\rho$=0.61, $p<0.0001$). **c.** reduction of cases/capita vs cases/tests (n=94, $\rho$=0.83, $p<0.0001$). **d.** reduction of cases/capita vs deaths/capita (n=136, $\rho$=0.76, $p<0.0001$). Linear correlation results tied to the origin are given within each figure panel and correspond to the dotted lines.

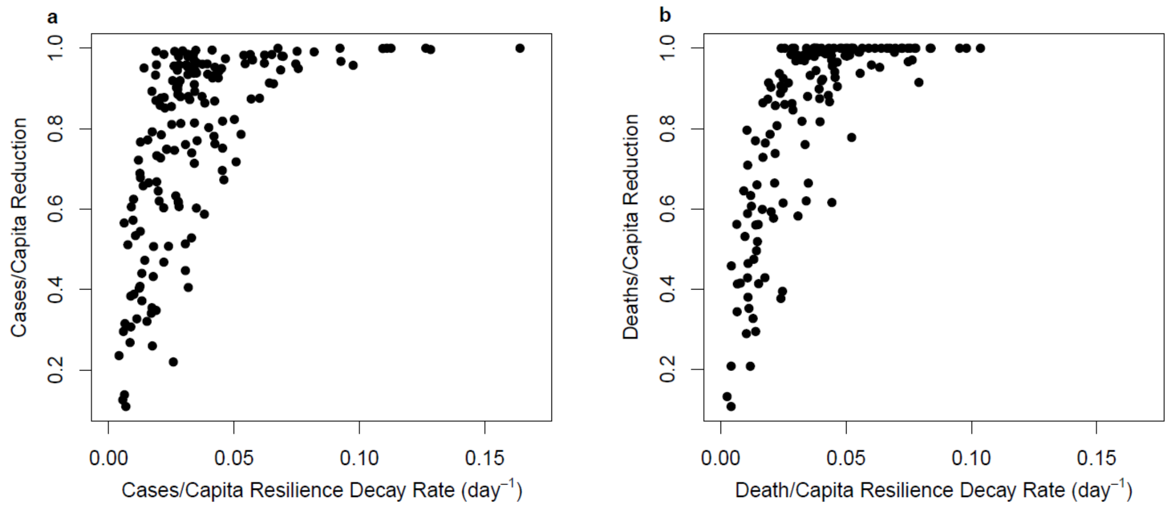

**Fig. S3.**

Comparing resilience to, and reduction of, COVID-19 across countries: **a.** cases/capita (n=165, $\rho$ =0.70, p<0.0001), **b.** deaths/capita (n=150, $\rho$ =0.78, p<0.0001).

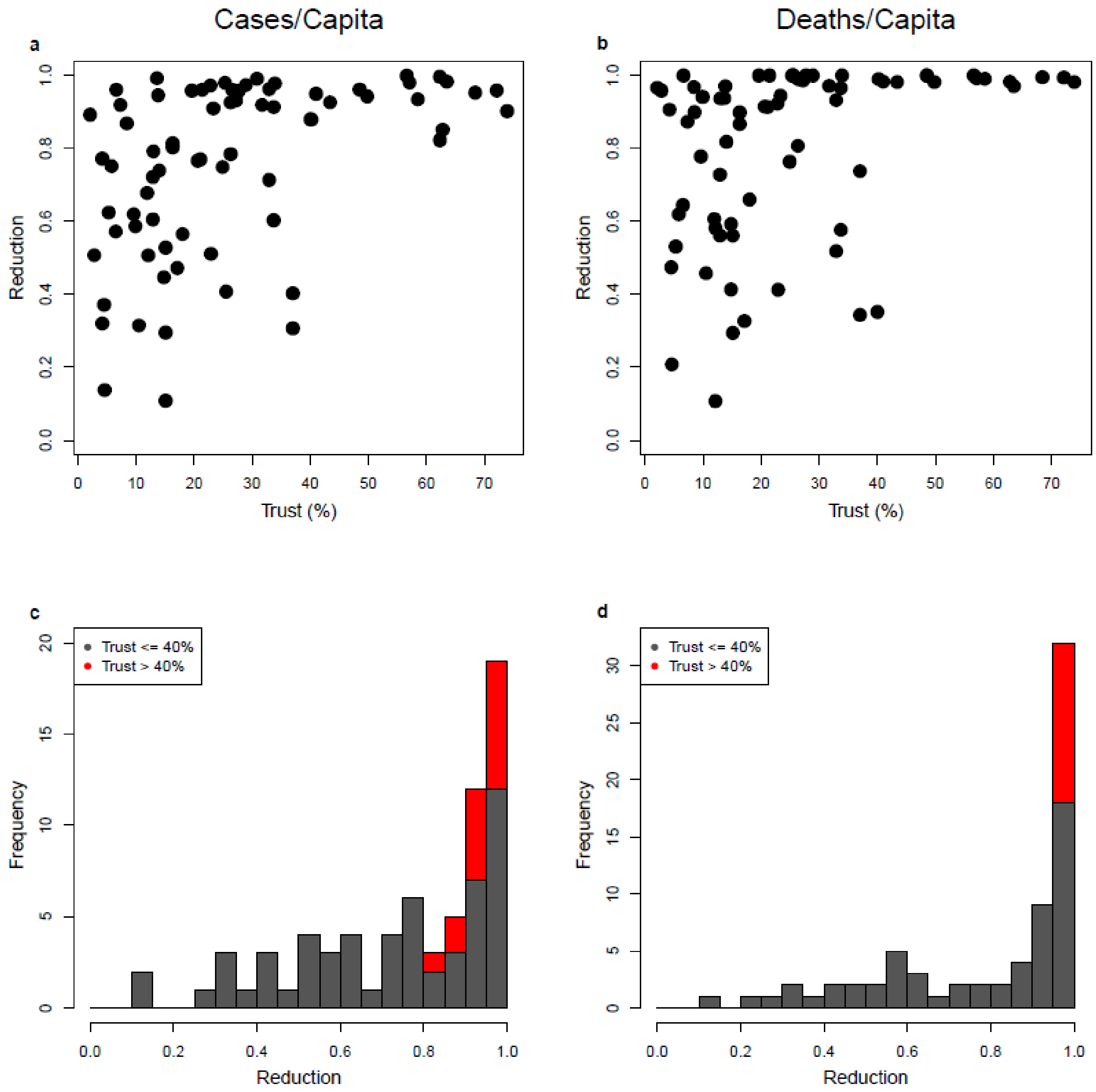

**Fig. S4.**

Relationships between generalized trust within society and reduction of COVID-19 cases and deaths: **a.** cases/capita (n=72, $\rho$=0.51, p<0.0001). **b.** deaths/capita (n=72, $\rho$=0.48, p<0.0001). Stacked frequency distributions partitioning the reduction results into trust ≤40% (grey) and >40% (red): **c.** cases/capita (n=57, n=15; Mann-Whitney U=169, p<0.001). **d.** deaths/capita (n=58, n=14; Mann-Whitney U=132.5, p<0.0001).

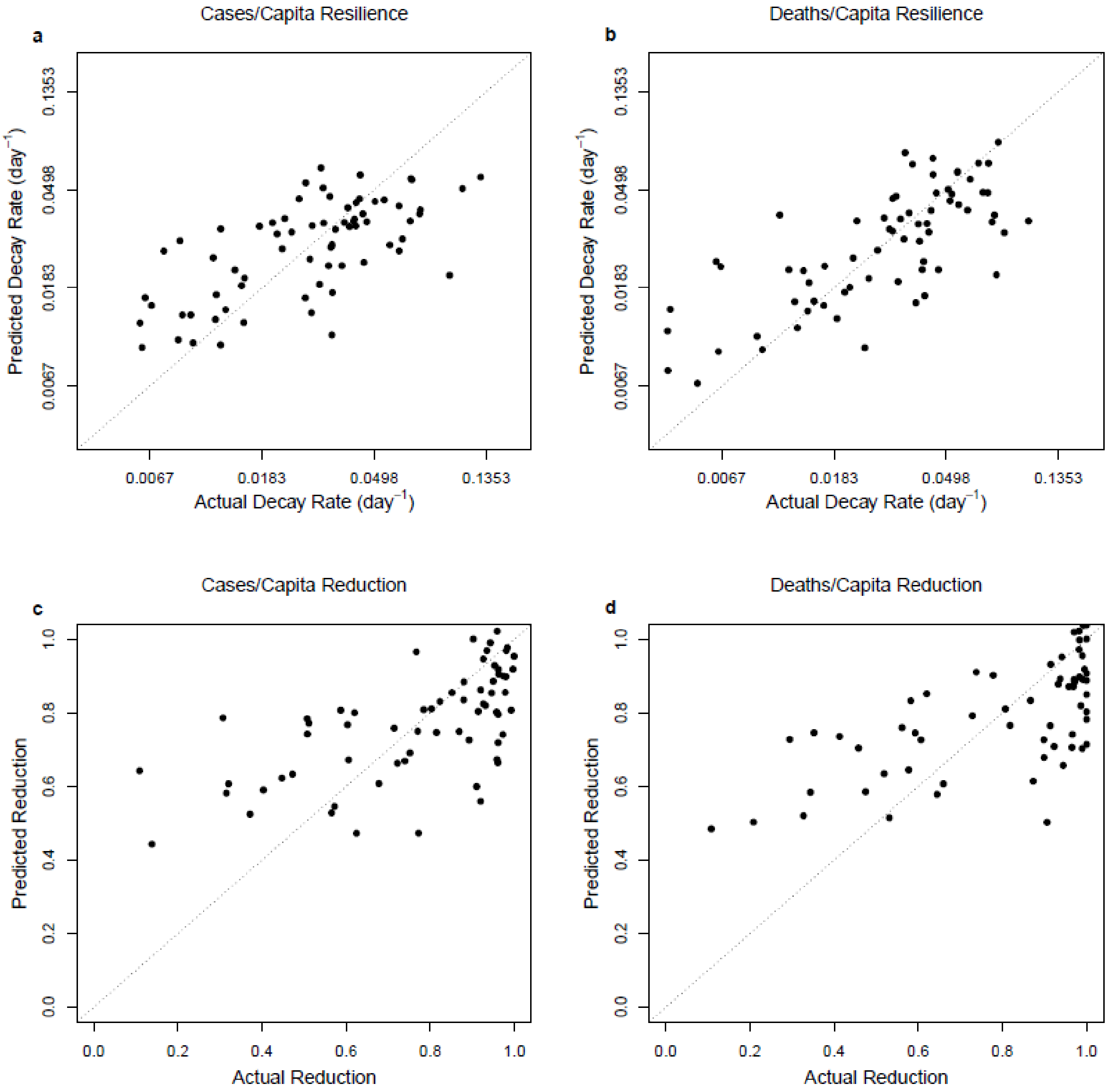

**Fig. S5.**

Optimized multiple linear regression models for: **a.** ln(resilience cases/capita) (n=71, $r^2$=0.409; Table S2). **b.** ln(resilience deaths/capita) (n=69, $r^2$=0.508; Table S3). **c.** cases/capita reduction (n=66, $r^2$=0.352; Table S4). **d.** deaths/capita reduction (n=66, $r^2$=0.414; Table S5).

**Table S1.**

Pairwise Spearman's rank correlations for resilience and reduction for the cases/tests dataset.

| | Resilience | | | Reduction | | |
|---|---|---|---|---|---|---|
| **Explanatory variable** | $\rho$ | **p** | **n** | $\rho$ | **p** | **n** |
| Population | (-0.13) | - | 105 | (-0.14) | - | 101 |
| Country size | (-0.19) | - | 104 | (-0.07) | - | 100 |
| Population density | (0.05) | - | 104 | (-0.11) | - | 100 |
| GDP/capita | 0.24 | <0.05 | 103 | 0.29 | <0.01 | 99 |
| Median age | 0.42 | <0.0001 | 105 | 0.37 | <0.001 | 101 |
| Life expectancy | 0.47 | <0.0001 | 105 | 0.37 | <0.001 | 101 |
| Human Dev. Index | 0.38 | <0.0001 | 104 | 0.35 | <0.001 | 100 |
| Hospital beds | 0.36 | <0.001 | 94 | 0.28 | <0.01 | 90 |
| Mean stringency | -0.39 | <0.0001 | 100 | -0.45 | <0.0001 | 96 |
| Decay stringency | (0.19) | - | 99 | (0.06) | - | 95 |
| Background stringency | -0.48 | <0.0001 | 100 | -0.53 | <0.0001 | 96 |
| Adaptive stringency | 0.67 | <0.0001 | 99 | 0.63 | <0.0001 | 95 |
| Trust | 0.41 | <0.01 | 55 | 0.44 | <0.001 | 54 |
| Power distance | -0.41 | <0.001 | 77 | -0.37 | <0.01 | 74 |
| Individualism | 0.28 | <0.05 | 77 | 0.33 | <0.01 | 74 |
| Masculinity | (0.05) | - | 77 | (-0.02) | - | 74 |
| Uncertainty avoidance | (-0.02) | - | 77 | (-0.17) | - | 74 |
| Long-term orientation | 0.33 | <0.01 | 88 | (0.19) | - | 84 |
| Indulgence | (0.13) | - | 87 | (0.17) | - | 83 |

**Table S2.** Optimized multiple linear regression model for ln(resilience cases/capita) considering trust, adaptive stringency, GDP/capita, population, and hospital beds (n=71, $r^2$=0.409).

| Factor | Coefficient | SE | t | p |
|---|---|---|---|---|
| (Intercept) | -3.239260 | 0.846889 | -3.825 | 0.000293 |
| Adaptive Stringency | 0.012153 | 0.003862 | 3.147 | 0.002477 |
| ln(Population) | -0.073385 | 0.045737 | -1.604 | 0.113382 |
| ln(Hospital beds) | 0.212200 | 0.094458 | 2.246 | 0.028024 |
| Trust | 0.011607 | 0.003735 | 3.107 | 0.002783 |

**Table S3.** Optimized multiple linear regression model for ln(resilience deaths/capita) considering trust, adaptive stringency, GDP/capita, population, and hospital beds (n=69, $r^2$=0.508).

| Factor | Coefficient | SE | t | p |
|---|---|---|---|---|
| (Intercept) | 0.654789 | 1.432163 | 0.457 | 0.64910 |
| ln(GDP/capita) | -0.433278 | 0.131188 | -3.303 | 0.00158 |
| Adaptive Stringency | 0.021039 | 0.003989 | 5.275 | 1.74e-06 |
| ln(Population) | -0.080640 | 0.052499 | -1.536 | 0.12953 |
| ln(Hospital beds) | 0.211839 | 0.110687 | 1.914 | 0.06018 |
| Trust | 0.022932 | 0.004746 | 4.832 | 9.02e-06 |

**Table S4.** Optimized multiple linear regression model for reduction of cases/capita considering trust, adaptive stringency, GDP/capita, population, and hospital beds (n=66, $r^2$=0.352).

| Factor | Coefficient | SE | t | p |
|---|---|---|---|---|
| (Intercept) | 1.262139 | 0.382539 | 3.299 | 0.00162 |
| ln(GDP/capita) | -0.091431 | 0.043852 | -2.085 | 0.04126 |
| Adaptive Stringency | 0.004322 | 0.001307 | 3.306 | 0.00159 |
| ln(Hospital beds) | 0.085271 | 0.036418 | 2.341 | 0.02249 |
| Trust | 0.006781 | 0.001548 | 4.381 | 4.72e-05 |

**Table S5.** Optimized multiple linear regression model for reduction of deaths/capita considering trust, adaptive stringency, GDP/capita, population, and hospital beds (n=66, $r^2$=0.414).

| Factor | Coefficient | SE | t | p |
|---|---|---|---|---|
| (Intercept) | 0.970530 | 0.330757 | 2.934 | 0.004682 |
| ln(GDP/capita) | -0.052773 | 0.036684 | -1.439 | 0.155305 |
| Adaptive Stringency | 0.007198 | 0.001241 | 5.799 | 2.43e-07 |
| Trust | 0.005689 | 0.001560 | 3.647 | 0.000545 |

**Table S6.** Optimized multiple linear regression model for ln(resilience cases/capita) considering Hofstede's six cultural dimensions, adaptive stringency, GDP/capita, population, and hospital beds (n=88, $r^2$=0.382).

| **Factor** | **Coefficient** | **SE** | **t** | **p** |
|---|---|---|---|---|
| (Intercept) | 0.065035 | 0.878650 | 0.074 | 0.941182 |
| ln(Population) | -0.151172 | 0.036012 | -4.198 | $6.94\times10^{-5}$ |
| ln(GDP/capita) | -0.160024 | 0.056346 | -2.840 | 0.005718 |
| Adaptive stringency | 0.013202 | 0.002829 | 4.667 | $1.21\times10^{-5}$ |
| Power distance | -0.006009 | 0.003291 | -1.826 | 0.071628 |
| Masculinity | 0.008514 | 0.003378 | 2.520 | 0.013710 |
| Uncertainty avoidance | -0.004315 | 0.003067 | -1.407 | 0.163351 |
| Long term orientation | 0.009039 | 0.002629 | 3.438 | 0.000933 |

**Table S7.** Optimized multiple linear regression model for ln(resilience deaths/capita) considering Hofstede's six cultural dimensions, adaptive stringency, GDP/capita, population, and hospital beds (n=83, $r^2$=0.376).

| **Factor** | **Coefficient** | **SE** | **t** | **p** |
|---|---|---|---|---|
| (Intercept) | 1.578207 | 1.033408 | 1.527 | 0.13092 |
| ln(Population) | -0.183831 | 0.040633 | -4.524 | $2.23\times10^{-5}$ |
| ln(GDP/capita) | -0.195482 | 0.066158 | -2.955 | 0.00418 |
| Adaptive stringency | 0.014473 | 0.003477 | 4.162 | $8.33\times10^{-5}$ |
| Power distance | -0.007450 | 0.003593 | -2.073 | 0.04158 |
| Masculinity | 0.007022 | 0.003768 | 1.864 | 0.06627 |
| Uncertainty avoidance | -0.009800 | 0.003316 | -2.955 | 0.00417 |
| Long term orientation | 0.006826 | 0.002932 | 2.328 | 0.02261 |

**Table S8.** Optimized multiple linear regression model for reduction of cases/capita considering Hofstede's six cultural dimensions, adaptive stringency, GDP/capita, population, and hospital beds (n=84, $r^2$=0.292).

| **Factor** | **Coefficient** | **SE** | **t** | **p** |
|---|---|---|---|---|
| (Intercept) | 1.9240358 | 0.2863038 | 6.720 | $2.67\times10^{-9}$ |
| ln(Population) | -0.0426342 | 0.0120621 | -3.535 | 0.000690 |
| ln(GDP/capita) | -0.0497983 | 0.0186822 | -2.666 | 0.009339 |
| Adaptive stringency | 0.0036423 | 0.0009931 | 3.667 | 0.000446 |
| Uncertainty avoidance | -0.0027465 | 0.0010402 | -2.640 | 0.010003 |
| Long term orientation | 0.0032869 | 0.0009391 | 3.500 | 0.000772 |

**Table S9.** Optimized multiple linear regression model for reduction of deaths/capita considering Hofstede's six cultural dimensions, adaptive stringency, GDP/capita, population, and hospital beds (n=79, $r^2$=0.374).

| Factor | Coefficient | SE | t | p |
|---|---|---|---|---|
| (Intercept) | 2.138040 | 0.306327 | 6.980 | $1.21x10^{-9}$ |
| ln(Population) | -0.045579 | 0.012150 | -3.751 | 0.000353 |
| ln(GDP/capita) | -0.043789 | 0.019633 | -2.230 | 0.028838 |
| Adaptive stringency | 0.004972 | 0.001104 | 4.505 | $2.51x10^{-5}$ |
| Power distance | -0.002686 | 0.001123 | -2.392 | 0.019391 |
| Masculinity | 0.001808 | 0.001164 | 1.553 | 0.124711 |
| Uncertainty avoidance | -0.002374 | 0.001023 | -2.319 | 0.023209 |

**Table S10.** Optimized multiple linear regression model for ln(resilience cases/capita) considering trust, Hofstede's six cultural dimensions, adaptive stringency, GDP/capita, population, and hospital beds (n=52, $r^2$=0.532).

| Factor | Coefficient | SE | t | p |
|---|---|---|---|---|
| (Intercept) | -2.257826 | 1.003801 | -2.249 | 0.029435 |
| ln(Population) | -0.132911 | 0.055857 | -2.379 | 0.021637 |
| Adaptive stringency | 0.015180 | 0.004181 | 3.631 | 0.000720 |
| Individualism | -0.009176 | 0.004104 | -2.236 | 0.030358 |
| Masculinity | 0.013417 | 0.003759 | 3.569 | 0.000866 |
| Indulgence | -0.008775 | 0.003724 | -2.356 | 0.022887 |
| Trust | 0.024357 | 0.004906 | 4.965 | $1.03x10^{-5}$ |

**Table S11.** Optimized multiple linear regression model for ln(resilience deaths/capita) considering trust, Hofstede's six cultural dimensions, adaptive stringency, GDP/capita, population, and hospital beds (n=52, $r^2$=0.530).

| Factor | Coefficient | SE | t | p |
|---|---|---|---|---|
| (Intercept) | -1.990951 | 1.159329 | -1.717 | 0.09295 |
| ln(Population) | -0.134119 | 0.063093 | -2.126 | 0.03918 |
| Adaptive stringency | 0.017690 | 0.004596 | 3.849 | 0.00038 |
| Individualism | -0.007397 | 0.004792 | -1.543 | 0.12988 |
| Masculinity | 0.013249 | 0.004533 | 2.923 | 0.00546 |
| Long term orientation | -0.008082 | 0.004749 | -1.702 | 0.09583 |
| Indulgence | -0.012791 | 0.005371 | -2.382 | 0.02163 |
| Trust | 0.030971 | 0.006366 | 4.865 | $1.51x10^{-5}$ |

**Table S12.** Optimized multiple linear regression model for reduction of cases/capita considering trust, Hofstede's six cultural dimensions, adaptive stringency, GDP/capita, population, and hospital beds (n=50, $r^2$=0.518).

| **Factor** | **Coefficient** | **SE** | **t** | **p** |
|---|---|---|---|---|
| (Intercept) | 2.192309 | 0.582217 | 3.765 | 0.00049 |
| ln(Population) | -0.036858 | 0.017365 | -2.122 | 0.03946 |
| ln(GDP/capita) | -0.129780 | 0.049491 | -2.622 | 0.01195 |
| Adaptive stringency | 0.004789 | 0.001483 | 3.230 | 0.00234 |
| Long term orientation | 0.003280 | 0.001060 | 3.094 | 0.00343 |
| Trust | 0.007225 | 0.001590 | 4.543 | $4.29 \times 10^{-5}$ |

**Table S13.** Optimized multiple linear regression model for reduction of deaths/capita considering trust, Hofstede's six cultural dimensions, adaptive stringency, GDP/capita, population, and hospital beds (n=51, $r^2$=0.533).

| **Factor** | **Coefficient** | **SE** | **t** | **p** |
|---|---|---|---|---|
| (Intercept) | 2.060966 | 0.621679 | 3.315 | 0.00182 |
| ln(Population) | -0.031950 | 0.018684 | -1.710 | 0.09415 |
| ln(GDP/capita) | -0.121991 | 0.050866 | -2.398 | 0.02068 |
| Adaptive stringency | 0.006631 | 0.001391 | 4.769 | $1.98 \times 10^{-5}$ |
| Masculinity | 0.002239 | 0.001217 | 1.840 | 0.07241 |
| Trust | 0.007950 | 0.001669 | 4.764 | $2.01 \times 10^{-5}$ |

**Table S14.**

Pairwise Spearman's rank correlations for resilience and reduction for the dataset of first peak only in each country.

| | Resilience | | | | | | Reduction | | | | | |
|---|---|---|---|---|---|---|---|---|---|---|---|---|
| | Cases/capita | | | Deaths/capita | | | Cases/capita | | | Deaths/capita | | |
| **Explanatory variable** | $\rho$ | **p** | **n** | $\rho$ | **p** | **n** | $\rho$ | **p** | **n** | $\rho$ | **p** | **n** |
| Population | -0.29 | <0.001 | 143 | -0.32 | <0.001 | 130 | -0.31 | <0.001 | 137 | -0.28 | <0.01 | 123 |
| Country size | -0.30 | <0.001 | 141 | -0.24 | <0.01 | 128 | -0.23 | <0.01 | 135 | -0.21 | <0.05 | 121 |
| Population density | (0.10) | - | 141 | (0.03) | - | 128 | (-0.01) | - | 135 | (0.01) | - | 121 |
| GDP/capita | 0.24 | <0.01 | 138 | (0.09) | - | 126 | (0.12) | - | 132 | (0.08) | - | 119 |
| Median age | 0.27 | <0.01 | 140 | (0.16) | - | 127 | (0.08) | - | 134 | (0.02) | - | 120 |
| Life expectancy | 0.30 | <0.001 | 142 | 0.19 | <0.05 | 129 | (0.10) | - | 136 | (0.06) | - | 122 |
| Human Dev. Index | 0.27 | <0.01 | 139 | (0.14) | - | 127 | (0.10) | - | 133 | (0.08) | - | 120 |
| Hospital beds | 0.33 | <0.001 | 127 | 0.28 | <0.01 | 116 | (0.17) | - | 122 | (0.13) | - | 110 |
| Mean stringency | -0.22 | <0.01 | 136 | -0.42 | <0.0001 | 128 | -0.45 | <0.0001 | 130 | -0.52 | <0.0001 | 121 |
| Decay stringency | 0.19 | <0.05 | 136 | (0.04) | - | 127 | -0.21 | <0.05 | 130 | -0.25 | <0.01 | 120 |
| Background stringency | -0.29 | <0.001 | 136 | -0.54 | <0.0001 | 128 | -0.52 | <0.0001 | 130 | -0.59 | <0.0001 | 121 |
| Adaptive stringency | 0.48 | <0.0001 | 136 | 0.46 | <0.0001 | 127 | 0.19 | <0.05 | 130 | 0.22 | <0.05 | 120 |
| Trust | 0.45 | <0.001 | 67 | 0.42 | <0.001 | 64 | 0.59 | <0.0001 | 63 | 0.55 | <0.0001 | 61 |
| Power distance | -0.36 | <0.001 | 89 | -0.31 | <0.01 | 84 | -0.24 | <0.05 | 88 | -0.32 | <0.01 | 81 |
| Individualism | 0.23 | <0.05 | 89 | 0.32 | <0.01 | 84 | (0.09) | - | 88 | 0.27 | <0.05 | 81 |
| Masculinity | (0.11) | - | 89 | (0.02) | - | 84 | (-0.04) | - | 88 | (0.0) | - | 81 |
| Uncertainty avoidance | (-0.04) | - | 89 | (-0.18) | - | 84 | -0.23 | <0.05 | 88 | -0.31 | <0.01 | 81 |
| Long-term orientation | 0.23 | <0.05 | 104 | (0.18) | - | 95 | (0.06) | - | 101 | (0.03) | - | 91 |
| Indulgence | (0.01) | - | 104 | (0.02) | - | 96 | (0.18) | - | 101 | 0.27 | <0.01 | 92 |

**Table S15.**

Pairwise Spearman's rank correlations for resilience and reduction for dataset of more stringent fits of exponential decay $r^2 \geq 0.9$.

| | Resilience | | | | | | Reduction | | | | | |
|---|---|---|---|---|---|---|---|---|---|---|---|---|
| | Cases/capita | | | Deaths/capita | | | Cases/capita | | | Deaths/capita | | |
| **Explanatory variable** | $\rho$ | **p** | **n** | $\rho$ | **p** | **n** | $\rho$ | **p** | **n** | $\rho$ | **p** | **n** |
| Population | -0.28 | <0.001 | 139 | -0.29 | <0.01 | 120 | -0.22 | <0.05 | 129 | -0.21 | <0.05 | 112 |
| Country size | -0.33 | <0.0001 | 137 | -0.20 | <0.05 | 120 | -0.19 | <0.05 | 127 | (-0.12) | - | 112 |
| Population density | (0.14) | - | 137 | (0.02) | - | 120 | (0.02) | - | 127 | (-0.06) | - | 112 |
| GDP/capita | 0.20 | <0.05 | 133 | (0.06) | - | 117 | (0.08) | - | 123 | (0.10) | - | 109 |
| Median age | 0.30 | <0.001 | 134 | (0.15) | - | 117 | (0.09) | - | 124 | (0.04) | - | 109 |
| Life expectancy | 0.28 | <0.001 | 137 | (0.14) | - | 119 | (0.08) | - | 127 | (0.03) | - | 111 |
| Human Dev. Index | 0.26 | <0.01 | 134 | (0.11) | - | 117 | (0.10) | - | 124 | (0.09) | - | 109 |
| Hospital beds | 0.32 | <0.001 | 122 | 0.30 | <0.01 | 110 | (0.14) | - | 113 | (0.19) | - | 103 |
| Mean stringency | -0.22 | <0.01 | 132 | -0.48 | <0.0001 | 116 | -0.42 | <0.0001 | 123 | -0.56 | <0.0001 | 108 |
| Decay stringency | 0.18 | <0.05 | 132 | (0.01) | - | 114 | (-0.17) | - | 123 | -0.29 | <0.01 | 106 |
| Background stringency | -0.31 | <0.001 | 132 | -0.55 | <0.0001 | 116 | -0.49 | <0.0001 | 123 | -0.61 | <0.0001 | 108 |
| Adaptive stringency | 0.49 | <0.0001 | 132 | 0.45 | <0.0001 | 114 | 0.28 | <0.01 | 123 | 0.24 | <0.05 | 106 |
| Trust | 0.47 | <0.001 | 55 | 0.40 | <0.01 | 58 | 0.48 | <0.001 | 51 | 0.55 | <0.0001 | 55 |
| Power distance | -0.37 | <0.001 | 87 | (-0.22) | - | 77 | -0.25 | <0.05 | 84 | -0.24 | <0.05 | 73 |
| Individualism | (0.17) | - | 87 | 0.26 | <0.05 | 77 | (0.06) | - | 84 | 0.25 | <0.05 | 73 |
| Masculinity | (-0.06) | - | 87 | (-0.02) | - | 77 | (-0.17) | - | 84 | (0.01) | - | 73 |
| Uncertainty avoidance | (0.03) | - | 87 | (-0.18) | - | 77 | (-0.15) | - | 84 | -0.26 | <0.05 | 73 |
| Long-term orientation | 0.28 | <0.01 | 104 | 0.22 | <0.05 | 87 | (0.07) | - | 98 | (0.12) | - | 81 |
| Indulgence | (0.0) | - | 103 | (-0.01) | - | 86 | (0.16) | - | 97 | 0.25 | <0.05 | 80 |
